# A bibliometric and visual analysis of neutrophil to lymphocyte ratio from 2004 to 2023

**DOI:** 10.1101/2025.08.20.25334135

**Authors:** YongCheng Jiang, XiangJun Wu, ZiYang Tao, Hang Li, YiWei Zhu, Lei He

## Abstract

**Background:** The Neutrophil to lymphocyte ratio (NLR) has emerged as a pivotal biomarker in the fields of inflammation and immunity research. NLR, calculated from routine blood counts, reflects the balance between neutrophil-driven innate immunity and lymphocyte-mediated adaptive responses. In recent decades, a growing body of research has underscored the application of this field across diverse medical disciplines, including oncology, cardiology, immunology, and other areas of medicine. However, there is a paucity of comprehensive studies that objectively map the evolving landscape, hotspots, and research frontiers of global NLR studies.

**Methods:** A comprehensive search was conducted using the Web of Science Core Collection to identify articles and reviews related to NLR from 2004 to 2023. A robust bibliometric approach was employed, utilizing tools such as VOSviewer, CiteSpace, and Bibliometrix (R-tool for R Studio). Analyses were focused on publication outputs, country/institutional collaborations, journal and author productivity, and keyword co-occurrence. This approach provides both quantitative and qualitative perspectives on the NLR field.

**Results:** The bibliometric review under consideration included a total of 14,877 publications, which indicates a marked upward trend in annual publications. The United States and China contributed the most papers and maintained leading positions in international collaborations and citation influence. A close examination of the most prolific journals in the field, including Frontiers in Oncology, Medicine, and PLOS One, has revealed their role as pivotal dissemination outlets. A comprehensive analysis of the literature revealed that keywords pertaining to oncology, immunotherapy, inflammation, prognosis, and clinical outcome exhibited a marked prevalence in research themes related to NLR. Of particular note is the escalating convergence of these themes with those concerning cancer immunotherapy, cardiovascular risk, and systemic inflammatory response syndromes. Co-citation and cluster analyses revealed an increasing connection between research on the nucleotide-binding leucine-rich repeat (NLR) proteins and emerging fields, including tumor immunology, metabolic disease, and personalized medicine. Network mapping revealed an increase in intercontinental collaborations and identified key researchers who are influencing the discipline.

**Conclusions:** This is the inaugural bibliometric and visual analysis to methodically map the global research landscape of neutrophil-to-lymphocyte ratio. The study reveals a thriving and multidisciplinary research landscape, with significant concentrations in oncology and immunology, as well as noteworthy advancements into emerging domains such as immunotherapy and precision medicine. By unveiling collaboration patterns, identifying leading contributors, and examining keyword trends, this work offers indispensable guidance for scholars seeking to identify impactful research topics and establish strategic partnerships. These insights not only illuminate the current status of NLR research but also forecast its promising directions, inviting further exploration and cross-disciplinary innovation in the years ahead.

## Introduction

Neutrophils and lymphocytes are the two main leukocytes in human blood. Neutrophils, which are the most abundant circulating leukocytes in the human immune system, accounting for approximately 50–70% of all circulating leukocytes in healthy adults, are polymorphonuclear and phagocytic leukocytes that constitute the first line of defense against invading microorganisms such as bacteria, fungi, and protozoa[1]. They resist pathogens through various mechanisms, including chemotaxis, degranulation, phagocytosis, the release of reactive oxygen species (ROS), granular proteins, and the production and liberation of cytokines[2]. They are also important effector cells during tissue injury-induced inflammation[3]. In addition to their pivotal role in innate immunity, recent studies have shown that neutrophils are also involved in immune regulation during both innate and adaptive immune responses[4]. They can secrete a number of pro-inflammatory and immunomodulatory cytokines and chemokines, which are capable of enhancing the recruitment and effector functions of other immune cells[5], such as dendritic cells (DCs), B cells, NK cells, CD4^+^, CD8^+^, and γδTcells, as well as mesenchymal stem cells. Consequently, neutrophils play a vital role in sepsis, infections, and cancer development[6], and they are the main cells in systemic inflammatory response syndrome (SIRS)[7].

Lymphocytes, which play the dominant role in adaptive immunity, are primarily involved in eliminating viral and intracellular bacterial infections. However, the number of lymphocytes is significantly reduced in severe diseases such as bacteraemia, infectious shock, polytrauma, and cancer[8]. The exact mechanism remains to be elucidated.

The Neutrophil to lymphocyte ratio (NLR), calculated as a simple ratio between the neutrophil and lymphocyte counts measured in peripheral blood, is a biomarker that conjugates two faces of the immune system: the innate immune response, mainly mediated by neutrophils, and adaptive immunity, supported by lymphocytes[9]. It was first proposed by Zahorec over twenty years ago as a tool to measure immune-inflammatory responses and neuroendocrine stress[10]. Zahorec found that the most severe critical illnesses were associated with significant increases in neutrophil counts and sharp decreases in lymphocyte counts. Since then, NLR has been used as a marker of systemic inflammatory response syndrome (SIRS) and stress in critically ill patients, to evaluate the severity of sepsis and systemic infections, including bacteremia and infectious shock[11].

Later, additional conditions leading to elevated neutrophils and decreased lymphocytes were identified, including bacterial or fungal infections, acute stroke, myocardial infarction, atherosclerosis, severe trauma, cancer, post-surgery complications, and any condition characterized by tissue damage activating SIRS. Several years later, NLR was found to serve as a predictor for a wide range of tumours, including but not limited to colorectal, breast, lung, prostate, and pancreatic cancers[12]. Walsh et al. were the first to apply this parameter for the prognosis of cancer patients undergoing colorectal surgery[13]. Through a cohort study, they identified a preoperative NLR >5 as a predictor of poor prognosis in colon cancer. Multiple studies have shown that NLR reflects the balance between pro-tumor inflammatory status[14] and antitumor immune status. Patients with elevated NLR exhibit relative lymphocytopenia and neutrophilic leukocytosis, indicating a shift toward pro-tumor inflammation associated with poor oncologic outcomes. Additionally, elevated NLR may alter the tumour microenvironment, thereby promoting tumour growth[6].

Recently, studies have linked elevated NLR to adverse prognoses in cardiovascular disease, diabetic complications, and rheumatic diseases. Following the COVID-19 outbreak, researchers found that NLR was significantly elevated in patients with coronavirus disease 2019 (COVID-19) compared to healthy individuals—a pattern contrasting with other forms of pneumonia[15]. Furthermore, NLR may predict the prognosis of patients infected with the SARS-CoV-2 virus[16]. In conclusion, NLR has become a versatile biomarker for diverse diseases[17] and plays an increasingly important role in medical practice.

Bibliometric analysis is a method that uses mathematical and statistical approaches to review and analyze studies in a specific research field over a defined period, conducting both qualitative and quantitative assessments[18]. This methodology focuses on examining countries, institutions, journals, authors, and keywords associated with research in a particular domain, offering readers an objective perspective on trends, hotspots, and frontiers in the field[19,20]. Bibliometric analysis has been widely applied across numerous research areas, including innate immunity[21], neutrophils, neutrophil extracellular traps (NETs)[22], inflammatory biomarkers, and immunotherapy. Existing bibliometric studies on the Neutrophil to lymphocyte ratio (NLR) have primarily focused on its correlation with specific medical conditions, such as rheumatic diseases and febrile seizures. However, despite the rapid advancement of neutrophil and lymphocyte-related research over the past two decades, there remains a notable gap in comprehensive bibliometric studies systematically organizing NLR-related investigations. Therefore, this study aims to analyze the global landscape of NLR-related research and identify emerging trends and frontier hotspots during the last two decades using three bibliometric software tools: VOSviewer, CiteSpace, and R. The findings may provide researchers with valuable insights to understand field dynamics, identify collaborative opportunities, and guide future research directions.

## Materials and methods

### Data sources

The data for the bibliometric and visual analysis of this study were obtained from the Web of Science Core Collection (WOSCC), a comprehensive, standardized database widely used in academia. In WOSCC, TS stands for Topic Sentence. The retrieval formula which was used in this study is “TS=neutrophil to lymphocyte ratio OR TS=neutrophil lymphocyte ratio”. The search period was limited to January 1, 2004 to December 31, 2023. Only “Article” and “Review” were selected as article types, and the language was limited to English, resulting in 14877 articles. The results were exported as plain text files in txt and CSV formats, according to the above formula for searching on WOSCC. The search was completed on September 22, 2024, to prevent data bias due to database updates.

### Data analysis and visualization

CiteSpace is currently the most widely used software for bibliometric analysis which is developed by Chaomei Chen in 2004. We used CiteSpace 6.1. R2 Advanced visualization to analyze country distribution and collaboration, the dual-map overlay of journals, institutional distribution, subject area distribution, keyword timeline graphs, reference collaboration and literature bursts. VOSviewer was developed by Nees Jan van Eck et al. and is mainly used for bibliometric network graph analysis[23]. We used VOSviewer 1.6.18 to visually analyze country distribution, institution distribution, author distribution and collaboration and keyword collaboration. The clustering, which relies on the similarity matrix and VOS mapping technique, was completed automatically and the corresponding labels were then added by the authors according to the content. In addition, we used Bibliometrix (R-Tool of R-Studio) to visually analyze the country distribution, references and keywords, and Microsoft Excel 365 to show the publication and citation trends of the literature over the years. Finally, we used MATLAB [R2024a] software to predict the number of NLR-related publications. All raw data used in this study were obtained from public databases and did not involve any human participants’ data. Therefore, this study did not require ethical review.

## Results

### Annual publications and citation trends

Figure 1A illustrates the number of annual publications and annual citations of articles relating to Neutrophil to lymphocyte ratio (NLR) in the major scientific journals of the world from 2004 to 2023, along with their respective trends. According to Fig. 1A, the number of annual publications pertaining to the Neutrophil to lymphocyte ratio (NLR) has been on the rise, reaching a peak in 2022. Moreover, the number of annual publications of the NLR-related literature have increased rapidly after 2013. It has undergone rapid growth in the five-year period between 2013 and 2018, exceeding 2000 for the first time by the end of 2021. Besides, regarding to the frequency of citations, the annual number of citations of researches related to NLR has also demonstrated an upward trend from 2004 to 2023. The most pronounced growth occurred between 2013 and 2021. While the rate of annual citations in the two subsequent years has exhibited a slight deceleration in comparison to the preceding period.

The graphs in Figure 1B show the model curves for the annual number of publications and annual citation frequency of NLR-related literature fitted with five statistical models, which is Linear, Quadratic, Exponential, Gaussian, and Cubic, at 95% and 99% confidence intervals. Among them, the Gaussian model (pink curve) possessed the best fit (R^2^= 0.985 and 0.98704 respectively) in the statistical analyses of both the number of publications and the number of citations. The results show that the annual publication volume of NLR-related literature is expected to exceed 2,500 articles around 2025 and will remain at a high level. There is a high probability that the annual citation volume of NLR-related literature will exceed 60,000 times per year for the first time in the next two years and will see a continuous growth.

### Distributions of countries/regions

Currently, a total of 56 countries/regions have participated in the publication of NLR-related researches, mainly in East Asia, Europe, Turkey and the United States. Country/region linkages occur primarily among European countries, North America, East Asia, Australia and the United States. Table 1 shows the top 10 countries/regions for NLR-related publications, as well as the frequency and centrality of their publications. The top three countries are China, Turkey and the United States. It is worth mentioning that China is far ahead in the number of publications. The top three countries cited were China, the United States and Turkey. In addition, publication centrality in the UK was 0.37, which is the highest, followed by Germany at 0.36. Figures 2A and 2B show the collaboration among the top 47 countries for NLR-related literature publication (Vosviewer software mapping). The size of the radius of the node represents the number of publications published by the country/region, and the number and thickness of the lines connecting the countries or regions represent the degree of international cooperation between the two places. In figure 2A, the colored nodes represent the countries/regions where the clusters are located. In figure 2B, the colored nodes are used to indicate when the NLR-related literature was published in the country/region. Figure 2C shows the degree of cooperation and communication between countries on the world map (R software mapping). In figure 2D, the radius of the red node still represents the number of publications in the country/region, and the thickness of the surrounding purple circle is positively correlated with the centrality of publications in that region. The number and color of the lines connecting the two places represent the number of interactions and the corresponding year. It is easy to find that the focus of international exchanges and cooperation still takes place among European countries.

**Table 1.** Top 10 countries/regions in terms of number of publications, the corresponding frequency of citations and centrality.

| rank | countries/regions | publications | citations | Centrality |
| --- | --- | --- | --- | --- |
| 1 | CHINA | 4959 | 97887 | 0.06 |
| 2 | TURKEY | 2230 | 35890 | 0.06 |
| 3 | USA | 1627 | 58020 | 0.14 |
| 4 | JAPAN | 1264 | 29202 | 0 |
| 5 | ITALY | 873 | 25276 | 0.03 |
| 6 | SOUTH KOREA | 725 | 16947 | 0 |
| 7 | ENGLAND | 505 | 21619 | 0.37 |
| 8 | SPAIN | 373 | 13081 | 0.3 |
| 9 | GERMANY | 358 | 11600 | 0.36 |
| 10 | AUSTRALIA | 310 | 14655 | 0.03 |

### Distribution of Institutions

Table 2 lists the top ten institutions according to the number of publications and the corresponding centrality of NLR-related publications. It can be seen that two of the top three institutions in terms of the number of articles are from China. Sun Yat-sen University is at the top of the list(344), and Fudan University is at the third place(256). In addition, Capital Medical University and Huazhong University of Science and Technology have the highest centrality, both at 0.03. SUN YAT SEN UNIVERSITY, UNIVERSITY OF HEALTH SCIENCES TURKEY, FUDAN UNIVERSITY, NANJING MEDICAL UNIVERSITY and SHANGHAI JIAO TONG UNIVERSITY are Identical in terms of centrality, both at 0.02, which is Slightly smaller than the two organizations above. It implies that these institutions occupy a significant position in research in the field of NLR.

**Table 2.**
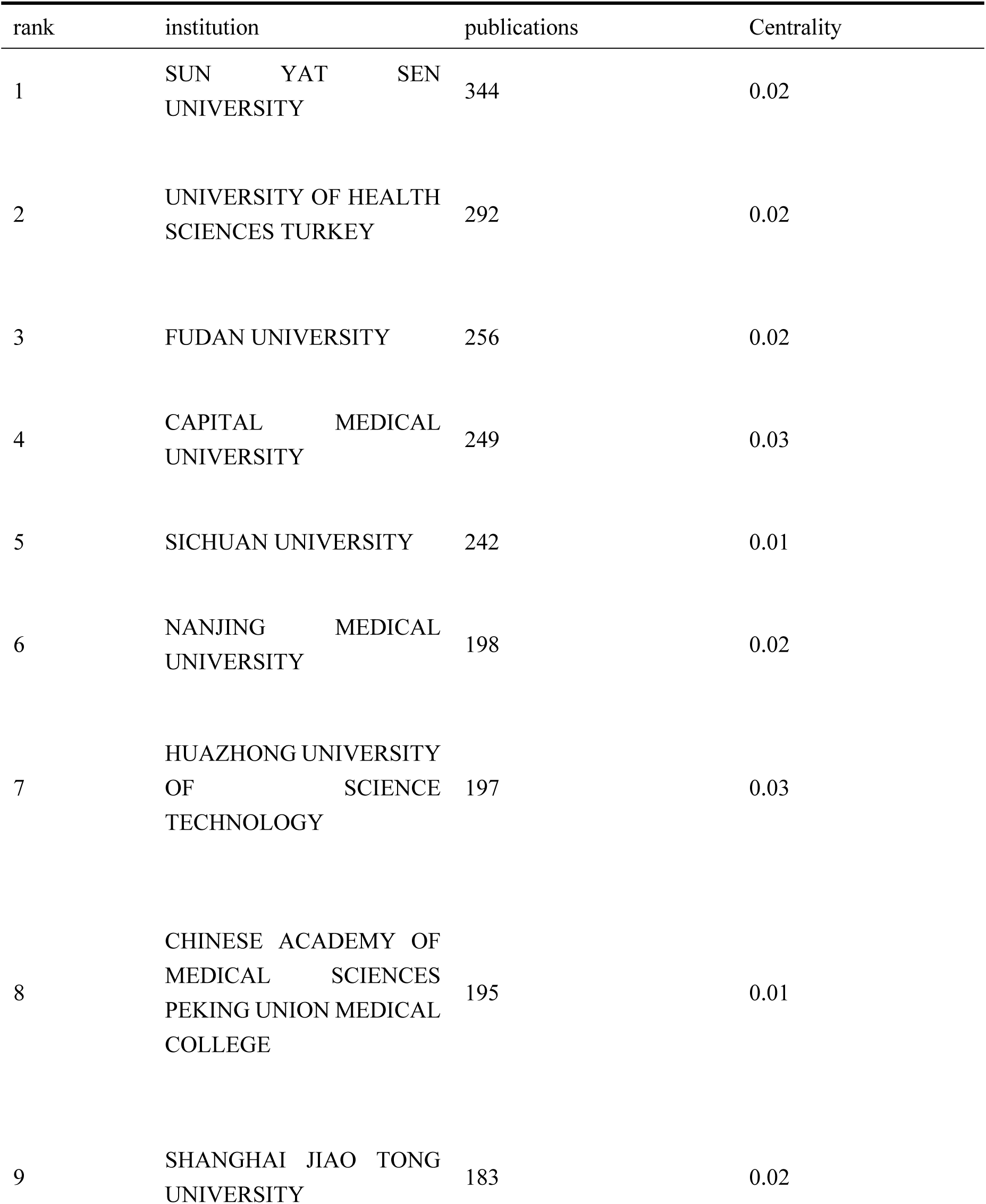
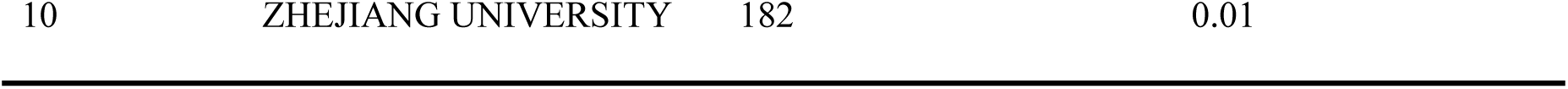
Top 10 institutions in terms of number of articles issued and the corresponding centrality.

Moreover, a comprehensive analysis of the data from table 2 reveals that nine of the top ten institutions in terms of the number of publications are from China. It suggests that in recent years, publications of NLR-related papers in China has been quite impressive in terms of quantity and importance. There is a potential to serve as a valuable source of reference and direction, by guiding the development of future international collaborations in this field. Figure 3A shows us the tens of institutions involved in the largest number of NLR-related publications in the world. The size of the radius of the node is positively correlated with the number of publications of the institution. And the institutions are distinguished by different colours depending on the direction of research focus. The number and thickness of the lines connecting the two nodes represent how closely the two institutions collaborate and communicate with each other in the same region or across regions. Figure 3B builds on Figure 3A with a higher level of visualisation and sophistication, using the colours of the nodes to show when the institution published NLR-related literature. Figure 3C shows us the 11 institutions in the world with the highest number of NLR-related publications in citespace. The larger radius of the node represents the higher number of papers published by that institution. The number and colour of the lines between the nodes represent the closeness of cooperation and communication between the two institutions and the corresponding time respectively. We could find that the University of Health Science Turkey is one of the most productive institution in the institutional cooperation network, but its centrality is low. In contrast, institutions such as Sichuan University, NanJing Medical University, Peking Union Medical College, and University of Texas System have higher centrality, indicating that they have extensive collaborations with academic institutions around the world. What’s more, it is easy to find that the cooperation and exchange between different institutions still focuses on Chinese universities. Figure 3D lists the 25 institutions with the highest citation frequency mutation index. The red line segments in the figure indicate the time periods when the citation frequency of the papers published by the institution increased significantly. Combining the temporal clues and the research focus of the institution, we can infer the hotspots and general direction of NLR-related research in academia during a certain period of time.

### Distribution of authors

This section is concerned with the analysis of published and cited authors of papers related to NLR. Collaboration between different authors is frequently utilised to analyse their collaboration patterns (in Figure 4A). The size of the nodes is typically associated with the number of publications or the frequency of collaboration of the corresponding authors. The utilisation of different colours for the nodes signifies the distinction between various collaboration groups or clusters. The thickness of the connecting lines between each author is indicative of the frequency of collaboration. The red nodes in the central region form a substantial cluster, indicating a close collaboration between these authors. The red clusters in the central region demonstrate a significant collaborative group. For instance, authors ZHANGWEI, WANGWEI and WANGJUN have a considerable influence in this subfield, while the smaller clusters in the periphery indicate minor collaborative relationships.

In a similar manner, Figure 4B was generated by the VOSviewer software in order to represent the distribution of cited authors (authors who published NLR-related literature which was cited by another author). The timeline colours in the figure range from blue to yellow in order to indicate the time span from 2016 to 2022.The alteration in colour serves to facilitate comprehension of the temporal trend of the collaboration. The dimensions of the nodes and the thickness of the connecting lines serve to indicate the intensity and frequency of the collaboration. Nodes of differing colours represent disparate groups of collaborations. Blue colouration is used to denote older collaborations (e.g. authors such as PICHLER.M and OYLUMLU.MUSTAFA), whereas yellow colouration is used to denote newer collaborations (e.g. authors such as KALLER.REKA). It is evident that there has been an increase in the number of collaborations between authors in recent years, as evidenced by the gradual yellowing of the colours (e.g. citation-citation relationships between ZINELLU.ANGELO, PIRINA.PIETRO and KALLER.REKA, JEMIELITY). The blue and green nodes in the centre area form a large cluster, indicating that these authors have been collaborating closely since earlier periods.

The term ‘co-cited authors’ is employed to denote the phenomenon in which the literature of two or more authors is cited by one or more subsequent papers at the same time. In such instances, these authors are said to constitute a co-citation relationship and are designated as co-cited authors (see Figure 4C).The utilisation of nodes of differing colours signifies the delineation of diverse clusters, with nodes of analogous colours denoting heightened collaborative relationships amongst these authors. These clusters are distributed across disparate regions, thereby establishing multiple autonomous collaboration networks. The red clusters are characterised by a greater density of nodes, signifying that these authors engage in frequent collaborative interactions with one another. As is evident, AZAB.B is the most influential co-cited author among the sample. In summary, the visual representation provides a clear illustration of the relationship between co-cited authors and citing authors of NLR-related literature.

### Distribution of journals

We used the bibliometric online analysis platform to identify journals with high publication volume and impact in NLR related fields. The journal’s impact factor (IF) and Journal Citation Reports (JCR) quartile are indicative of its influence. Journals with the top 25% (including 25%) of IF are in JCR quartile 1(Q1), and top 25%-50% (including 50%) of IF are in JCR quartile 2(Q2). Table 3 shows the top 10 journals in the number of articles, corresponding IF (JCR2023) and JCR quartile. The journal with the highest number of publications is MEDICINE (1.3, Q2) (311), followed by PLOS ONE(2.9, Q1) (280), FRONTIERS IN ONCOLOGY (3.5, Q2) (272), and SCIENTIFIC REPORTS (3.8, Q1) (252). Among the top ten journals in terms of the number of publications, five journals are distributed in the Q1 JCR. The most frequently co-cited journals are PLOS ONE (2.9, Q1) (8879) and NEW ENGLAND JOURNAL OF MEDICINE (96.3, Q1) (8651). Among the top 10 journals in co-citation frequency (in table 4), nine journals are distributed in Q1 JCR and five journals have an IF over 10. It is important to note that three of the top ten journals in terms of publication volume are also among the top ten journals in terms of co-citation frequency. These journals include PLOS ONE, SCIENTIFIC REPORTS and ONCOTARGET, which indicate a strong influence on the field.

**Table 3.** Top 10 journals in terms of number of publications, corresponding IF (JCR2023) and JCR quartile.

| rank | journal | publications | IF (JCR2023) | JCR quartile |
| --- | --- | --- | --- | --- |
| 1 | MEDICINE | 311 | 1.3 | Q2 |
| 2 | PLOS ONE | 280 | 2.9 | Q1 |
| 3 | FRONTIERS IN ONCOLOGY | 272 | 3.5 | Q2 |
| 4 | SCIENTIFIC REPORTS | 252 | 3.8 | Q1 |
| 5 | JOURNAL OF CLINICAL MEDICINE | 194 | 3.0 | Q1 |
| 6 | CANCERS | 169 | 4.5 | Q1 |
| 7 | BMC CANCER | 167 | 3.4 | Q2 |
| 8 | CANCER MANAGEMENT AND RESEARCH | 160 | 2.5 | Q3 |
| 9 | FRONTIERS IN IMMUNOLOGY | 157 | 5.7 | Q1 |
| 10 | ONCOTARGET | 126 | / | / |

**Table 4.** Top 10 journals in terms of number of citations, corresponding IF (JCR 2023) and JCR quartile.

| rank | journal | Citations | IF (JCR2023) | JCR quartile |
| --- | --- | --- | --- | --- |
| 1 | PLOS ONE | 8879 | 2.9 | Q1 |
| 2 | NEW ENGLAND JOURNAL OF MEDICINE | 8651 | 96.3 | Q1 |
| 3 | BRITISH JOURNAL OF CANCER | 7362 | 6.4 | Q1 |
| 4 | JOURNAL OF CLINICAL ONCOLOGY | 7064 | 42.1 | Q1 |
| 5 | LANCET | 6707 | 98.4 | Q1 |
| 6 | ANNALS OF SURGICAL ONCOLOGY | 5356 | 3.4 | Q1 |
| 7 | NATURE | 4881 | 50.5 | Q1 |
| 8 | SCIENTIFIC REPORTS | 4754 | 3.8 | Q1 |
| 9 | ONCOTARGET | 4207 | / | / |
| 10 | CLINICAL CANCER RESEARCH | 4083 | 10.4 | Q1 |

After that, we used bibliometrics online analysis software to identify journals with high publication volume and impact in areas related to NLR. As illustrated in Figure 5A, the utilisation of diverse colour represents different categories of publications. Each node, corresponding to a specific journal, is assigned a colour in accordance with the specified categorisation. The size of nodes are positively correlated with the frequency of occurrence of that journal. The classification system comprises six primary categories. The red categories represent research related to diseases of the endocrine, cardiovascular, and respiratory systems, as well as research related to pharmacology. Within the red categories, it is evident that PLOS ONE and JOURNAL OF CLINICAL MEDICINE occupy prominent positions within the high-impact bracket. The green ones pertain to the field of surgical oncology (WORLD JOURNALS OF SURGERY, ANNALS OF SURGICAL TREATMENT, etc). The nodes coloured blue are cancer related journals, where FRONTIERS IN ONCOLOGY and CANCERS occupying a predominant position. The yellow and purple represent neurology and gynaecology diseases (FRONTIERS IN NEUROLOGY, EUROPEAN JOURNAL OF, OBSTETRICS etc), respectively. And finally, the nodes in orange are considered to be journals related to head and face disorders, where the term laryngoscopy is mentioned several times.

Similar to Figure 5A, Figure5B describes the distribution of co-cited journals and the links between them. A higher number of co-citations suggests that a journal may be important in NLR-related research and a larger radius of the nodes indicates a higher co-citation frequency. What’s more, the density of links between them indicates the frequency of contact between different journals. It is noteworthy that PLOS ONE maintains its high impact status in this analysis, with JAMA-J AM MED ASSOC, CIRCULATION, and AMERICAL JOURNAL OF CARDIOLOGY exhibiting high co-citation frequencies in the red nodes, which represent cardiovascular-related journals. Conversely, cellular immunology is indicated in blue, with significant attention from JOURNAL OF IMMUNOLOGY and NATURE. And among the journals related to oncology (green nodes), BRITISH JOURNAL OF CANCER, JOURNAL OF CLINICAL ONCOLOGY, and ANNAL OF SURGICAL ONCOLOGY are of particular interest.

Figure5C and Figure5D focus on the temporal dimension that based on Figure5A and Figure5B. The colour of the lines corresponds to the time indicated in the legend, which highlights the time of occurrence of publications (Figure5C) and co-citations (Figure5D). As illustrated in Figure 5C, Early studies pertaining to the NLR have been published in journals such as international JOURNAL OF CLINICAL MEDICINE and ANNALS OF SURGICAL ONCOLOGY. Subsequent to 2020, the majority of studies have been published in CANCERS, FRONTIERS IN ONCOLOGY and FRONTIERS IN IMMUNOLOGY. Perusal of the aforementioned journals may allow the identification of research hotspots related to NLR. What’s more, as demonstrated in Figure D, the earlier cited literature pertaining to NLR was predominantly published in journals such as ONCOTARGET and TUMOR BIOLOGY, while the more recent literature has been published more frequently in CANCERS, FRONTIERS IN ONCOLOGY, FRONTIERS IN IMMUNOLOGY and other similar journals.

The local impact of journals is measured by the H index in Figure5E. The H index is a hybrid quantitative metric that assesses the amount of scholarly output versus the level of scholarly output by researchers. The journals under scrutiny in this particular analysis encompass PLOS ONE, BRITISH JOURNAL OF CANCER, and ONCOTARGET, amongst others. The H-index of these journals ranges from 22 to 49, with PLOS ONE exhibiting the highest H-index of 49, followed by BRITISH JOURNAL OF CANCER with an H-index of 47. Moreover, it is evident that prominent journals, including ONCOTARGET, ANNALS OF SURGICAL ONCOLOGY, SCIENTIFIC REPORTS, BMC CANCER and MEDICINE possess a significant impact in their respective domains, as evidenced by their H-indexes which exceed 30. The chart illustrates the impact of the journals within the academic sphere.

The application of knowledge flow analysis facilitates the exploration of the evolution between cited and citing journals.The overlay analysis of journal biplots (Figure 5F) demonstrates the thematic distribution of academic journals, alterations in citation trajectories, and shifts in research centres. As illustrated in the figure, the labels on the left side of the figure represent the journals that made the citation, while the labels on the right side represent the journals that were cited. The coloured curves in the middle originate from the journals that made the citation and point to the cited journals.The analysis revealed that the journals which were cited were predominantly in clinical pharmacology, molecular biology, immunology, biochemistry, and natural science journals, while the journals that were cited were predominantly in computer science, environmental science, toxicology, nutrition, molecular biology, genetics, nursing, psychology, ecology, and even journals in literary disciplines such as history and politics.

### Keyword analysis

As an overview of the core content of the article, the keywords can be utilised to analyze the frontiers and hotpots of network research in recent years. This may facilitate comprehension of the prevailing research direction concerning the “Netrophil to lymphocyte ratio” (NLR). The Table 5 demonstrates the top 20 keywords by Occurrences. The word which occured most frequently is “neutrophil to lymphocyte ratio” (5670), followed by “inflammation” (3762). In addition, “neutrophil” (3489), “survival” (3141) “prognosis”(2261) and “lymphocyte ratio”(2007) are frequent keywords, indicating that their corresponding fields are popular in NLR-related research.

**Table 5.**
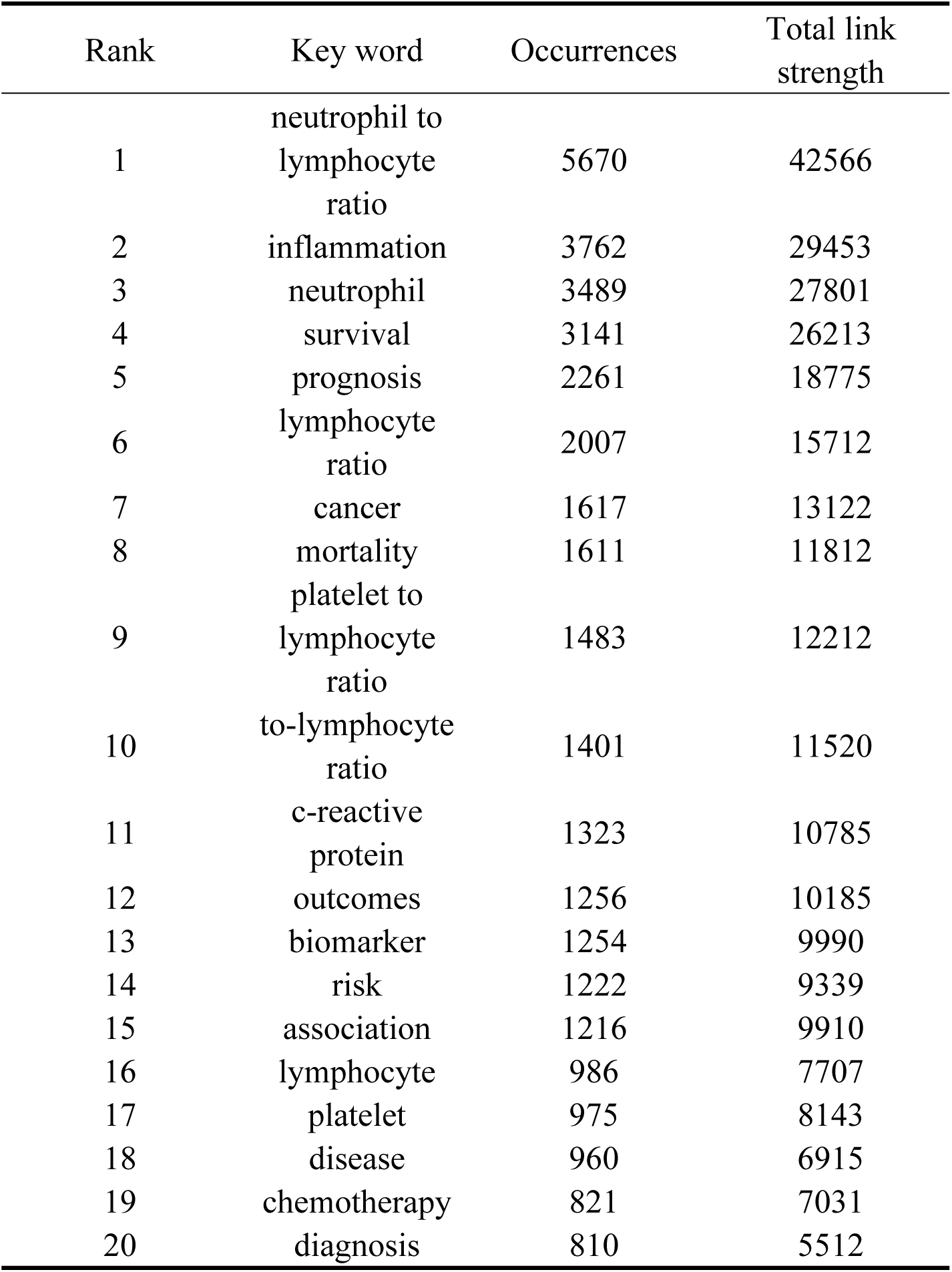
Top 20 keywords in terms of frequency of occurrence and the corresponding total link strength.

Figure 6A shows the strong citation bursts in the academic literature for 25 keywords between 2004 and 2024. The term ‘citation bursts’ is employed to denote the phenomenon of a sudden and significant increase in the number of citations of a particular keyword within a given time period. This occurrence is widely regarded as an indication that the subject in question has garnered a notable degree of attention from the academic community during that particular time frame.Among them, the term ‘year’ is used to denote the year in which the citation surge commenced, while ‘strength’ is used to indicate the strength of the citation surge. It is important to note that a larger value indicates a greater degree of attention received by the keyword during that specific time period. The terms ‘begin’ and ‘end’ denote the year in which the citation surge began and ended, respectively. As demonstrated in Figure 6A, the term “treatment neutrophil” exhibited the most significant citation surge intensity (54.84) in 2013, with this trend persisting until 2018. Notably, the term “long term mortality” emerged in 2008 and remained among the most persistent surges until 2016. The terms “coronavirus disease 2019” and “pneumonia” began to show strong citation surges in 2020, coinciding with the timing of the global outbreak of the disease.As is apparent from the results, earlier studies concentrated more on the relationship between the neutrophil-to-lymphocyte ratio (NLR) and immune cells, immunopathological responses, and the predictive role of the NLR in disease survival.After 2015, more research has focused on the relationship between NLR and various diseases, such as atherosclerosis, arterial disease.In recent years, the relationship between NLR and systemic immune indices and immune checkpoints has become a new research hotspot.

Figure 6B and Figure 6C are both keyword co-occurrence network diagrams of NLR-related literature, which was drawn by VOSviewer. This figure is employed to illustrate the prevalence and trends of research subjects within a given field. Each circle in the figure corresponds to a keyword, with the size of the circle denoting the frequency of the keyword’s appearance in the literature. The connecting line between the circles signifies the co-occurrence relationship between the keywords. For instance, the frequency with which they are documented in the same literary sources can be measured. Figure 6B employs a colour coding system to represent different time periods, with the data set spanning from 2018 to 2021. Older studies predominantly focus on the darker regions of the figure, such as the purple and blue areas, while newer studies concentrate on the lighter regions, such as the green and yellow areas. A cursory investigation reveals that the keyword “neutrophil to lymphocyte ratio” is the largest node, indicating that this is a very important research topic in this field. An examination of the keywords reveals many related topics, such as “inflammation”, “mortality”, “survival” and “cancer”. As illustrated in Figure 6C, the utilisation of diverse coloured nodes may signify the representation of varied temporal periods or subject categories. As illustrated in Figure, it is evident that certain keywords are grouped in a discernible manner, thereby delineating distinct clusters. These clusters, in turn, serve as representations of specific research topics or domains of paramount relevance. In the figure, The keywords are clustered according to the research direction and roughly divided into 4 categories: keywords in the blue cluster are related to diseases associated with NLR and various immune markers linked to NLR (anemia, insulin resistance, c-reactive protein, neutrophil count etc.). The keywords in the green cluster are related to the relationship between NLR and various malignant tumours, including lung cancer, breast cancer and colon cancer (circulating tumor-cells, liver metastasis, overall survival, etc.). Keywords in the red cluster are related to relationship between NLR and multiple diseases of different organ and system (covid-19, coronavirus disease, etc.) and immune cells, immune markers and immunopathological responses (leukocytes, macrophages, pathway, etc.). The keywords in the purple cluster are related to predictive role of NLR in disease, especially tumours (prediction model, prognostic indicator, etc.). The keywords in the yellow cluster (platelet count, predicts prognosis, etc.), the light blue cluster (adverse outcomes, disease activity, etc.), and the orange cluster (thrombosis, inflammatory etc.) are linked to several clusters, indicating that they are cross-cutting areas in each research direction.

Furthermore, the graph elucidates the existence of associations between disparate subject areas. To illustrate this point, the graph may, for instance, concurrently encompass keywords pertinent to multiple domains, including but not limited to cardiovascular disease, cancer, and immunology. This phenomenon signifies the convergence and interplay between these diverse fields of study. It is evident that the two diagrams provide a valuable insight into the research hotspots, development trends and correlations between different research topics in NLR in terms of time and space. This is of great significance for researchers, as it enables them to plan their research directions, understand the research dynamics and discover potential collaboration opportunities.

Figure 6D is a keyword co-occurrence network graph generated by CiteSpace, which demonstrates the evolution and trends of various research topics between 2004 and 2024. The size of a node is indicative of the frequency with which a keyword appears in the literature, while the connecting line between nodes signifies the co-occurrence relationship between keywords. The horizontal axis denotes the year of first appearance of a keyword in the NLR-related literature, with the vertical axis listing the top ten subjects in chronological order. In this figure, the earliest and largest cluster is #0 (prognosis). In this field, the earliest keywords include netrophil to lymphocyte ratio (NLR) and prognosis, while progonostic model, inflammatory burden index, age related macular degeneration are the latest research targets. Another large cluster that emerged earlier is #1 (cornary artery disease). In this field, keywords such as pan-immune-inflammation value and infract-related artery are the research frontiers. It is worth noting that 3 of the 10 clusters are still in progress, namely, #0(prognosis), #1 (cornary artery disease) and #7 (covid-19), indicating that relevant research in this field is ongoing. In addition, #9 (acute ischemic stroke) is the latest cluster; the main keywords are acute ischemic stroke, intravenous thrombolysis and mechanical thrombectomy, which is a frontier hotspot of NLR-related research.In summary, the closer a keyword is to the top right corner of the graph, the more recent and popular it is.

Figure 6E is a chord diagram which shows the keywords’ annual popularity (number of citations in the year/total citations in the year) from 2004 to 2024. As illustrated in the diagram, the circle in the centre is divided into sectors, with each sector representing a distinct category or group of keywords. As evident in the figure, keywords such as diagnosis and biomarkers have had relatively high annual popularity in recent years, which suggests that these keywords represent emerging frontier areas. In contrast, the annual popularity of keywords such as resection, pretreatment neutrophil, colorectal cancer and carcinoma have been relatively much lower in recent years. Nevertheless, the prevailing tendency indicates an augmentation in the heat level of all keywords in recent years. This finding suggests a gradual escalation in the significance of NLR-related research and a concomitant rise in its importance.

### Subject area analysis

This figure is a highly cited reference. The highly cited reference analysis is typically employed to represent a research topic that is currently receiving significant attention or a discovery of considerable importance within a specific field. The analysis of such literature can facilitate the rapid comprehension of current research priorities.By engaging with and analysing highly cited literature, researchers can acquire knowledge regarding high-quality research design, methodology and research style, thereby enhancing the quality of their own research endeavours.

Table 6 shows the top ten articles in terms of citation frequency and average annual citation frequency. The most frequently cited article is “Prognostic role of Neutrophil to lymphocyte ratio in solid tumors: a systematic review and meta-analysis”, whose author is Arnoud J Templeton et al, published in 2004 (522). This research focused on predictive role of high NLR in poor prognosis of various solid tumours. Next, “The systemic inflammation-based neutrophil-lymphocyte ratio: experience in patients with cancer” (Graeme J K Guthrie et al. 2013) (302). This systematic review highlights the prognostic value of the Neutrophil to lymphocyte ratio (NLR) across various cancer types and stages, demonstrating its potential clinical utility in reflecting tumor aggressiveness and systemic inflammation. The following one is Neutrophil to Lymphocyte ratio (NLR) and Platelet-to-Lymphocyte ratio (PLR) as prognostic markers in patients with non-small cell lung cancer (NSCLC) treated with nivolumab. This study found that the Neutrophil to lymphocyte ratio (NLR) and platelet-to-lymphocyte ratio (PLR) are independent prognostic markers in NSCLC patients treated with nivolumab, with higher NLR and PLR significantly associated with shorter overall survival and lower response rates.

**Table 6.**
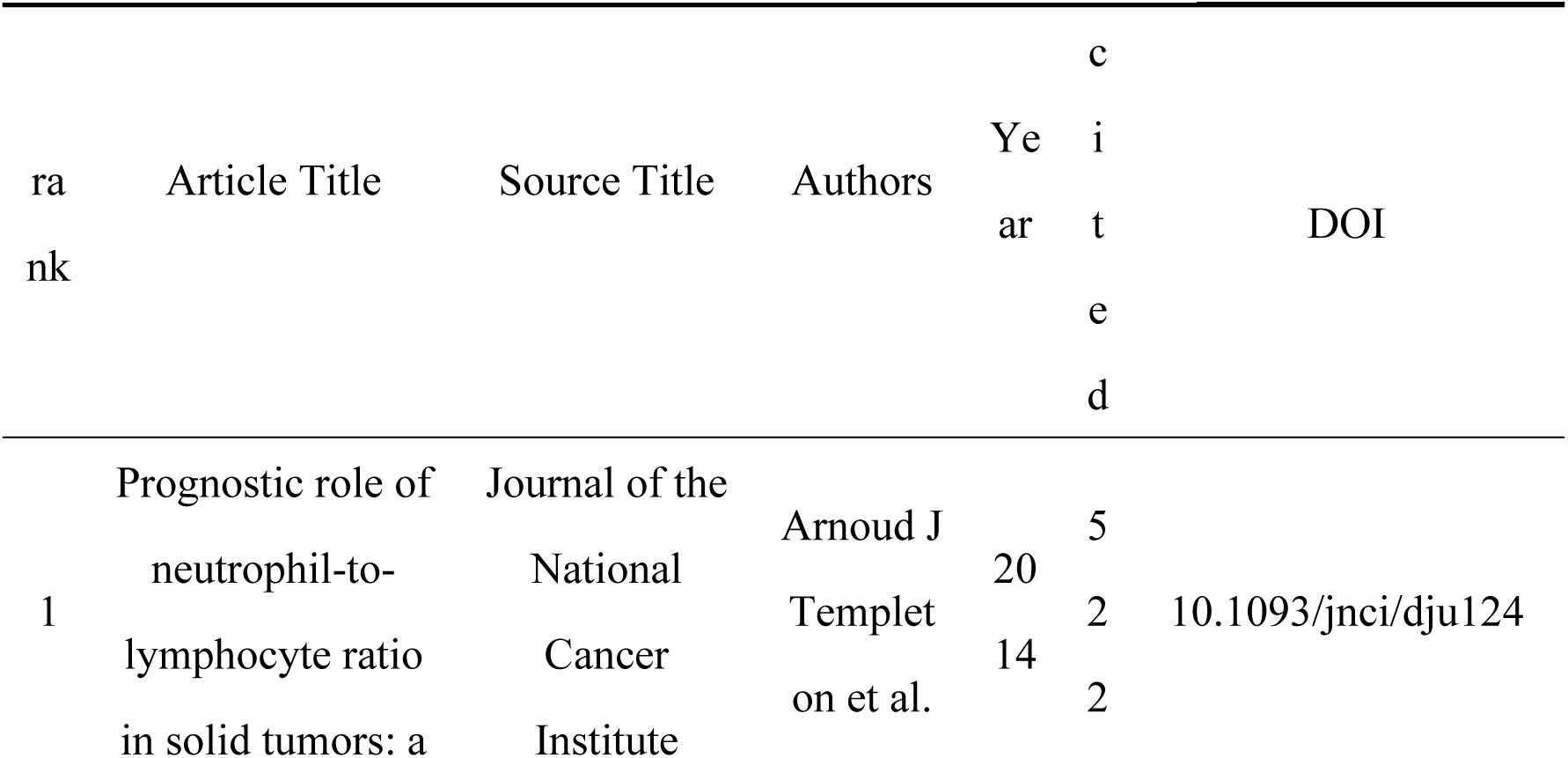
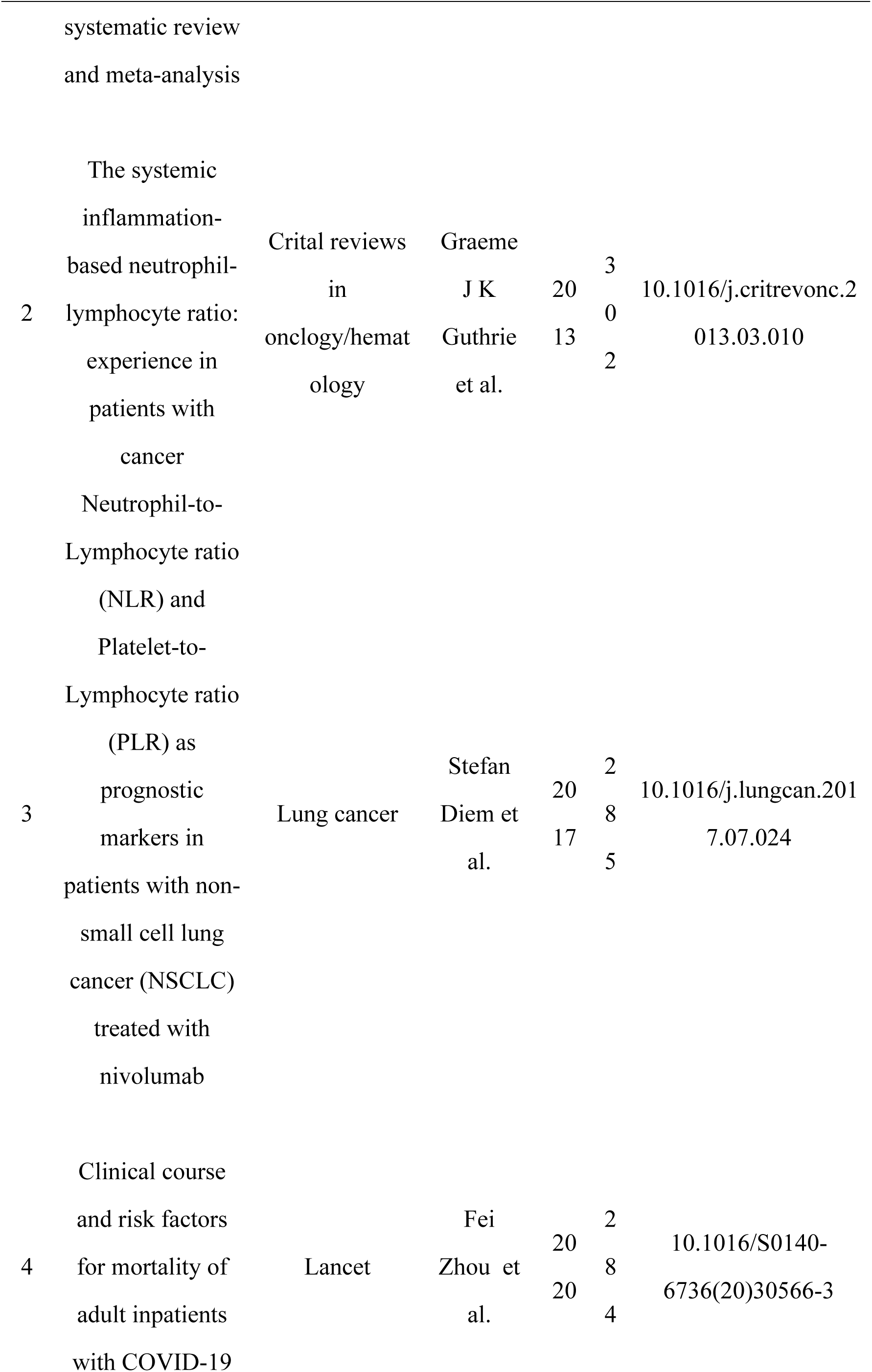
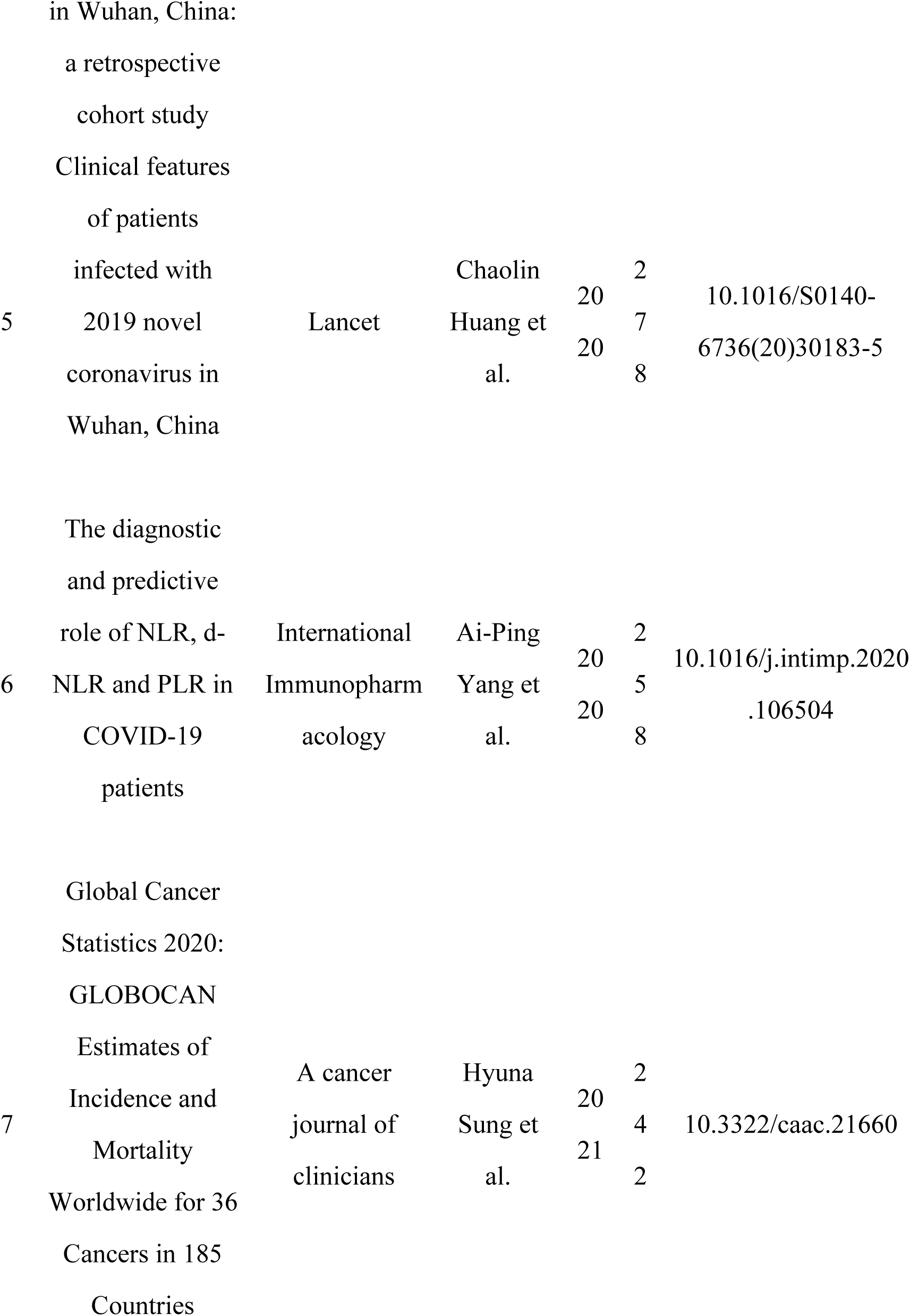
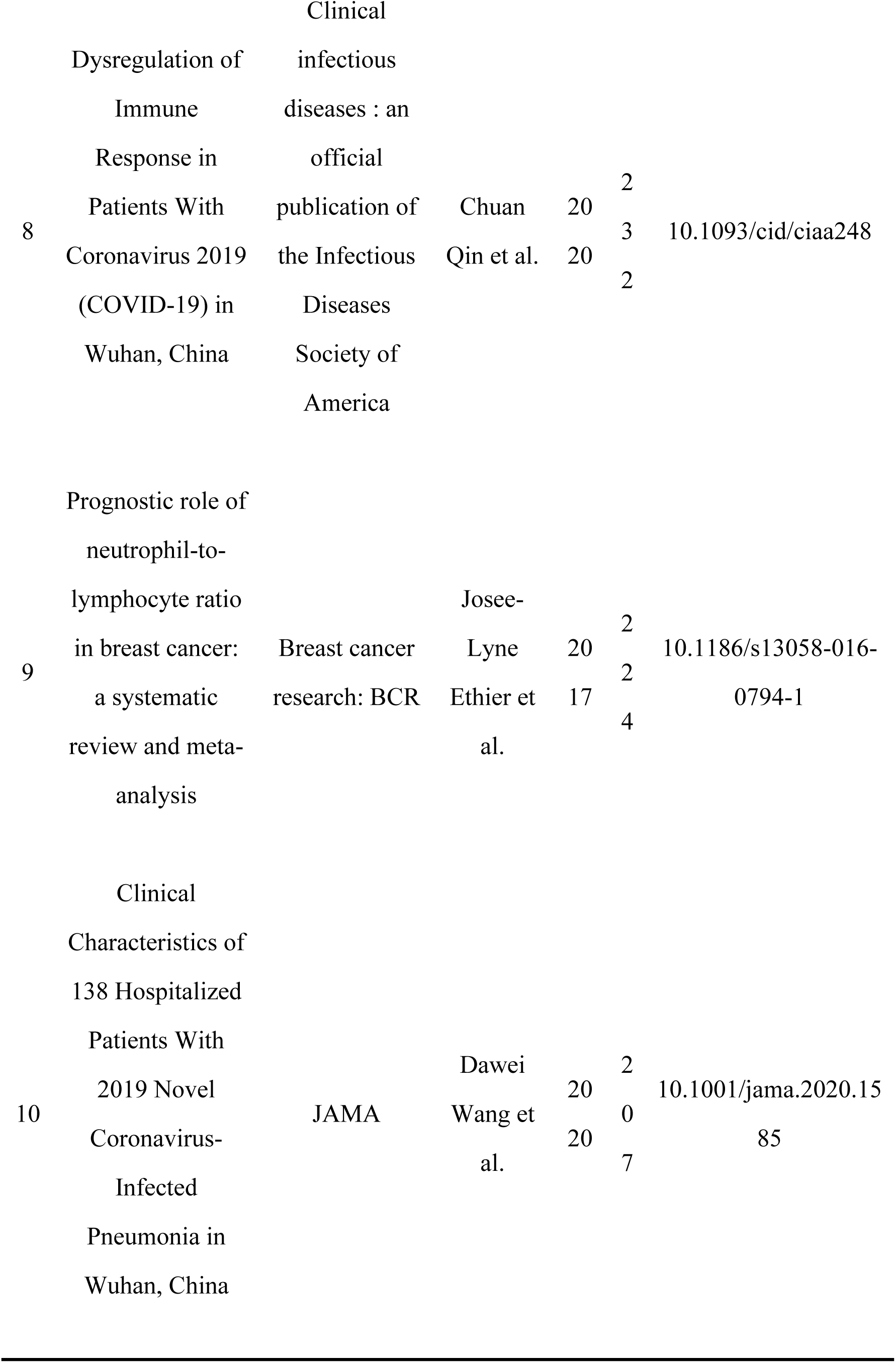
Top 10 highly cited references.

As illustrated in Figure 7A, the 25 references exhibiting the most significant citation burst intensity are identified, accompanied by the author’s name, the year of publication, the journal title, the volume number, and the page number. The right-hand side of the table features a timeline graph, which illustrates the time period of citation outbursts for each reference. The red bar in the graph indicates the time period of citation outbursts, which, as is evident from the timeline graph, are concentrated between 2010 and 2018 for the majority of references. The first citation burst occured in 2009, with the title “Pre-treatment neutrophil to lymphocyte ratio is elevated in epithelial ovarian cancer and predicts survival after treatment”. The highest citation burst intensity is observed in Prognostic role of Neutrophil to lymphocyte ratio in solid tumors: a systematic review and meta-analysis(Arnoud J Templeton et al), with an intensity of 174.35, and an outbreak period from 2016 to 2019. The results show that 2014 and 2020 had the highest number of new citation bursts (4 times), followed by 2013 and 2016 (3 times), which indicates that the high-burst papers in these two

Years caused a related research boom. What’s more, as is evident from the results, the majority of these co-cited highlighted articles are tumour-related and primarily explore the predictive role of the Neutrophil to lymphocyte ratio (NLR) in the occurrence and prognosis of various solid tumours. However, articles published between 2019 and 2021 primarily focus on covid-19 pneumonia, with a significant proportion of these exploring the predictive role of NLR in determining the prognosis of the disease.

Figure 7B was generated by CiteSpace and illustrates the citation relationships and knowledge structure between disparate literatures. Each node in the figure represents a piece of literature, and the nodes are labelled with the author and the year of publication. The lines connecting the nodes represent the citation relationship, with the thickness and colour of the edges indicating the strength or duration of the citation. For instance, the substantial size of the nodes representing ‘Hanahan D (2011)’ and ‘Chua W (2011)’ in the graph suggests that these publications have a significant impact on the field. The colour distribution reveals the presence of both older (blue) and newer (red) literature in the graph, thus indicating the research history and development of the field. By conducting a thorough analysis of the clusters and significant literature, it is possible to identify the research hotspots and cutting-edge issues within the field.

Figure 7C provides an analysis of the top ten cluster labels, with one label assigned to each cluster. The labels are listed in descending order of citation frequency. There are 10 labels in total: #0_to_lymphocyte_ratio, #1(long-term mortality), #2(poor prognosis), #3(delta neutrophil index), #4(laryngeal squamous cell carcinoma), #5(systemic immune-inflammation index), #6(hepatocellular carcinoma), #7 (checkpoint inhibitors), #8(prognostic factor), #9(renal cell carcinoma). The figure reveals the research hotspots, trends, and knowledge structure of NLR, which helps researchers understand the dynamics and impact of research in a particular field. As illustrated in the figure, the earliest research areas in the NLR field are two separate research clusters: #7 (checkpoint inhibitors) (#8 (prognostic factor), followed by #6(hepatocellular carcinoma). Furthermore, recent studies have concentrated on the following areas: #2(poor prognosis), #3(delta neutrophil index), #4(laryngeal squamous cell carcinoma), #5(systemic immune-inflammation index).

As illustrated in Figure 7D, a bar chart is presented that enumerates the most frequently cited publications within the domain of NLR on a global scale.The Y-axis of the chart provides the citation information of the publications, including the names of the authors, the years of publication and the names of the journals. The X-axis indicates the number of citations received by each publication, with a range of values from 0 to more than 3,000. Each data point corresponds to a specific piece of literature, with its position denoting the global citation ranking of that literature. The literature with the highest number of citations is Dysregulation of Immune Response in Patients With Coronavirus 2019 (COVID-19) in Wuhan, China (Chuan Qin et al., 2020), with 3,398 citations, followed by Prognostic role of neutrophil-to-lymphocyte ratio in solid tumors: a systematic review and meta-analysis (Stefan Diem et al., 2014), with 2,094 citations. The majority of the literature had an average citation count of less than 100, with a smaller subset having an average citation count of more than 1000. The time frame of publication of the literature extends from 2005 to 2021, thereby demonstrating the impact of these publications across different periods.

As illustrated in Figure 7E, a bar chart is employed to display the most frequently cited literature within a specific region or database.The Y-axis of the chart enumerates the citation information of the literature, including the author, year of publication, and journal name. The X-axis indicates the number of local citations received by each piece of literature, with a range of values from 0 to over 600. The document that has received the highest number of citations is Neutrophil-lymphocyte ratio as a prognostic factor in colorectal cancer (S R Walsh et al., 2005), with 671 citations, followed by Association between admission neutrophil to lymphocyte ratio and outcomes in patients with acute coronary syndrome (Umesh U Tamhane et al., 2008), with 528 citations. This graph assists researchers in comprehending the literature with the highest impact in the region or database. Through analysis of this literature, it becomes possible to identify significant research results and research hotspots in various subfields of NLR.

Figure 7F was generated by CiteSpace, which demonstrates the co-citation relationship between documents from disparate years. Each node in the network represents a piece of literature, and the nodes are labelled with the author and the year of publication. The lines between the nodes indicate the co-citation relationship between the documents. The colour of a node serves to indicate the year of publication of the literature, whilst the size of the node is indicative of the number of citations received by the literature. The presence of clusters of nodes of different colours signifies the existence of distinct clusters. The larger size of some of the nodes indicates that they have been cited more frequently and are of significant importance within the field. For instance, the result shows Neutrophil-lymphocyte ratio as a prognostic factor in colorectal cancer, published by S R Walsh and Association between admission neutrophil to lymphocyte ratio and outcomes in patients with acute coronary syndrome, published by Stefan Diem are notably larger, suggesting that these documents have a considerable impact on the filed.

A highly cited literature analysis is an important tool in academic research and knowledge management, which can help stakeholders from all sides to better understand and utilise the results of academic research.

### Subject area analysis

We used CiteSpace to analysis the citation relationship of NLR-related literature by subject area (Figure 8). It is evident that the field of “ONCOLOGY” has received the highest number of citations compared with other fields related to the NLR. Furthermore, the fields of “MEDICINE, RESEARCH & EXPERINMENT”, “IMMUNOLOGY”, “SURGERY”, “MEDICINE, GENERAL & INTERNAL”, “CLINICAL NUEROLOGY”, “NEUROSCIENCES”, “GASTROENTEROLOGY & HEPATOLOGY”, “PHARMACOLOGY & PHARMACY”, “MULTIDISCIPLINARY SCIENCES” and “BIOCHEMISTRY & MOLECULAR BIOLOGY” also play a pivotal role in the realm of NLR-related research. Last but not least, “ONCOLOGY”, “MEDICINE, RESEARCH & EXPERINMENT”, “IMMUNOLOGY”, “PHARMACOLOGY & PHARMACY”, “BIOCHEMISTRY & MOLECULAR BIOLOGY” are marked by purple circles, indicating that these disciplines have a greater influence in this field. The influence of these disciplines in this field is indicated by the purple circles around these disciplines.

## Discussion

CiteSpace, VOSviewer, and R-bibliometrix were employed to analyze 14,877 NLR-related articles from the Web of Science Core Collection (2004–2023), evaluating the spatiotemporal distribution, author contributions, core journals, research hotspots, and frontiers in this field.

### General Distribution

The analysis revealed an exponential growth in NLR-related publications over the past two decades, with China dominating both publication volume (4,959 articles) and citation frequency (97,887 citations). This reflects China’s substantial investment in NLR research, particularly in oncology and cardiovascular diseases. While Turkey and the United States ranked second and third in productivity, their centrality scores (0.06 and 0.14, respectively) were overshadowed by the United Kingdom (centrality: 0.37) and Germany (centrality: 0.36), indicating their pivotal roles as collaborative hubs. Notably, nine of the top ten institutions by publication output were Chinese universities, with Sun Yat-sen University leading (344 publications). However, institutions like Capital Medical University and Huazhong University of Science and Technology exhibited higher centrality (0.03), suggesting their influence in bridging interdisciplinary collaborations.

### Hotspots and Frontiers

Keyword and co-citation analyses identified NLR as a versatile biomarker across diverse medical fields. The timeline graph revealed three major clusters: #1 (cancer prognosis), #2 (cardiovascular diseases), and #3 (COVID-19). The surge in NLR-related COVID-19 studies post-2020 highlights its emerging role in predicting disease severity and mortality. Journals such as Frontiers in Immunology (IF: 8.786) and PLOS ONE (IF: 3.752) emerged as key platforms, while CA-CANCER J CLIN and LANCET dominated co-citation networks, underscoring NLR’s relevance in high-impact clinical research.

### NLR and Cancer

The Neutrophil to lymphocyte ratio (NLR) has become a widely recognised and versatile biomarker in oncology, with mounting evidence supporting its significance in cancer prognosis[24,25], response to therapy, and disease monitoring[12,17]. Elevated Neutrophil to lymphocyte ratio (NLR), a readily obtainable indicator, reflects a systemic inflammatory response that may facilitate tumour progression[26] and suppress anti-tumour immunity[6].

Recent studies have demonstrated the prognostic importance of the Neutrophil to lymphocyte ratio (NLR) in a broad range of malignancies. High Neutrophil to lymphocyte ratio (NLR) has been shown to be an independent predictor of poorer overall survival (OS), reduced disease-free survival[27], and more aggressive tumour characteristics in cancers such as colorectal, lung, breast, and gastric cancer[28,29]. It is noteworthy that elevated NLR is a significant predictor of poor cancer-specific survival (hazard ratio [HR] = 3.51) and disease-free survival (HR = 2.88) in rare tumours, such as penile squamous cell carcinoma (PSCC) and testicular cancer (TCa), irrespective of tumour stage[30,31]. These findings emphasise the broad applicability of NLR as a prognostic indicator across common and rare cancers[32].

The utilisation of NLR extends beyond the realm of prognosis; it is increasingly investigated as a predictive biomarker for cancer therapy response, particularly in the context of immunotherapy[33,34]. Emerging evidence suggests that a high Neutrophil to lymphocyte ratio (NLR) may serve as a prognostic indicator, indicating patients who are more likely to benefit from immune checkpoint inhibitors (ICIs)[34]. This association is supported by findings that a high NLR is associated with reduced efficacy and shorter progression-free survival in patients receiving ICIs[35]. This phenomenon may be ascribed to the presence of an immunosuppressive tumour microenvironment, characterised by elevated neutrophil and diminished lymphocyte counts.

Furthermore, there is an increasing interest in the predictive ability of NLR for rare tumours (e.g. neuroendocrine neoplasms), and ongoing research aims to validate its universality[36]. Large-scale analyses and cross-sectional studies, such as those utilising the NHANES dataset, further substantiate the correlation between elevated NLR and augmented cancer risk, even in the general adult population.

The appeal of NLR lies in its accessibility, low cost, and prognostic utility, making it an attractive tool in cancer clinical practice. As research progresses, the combined use of NLR with other biomarkers has the potential to enhance the accuracy of cancer risk assessment, patient stratification, and individualised treatment planning[37].

### NLR and Cardiovascular Diseases

The Neutrophil to lymphocyte ratio (NLR) has emerged as a significant biomarker for systemic inflammation and has garnered considerable attention in the field of cardiovascular diseases (CVD)[38]. Numerous studies have demonstrated that an elevated Neutrophil to lymphocyte ratio (NLR) is closely related to increased risk, severity, and poor prognosis in various cardiovascular conditions, such as coronary artery disease, acute myocardial infarction, heart failure, atrial fibrillation, and peripheral artery disease[38–40].

The Neutrophil to lymphocyte ratio (NLR) is a simple and cost-effective parameter derived from routine blood tests. It reflects the balance between pro-inflammatory neutrophils and regulatory lymphocytes[41]. An increase in the Neutrophil to lymphocyte ratio (NLR) has been demonstrated to be a valuable predictor of enhanced inflammatory response and immune system imbalance. These factors have been identified as crucial contributors to the initiation and progression of atherosclerosis and other cardiovascular events[42,43]. Numerous large-scale observational studies have demonstrated that patients with elevated NLR values are more prone to major adverse cardiovascular events, encompassing re-infarction, heart failure, and cardiac death[38,39,44].

For instance, in patients undergoing percutaneous coronary intervention (PCI), a high postoperative NLR has been demonstrated to be associated with both short-term and long-term adverse cardiovascular outcomes[45]. Conversely, in the context of heart failure and hypertension, elevated NLR has been shown to independently predict increased mortality and hospitalisation[46]. In patients with diabetes and prediabetes, a high Neutrophil to lymphocyte ratio (NLR) has been identified as an independent predictor of all-cause and cardiovascular mortality[47]. This finding serves to further illustrate the value of NLR in risk stratification across different high-risk groups.

Due to its accessibility and predictive power, NLR is gaining traction not only as a tool for risk assessment and prognosis but also as a potential target for further research to guide clinical decision-making in cardiovascular medicine[41,48]. Further research is needed to elucidate the combination of NLR with other inflammatory biomarkers and its role in optimising prevention and treatment strategies for cardiovascular diseases[42,49].

### NLR in Sepsis, SIRS, and MODS

The Neutrophil to lymphocyte ratio (NLR) has emerged as a critical biomarker in systemic inflammatory conditions, particularly sepsis, systemic inflammatory response syndrome (SIRS), and multiple organ dysfunction syndrome (MODS)[50]. Sepsis is a condition characterised by a dysregulated host response to infection. It is closely linked to Systemic Inflammatory Response Syndrome (SIRS), and often progresses to Multiple Organ Dysfunction Syndrome (MODS), a life-threatening complication involving the failure of two or more organ systems. NLR is considered a valuable tool in these processes due to its dual reflection of innate (neutrophils) and adaptive (lymphocytes) immune responses[27].

Researchers have hypothesised that the Neutrophil to lymphocyte ratio (NLR) could play a role in the diagnosis and prognosis of sepsis and systemic inflammatory response syndrome (SIRS)[50]. Elevated Neutrophil to lymphocyte ratio (NLR) levels have been demonstrated to be strongly associated with the severity of sepsis and systemic inflammatory response syndrome (SIRS). Neutrophilia and lymphopenia, driven by cytokine storms (e.g., IL-6, TNF-α), are hallmarks of early sepsis[51]. Research has demonstrated that an NLR >10 has a high degree of sensitivity (85%) and specificity (78%) in predicting septic shock[52,53]. Furthermore, in SIRS, an NLR ratio greater than 7 has been shown to be a significant predictor of higher Sequential Organ Failure Assessment (SOFA) scores and mortality rates[54,55]. It is noteworthy that NLR has been shown to outperform conventional markers such as C-reactive protein (CRP) and procalcitonin (PCT) in the differential diagnosis of sepsis from non-infectious SIRS[56,57].

Furthermore, NLR has been demonstrated to possess the capacity to function as a predictor of MODS. The term MODS is used to denote the terminal stage of uncontrolled inflammation[58]. NLR dynamics have been shown to reflect the transition from hyperinflammation to immunosuppression in sepsis[59]. A sustained Neutrophil to lymphocyte ratio (NLR) elevation (>12) beyond 72 hours post-admission has been demonstrated to be associated with the development of multi-organ dysfunction (MODS)[50,60]. Lymphopenia, a fundamental component of the Neutrophil to lymphocyte ratio (NLR), is indicative of immunosuppression and has been demonstrated to correlate with secondary infections and organ failure. For instance, in septic patients, an NLR >15 has been shown to predict acute kidney injury (AKI) and acute respiratory distress syndrome (ARDS) with odds ratios of 3.2 and 4.1[46,61], respectively.

As investigation into the mechanism, NLR’s prognostic value in sepsis lies in its ability to mirror the balance between pro-inflammatory neutrophil activity and lymphocyte-mediated immune regulation. Neutrophils release reactive oxygen species (ROS) and extracellular traps (NETs), exacerbating endothelial damage and organ injury. Concurrently, lymphopenia reflects apoptosis of CD4+ T cells and dendritic cells, impairing pathogen clearance[8]. This imbalance perpetuates tissue hypoxia and mitochondrial dysfunction, driving MODS[62,63].

Dynamic Neutrophil to lymphocyte ratio (NLR) monitoring has been demonstrated to facilitate risk stratification and therapeutic decision-making[50]. For instance, early NLR-guided immunotherapy (e.g., IL-7 administration to reverse lymphopenia) is a subject under investigation. However, heterogeneity in cutoff values and timing of measurements poses challenges to standardisation. It is recommended that future studies explore the integration of NLR into machine learning models that combine lactate, SOFA scores and genomic data, with a view to enhancing predictive accuracy[64].

To summarise, the Neutrophil to lymphocyte ratio (NLR) is a simple, economical and effective biomarker with a significant role in the diagnosis, assessment and prognosis of sepsis[50]. The exploration of the relationship between the NLR and sepsis has been demonstrated to facilitate a more profound comprehension of the underlying pathogenesis of sepsis, thereby contributing to the refinement of clinical strategies. It is recommended that future research examine the potential of combining NLR with other biomarkers, with a view to enhancing the accuracy of diagnosis and prognosis in cases of sepsis.

### NLR in Diabetes Mellitus: From Pathogenesis to Complications

The Neutrophil to lymphocyte ratio (NLR) has gained attention as a biomarker reflecting chronic inflammation and immune dysregulation in diabetes mellitus (DM), encompassing type 1 (T1DM), type 2 (T2DM), gestational diabetes (GDM), and monogenic forms such as maturity-onset diabetes of the young (MODY)[65]. In T1DM, elevated NLR is associated with autoimmune-driven β-cell destruction. The exacerbation of islet inflammation is attributable to neutrophil activation (via IL-17/Th17 pathways) and reduced regulatory T lymphocytes (Tregs). Research findings have indicated that a Neutrophil to lymphocyte ratio (NLR) greater than 3.5 is present in patients diagnosed with type 1 diabetes mellitus (T1DM) for the first time. This has been shown to be associated with elevated glycated hemoglobin (HbA1c) levels and a greater loss of residual β-cell function[66]. Furthermore, elevated NLR has been demonstrated to be a valuable predictor of rapid progression to diabetic ketoacidosis (DKA), with a hazard ratio (HR) of 2.1 for NLR >4[67].

In type 2 diabetes mellitus (T2DM), Neutrophil to lymphocyte ratio (NLR) has been identified as a surrogate marker for subclinical inflammation. Type 2 Diabetes Mellitus (T2DM) has been defined as a chronic inflammatory and metabolic syndrome. Meta-analyses indicate that a Neutrophil to lymphocyte ratio (NLR) of 2.2 or greater is associated with a 1.8-fold increased risk of macrovascular complications, such as coronary artery disease, and microvascular outcomes, including nephropathy[46,68]. Mechanistically, adipose tissue-derived cytokines (e.g., IL-6, TNF-α) have been demonstrated to drive neutrophilia and lymphopenia, thereby perpetuating insulin resistance[69]. It is noteworthy that a reduction in Neutrophil to lymphocyte ratio (NLR) through lifestyle modifications, such as the adoption of a Mediterranean diet, has been observed to be associated with enhanced glycemic control, as evidenced by a decrease in glycated hemoglobin (HbA1c) levels of up to 0.5% (p < 0.01)[70].

In GDM, an NLR >3.0 during the second trimester has been shown to predict insulin resistance (HOMA-IR >2.5) and adverse pregnancy outcomes, including preterm birth (OR=2.3) and neonatal hypoglycaemia (OR=1.9). It is evident that placental hypoxia and neutrophil infiltration (via NETosis) result in impaired trophoblast function, while lymphopenia is indicative of maternal immunosuppression. Postpartum, the sustained elevation of the neutrophil-to-lymphocyte ratio (NLR) (>2.8) has been demonstrated to indicate a fourfold higher risk of progressing to type 2 diabetes mellitus (T2DM) within a five-year period.

In MODY and other monogenic diabetes, NLR can be regarded as a unique immunological profile. The extant data set is limited, but it suggests that the Neutrophil to lymphocyte ratio (NLR) patterns associated with monogenic diabetes mellitus (MODY) differ from those associated with polygenic diabetes mellitus (DM)[71,72] . For instance, patients diagnosed with HNF1A-MODY exhibit a lower median NLR (1.6 vs. 2.4 in T2DM) which may be attributable to preserved β-cell function and reduced systemic inflammation[71]. However, GCK-MODY demonstrates no significant deviation in NLR, thus highlighting subtype-specific heterogeneity. Consequently, there is a necessity for further study to be conducted on the relationship between these diseases and NLR.

In conclusion, the cost-effectiveness and accessibility of NLR renders it a promising instrument for diabetes stratification[46,65]. The following potential applications are suggested: firstly, the integration of the Neutrophil to lymphocyte ratio (NLR) with genetic markers (e.g. TCF7L2 variants) to refine the diagnosis of maturity-onset diabetes of the young (MODY) is suggested[72]. Furthermore, in the domain of therapeutic monitoring, NLR-guided anti-inflammatory therapies (e.g., SGLT2 inhibitors) have been employed in the management of T2DM[73]. Finally, in the field of precision medicine, targeting of NLR pathways (e.g. CXCR2 inhibitors) in high-risk GDM cohorts is recommended.

### NLR in Rheumatic Diseases: A Bridge Between Inflammation and Autoimmunity

The Neutrophil to lymphocyte ratio (NLR) has emerged as a dynamic biomarker in rheumatic diseases, reflecting both systemic inflammation and immune dysregulation[74]. Its utility is demonstrated by its application in the assessment of disease activity, prognosis prediction, and therapeutic monitoring across a range of conditions, including rheumatoid arthritis (RA)[75], systemic lupus erythematosus (SLE), and ankylosing spondylitis (AS).

In RA, elevated levels of the Neutrophil to lymphocyte ratio (NLR) have been demonstrated to correlate with synovial neutrophil infiltration and pro-inflammatory cytokine release (e.g., IL-6, IL-17)[76,77]. A meta-analysis of 15 studies demonstrated that a Neutrophil to lymphocyte ratio (NLR) of 3.2 or more is indicative of a higher Disease Activity Score-28 (DAS-28) (pooled odds ratio [OR] = 2.8)[78] and radiographic progression (hazard ratio [HR] = 1.9). Mechanistically, the process of neutrophil extracellular traps (NETs) has been demonstrated to drive citrullinated autoantigen production, thereby exacerbating anti-cyclic citrullinated peptide (anti-CCP) antibody responses[74,79]. In contrast, lymphopenia (defined as a count of less than 1.2×109/L) is indicative of CD4+ T-cell exhaustion and is associated with a suboptimal response to TNF-α inhibitors (as indicated by a decrease in DAS-28 score of less than 1.2, p=0.03).

In SLE, NLR dynamics mirror disease flares. During periods of active nephritis, an elevated NLR (Neutrophil to lymphocyte ratio) greater than 4.5 has been shown to be a reliable predictor of renal biopsy severity (class IV lupus nephritis: odds ratio [OR] = 3.1) and the progression to end-stage renal disease (hazard ratio [HR] = 2.4) (Kim et al., 2020). Neutrophil activation via IFN-α signaling and complement deposition amplifies tissue damage, while lymphopenia (<0.8×10^9^/L) indicates immunosuppressive therapy toxicity[80,81]. It is noteworthy that the normalisation of NLR (defined as ≤2.0) following treatment has been shown to correlate with a reduction in flare frequency (RR=0.6).

Recent studies have emphasised the clinical significance of the Neutrophil to lymphocyte ratio (NLR) as a readily accessible biomarker reflecting systemic inflammation in ankylosing spondylitis (AS). Elevated Neutrophil to lymphocyte ratio (NLR) values (defined as >2.8) have been shown to be associated with higher C-reactive protein (CRP) levels, an increased risk of MRI-detected sacroiliitis (odds ratio ≈ 2.5), and a greater likelihood of syndesmophyte progression over time (hazard ratio ≈ 1.7). From a pathophysiological perspective, the release of matrix metalloproteinases (MMPs) by neutrophils has been identified as a key factor in the tissue damage observed in AS. These enzymes are involved in the degradation of cartilage, contributing to the inflammatory response in the affected tissues. Furthermore, the presence of relative lymphopenia has been correlated with overactivation of the IL-23/Th17 axis, a hallmark of AS immunopathology. In addition, a number of prospective cohort studies have demonstrated that a decrease in the Neutrophil to lymphocyte ratio (NLR) (ΔNLR ≥1.0) following the administration of interleukin-17 (IL-17) inhibitors is a reliable predictor of substantial improvement in disease activity, as measured by the Bath Ankylosing Spondylitis Disease Activity Index (BASDAI) (β ≈ 0.4, p < 0.01). Collectively, these findings suggest that NLR not only serves as a marker for disease activity and inflammatory burden in AS, but also may be used to assess and monitor therapeutic responses, providing a simple and cost-effective tool in both clinical and research settings.

Moreover, A number of studies have been conducted on the use of NLR in a range of other rheumatic diseases, including psoriatic arthritis[82], systemic sclerosis and vasculitis. In psoriatic arthritis, a positive correlation has been demonstrated between the Neutrophil to lymphocyte ratio (NLR) and disease activity, as well as skin involvement. In systemic sclerosis, the Neutrophil to lymphocyte ratio (NLR) has been demonstrated to be associated with disease severity and pulmonary arterial hypertension. In the context of vasculitis, the Neutrophil to lymphocyte ratio (NLR) has been identified as a potential marker of disease activity and response to treatment.

The correlation between the Neutrophil to lymphocyte ratio (NLR) and a range of rheumatic diseases underscores its promise as a straightforward, economical, and effective biomarker for evaluating disease activity, predicting prognosis, and gauging treatment response[83]. NLR offers several advantages over traditional inflammatory markers, such as CRP and ESR, including its stability and ease of measurement. However, further research is required to standardise NLR measurement protocols and to determine optimal cutoff values for different rheumatic diseases. It is recommended that future studies concentrate on large-scale, multicentre cohorts, with a view to investigating the potential of NLR in combination with other biomarkers. The aim of this would be to enhance the diagnostic and prognostic accuracy of rheumatic diseases.

### NLR in COVID-19: A Hallmark of Immune Dysregulation and Disease Severity

The Neutrophil to lymphocyte ratio (NLR) has emerged as a pivotal biomarker in cases of Coronavirus Disease 2019 (covid-19), reflecting the interplay between hyperinflammation and immune exhaustion[15,84]. Its prognostic utility is evident in the early risk stratification, severity prediction and mortality assessment of patients, particularly in the context of viral-induced cytokine storms and secondary organ damage[85,86].

NLR has been demonstrated to serve as a predictor of disease progression. Elevated Neutrophil to lymphocyte ratio (NLR) levels have been demonstrated to be a significant predictor of the severity of cases of Coronavirus disease 2019 (Covid-19)[87,88]. A meta-analysis of 25 studies demonstrated that a NLR >6.5 has a pooled odds ratio (OR) of 4.2 (95% CI: 3.1–5.7) for predicting progression from mild to severe disease[87]. In addition, in a cohort of hospitalised patients, an NLR >9.0 has been shown to correlate with the development of acute respiratory distress syndrome (ARDS) (HR=3.8) and intensive care unit (ICU) admission (HR=2.9)[61,89]. Mechanistically, the infection of SARS-CoV-2 activates the release of the chemokine IL-8, which in turn activates the CXCR2 receptor on the surface of neutrophils. This results in an increase in the number of neutrophils in the blood, known as neutrophilia. The infection also leads to the death of T-cells, a type of white blood cell, through a process called apoptosis[90]. This combination of increased neutrophilia and decreased T-cell count contributes to an imbalance in the body’s natural killer (NK) cells, known as the NLR balance[84].

There is a strong correlation between NLR dynamics and mortality from COVD-19. A multicentre cohort study reported that a Neutrophil to lymphocyte ratio (NLR) >12.3 at admission predicts 28-day mortality with 82% sensitivity and 76% specificity (area under the curve [AUC] = 0.88). Furthermore, persistent NLR elevation (>10) beyond day 7 of hospitalization has been shown to be associated with a 5-fold increased risk of death, independent of age and comorbidities (HR=5.1, p<0.001)[85,91]. This finding is indicative of unresolved inflammation and impaired viral clearance, particularly in elderly and immunocompromised populations[86,92].

NLR has been shown to have significant prognostic value in relation to cases of the novel corona virus (SARS-CoV-2). This is due to the fact that it is indicative of both innate and adaptive immune dysfunction[15]. Firstly, elevated NLR values may be indicative of neutrophil hyperactivation. Neutrophil extracellular traps (NETs) have been demonstrated to contribute to microthrombosis and endothelial injury, resulting in elevated D-dimer levels (r=0.52, p<0.01)[93,94]. Furthermore, an elevated NLR may also serve as an indicator of lymphocyte exhaustion[90]. Lastly, CD4^+^ and CD8^+^ T-cell depletion (lymphocyte count <0.8×10^9^/L)[95] has been shown to correlate with viral persistence and secondary bacterial infections (OR=2.4)[92].

Moreover, it can be argued that NLR can serve as an indicator for both clinical applications and therapeutic implications. In the initial triage, a Neutrophil to lymphocyte ratio (NLR) greater than 5.0 at the time of emergency department presentation prioritises the allocation of intensive care unit resources (NLR >5.0 vs. ≤ 5.0: mortality 23% vs. 4%, p<0.001)[87,89,96]. With regard to the monitoring of therapeutic interventions, a reduction in the Neutrophil to lymphocyte ratio (ΔNLR ≥ 3.0) following dexamethasone therapy has been shown to predict enhanced oxygenation (PaO2/FiO2 ratio increase >50 mmHg, p=0.02)[97,98]. In conclusion, the integration of NLR with ferritin (>1000 ng/mL) and lactate dehydrogenase (>350 U/L) has been demonstrated to enhance mortality prediction, with an area under the curve (AUC) value of 0.94[99].

Recent research has begun to explore the role of the Neutrophil to lymphocyte ratio (NLR) in the prolonged effects of SARS-CoV-2, with elevated NLR levels (defined as >3.5) being associated with ongoing fatigue (odds ratio [OR] = 2.1) and cognitive impairment (OR = 1.8) at the 6-month follow-up stage[100,101]. Furthermore, there is ongoing clinical validation of NLR-driven algorithms for antiviral therapy (e.g. the timing of remdesivir administration) and immunomodulation (e.g. the selection of IL-6 inhibitors). The neutrophil to lymphocyte ratio (NLR) has emerged as a versatile biomarker across diverse medical fields, reflecting systemic inflammation, immune dysregulation, and disease progression[84].

### NLR and other diseases

In addition to the applications previously mentioned, NLR has been demonstrated to possess diagnostic and therapeutic value in a wide range of other diseases. For instance, the presence of an elevated NLR has been observed to be associated with the development of neurological diseases, such as Alzheimer’s and Parkinson’s disease, in which inflammation plays a pivotal role[102,103]. NLR alterations have the capacity to mirror neuroinflammation and neuronal impairment, thereby providing a foundation for the early diagnosis and prognosis assessment of these conditions. In the context of stroke-related diseases, elevated NLR levels, defined as >5.0 at admission, have been shown to be a significant predictor of neurological deterioration[104]. In Alzheimer’s disease, a NLR greater than 2.8 has been shown to be a significant predictor of accelerated cognitive decline, as indicated by a minimum of 3 points decrease on the Mini-Mental State Examination (MMSE) per year (p = 0.01)[103].

In the domain of kidney diseases, the NLR has been demonstrated to be associated with the manifestation, progression, and prognosis of renal diseases[105]. In the context of diabetic nephropathy, chronic glomerulonephritis, and acute kidney injury, elevated NLR has been demonstrated to be a reliable indicator of increased inflammation and kidney damage[106]. NLR has been demonstrated to possess the capacity to predict disease progression and adverse outcomes, thereby facilitating treatment decisions. In particular, elevated NLR > 3.0 has been demonstrated to be associated with renal fibrosis progression (a twofold increase in the rate of decline in estimated glomerular filtration rate [eGFR]) and cardiovascular events in dialysis patients with chronic kidney disease (CKD).

In addition, in the context of liver-related diseases, elevated NLR levels have been observed to be associated with advanced fibrosis[107]. Specifically, NLR levels greater than 2.5 have been shown to be a reliable predictor of advanced fibrosis[108]. Additionally, NLR levels exceeding 4.0 have been identified as a significant risk factor for spontaneous bacterial peritonitis.

### Summary

This comprehensive analysis underscores the Neutrophil to Lymphocyte Ratio (NLR) as a remarkably versatile, accessible, and cost-effective biomarker with significant clinical utility across a broad spectrum of medical disciplines[87,109]. The exponential growth in research related to NLR, particularly that contributed by Chinese scholars, reflects an increasing global recognition of the field. NLR has been shown to have consistently robust prognostic and predictive value in a number of medical areas. In the field of oncology, for example, it has been used to predict survival, therapy response, and immunotherapy benefit[12]. In the area of cardiovascular diseases, it has been employed to stratify risk and predict adverse events. In the context of sepsis/SIRS/MODS, it has been useful in aiding diagnosis, severity assessment, and predicting organ failure[110]. Diabetes mellitus is characterised by chronic inflammation, which can predict the development of complications and the progression of the disease. Rheumatic diseases are evaluated based on factors such as disease activity, progression, and treatment response[74,111]. Notably, the novel severe acute respiratory syndrome (SARS-CoV-2) virus, which causes the disease known as Coronavirus disease 2019 (Covid-19), can serve as a key indicator of disease severity, progression to critical illness, and mortality[86].

In conclusion, NLR is a simple, cost-effective, and valuable biomarker with significant clinical utility across various diseases[87,109]. Its predictive mechanisms involve multiple aspects, including immune response imbalances, cytokine effects, oxidative stress, and disease-specific immune reactions[12]. The strength of the Neutrophil to lymphocyte ratio (NLR) lies in its ability to integrate information on both innate (neutrophil-driven inflammation) and adaptive (lymphocyte-mediated regulation) immune responses, thereby providing a comprehensive overview of systemic inflammation and immune dysregulation[111]. The fact that it is derived from a routine complete blood count means that it can be implemented in a variety of clinical settings[87,109]. While it has been established as a powerful standalone indicator, future research holds immense promise in refining its application. Key directions for future research include standardising measurement protocols and optimal cut-off values across different diseases and populations, integrating NLR with other biomarkers (genetic, inflammatory, metabolic) and clinical data through advanced analytics (e.g. machine learning) to enhance diagnostic and prognostic accuracy, and exploring its potential as a dynamic tool for guiding personalised therapeutic interventions (e.g. immunotherapy timing, anti-inflammatory strategies)[17]. Addressing these challenges will serve to consolidate NLR’s position in the advancement of precision medicine and the enhancement of patient outcomes on a global scale[111].

### Research Significance and Future Directions

This pioneering bibliometric mapping of NLR research makes three significant contributions, thus surpassing conventional reviews. Firstly, it establishes neutrophil-lymphocyte ratio (NLR) as a dynamic immune-inflammatory integrator rather than a standalone biomarker. This unifies 20 years of multidisciplinary evidence into a paradigm-shifting framework that links neutrophil-driven innate responses with lymphocyte-mediated adaptive regulation across disease states. These states range from cancer-induced immunosuppression to cytokine storms in cases of severe acute respiratory syndrome (SARS-CoV-2). Secondly, the analysis identifies actionable clinical priorities, namely the urgent need for standardised NLR protocols (context-specific cutoffs, measurement timing, and reporting criteria) to resolve current heterogeneity, alongside leveraging NLR’s cost-effectiveness for point-of-care deployment in resource-limited settings, particularly for sepsis triage and immunotherapy response monitoring. Thirdly, it delineates methodological imperatives: future studies must integrate multi-omics dimensions (e.g., combining NLR with TCF7L2 variants in diabetes or NETosis-related proteomics in cancer) through machine learning to construct predictive models for precision stratification, while expanding real-world validation via electronic health records in underrepresented populations like paediatric sepsis or geriatric oncology. It is imperative that mechanistic questions that have not yet been resolved are the focus of targeted investigation. This is particularly important in determining whether NLR elevation actively propagates pathology (for example, via neutrophil extracellular traps facilitating metastatic niches) or merely reflects bystander inflammation. Furthermore, it is essential to ascertain whether NLR-guided interventions (for example, CXCR2 inhibitors in gestational diabetes or IL-7 administration in lymphopenic sepsis) can demonstrably improve outcomes in randomised trials.As the role of the neutrophil-to-lymphocyte ratio (NLR) shifts from a prognostic indicator to a therapeutic compass, the present study provides the foundational cartography to facilitate its integration into global precision medicine frameworks, thereby transforming a simple hematologic ratio into a universal metric of immune resilience.

### Limitations

This study is pioneering in its use of bibliometric visualization to analyse all of the NLR-related research conducted over the past two decades. However, it inevitably has certain limitations. Firstly, it should be noted that the data utilised in this study is exclusively from the WOSCC database, excluding data from alternative databases, including PubMed, CNKI, Cochrane Library, and Google Scholar. Despite the comprehensiveness and reliability of the WOSCC, it is acknowledged that there is a possibility of certain articles being absent from the database. Secondly, it is pertinent to acknowledge that this study has been conducted using exclusively English-language literature, which may result in outcomes that are not representative of the wider population. Moreover, it is acknowledged that certain studies may encompass a range of disciplines, and it is recognised that a straightforward categorisation of the results by bibliometric analysis software, which may compromise the precision of the findings. Finally, the data in this study may be inconsistent in various aspects. For example, the same institution may have used different names at different time periods. In addition, it appears that the names of some of the authors have been altered, which has a consequential effect on the presentation of the study’s results.

### Conclusions

In this study, bibliometric analysis was employed to review the trends, hot spots, and frontiers of NLR-related research over the past two decades. FRONTIERS IN ONCOLOGY, MEDICINE, PLOS ONE etc., are influential journals in this field. ONCOLOGY, IMMUNOLOGY, and INFLAMMATION are hot topics in this field, and the relationship between NLR and immunotherapy and cancer-related diseases may be a direction for future research. Our study illustrates basic scientific knowledge and various interrelationships about neutrophil extracellular traps (NETs) and provides important clues about research trends and frontiers. We hope this study will help researchers better grasp current general trends in the field.

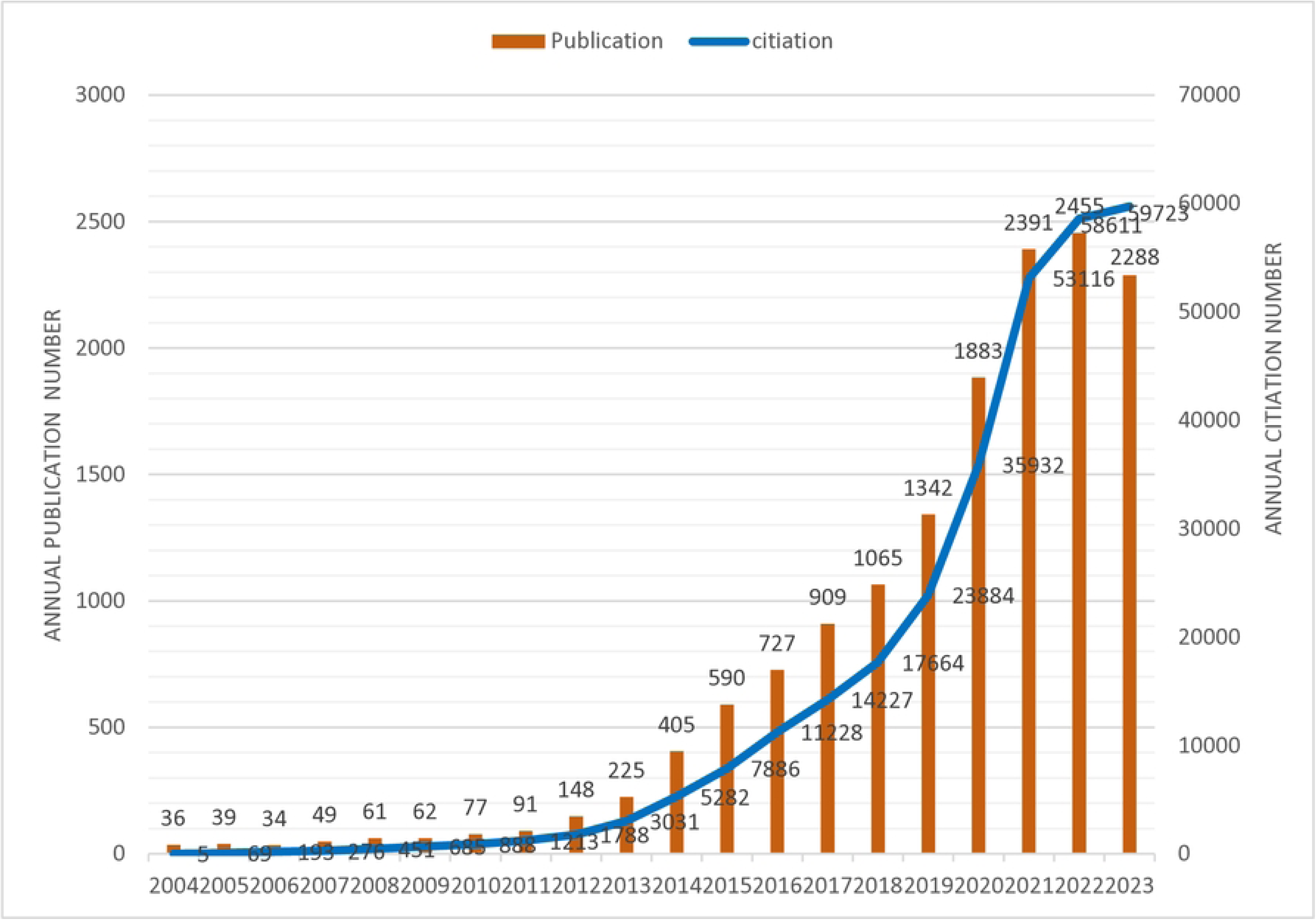

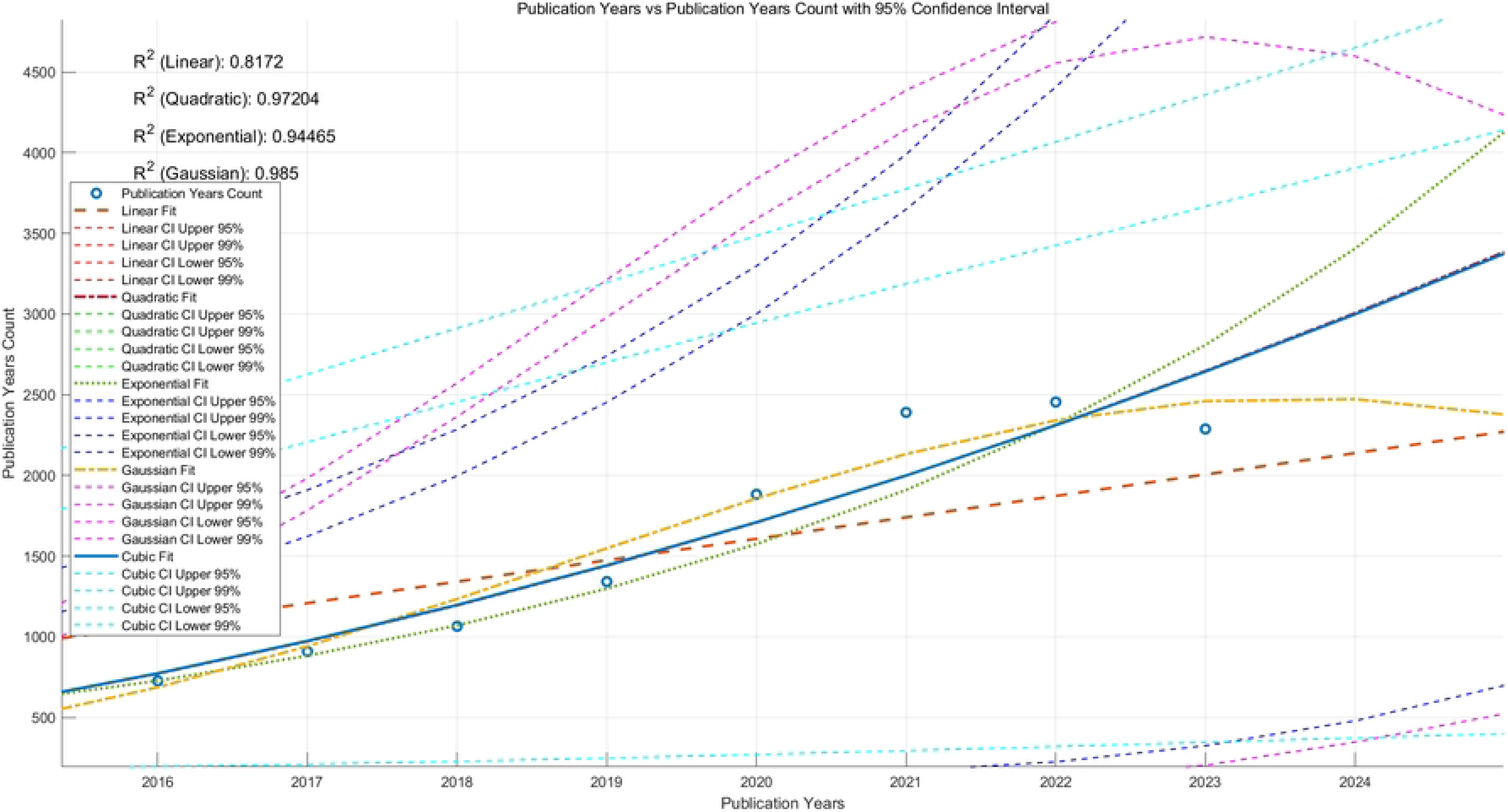

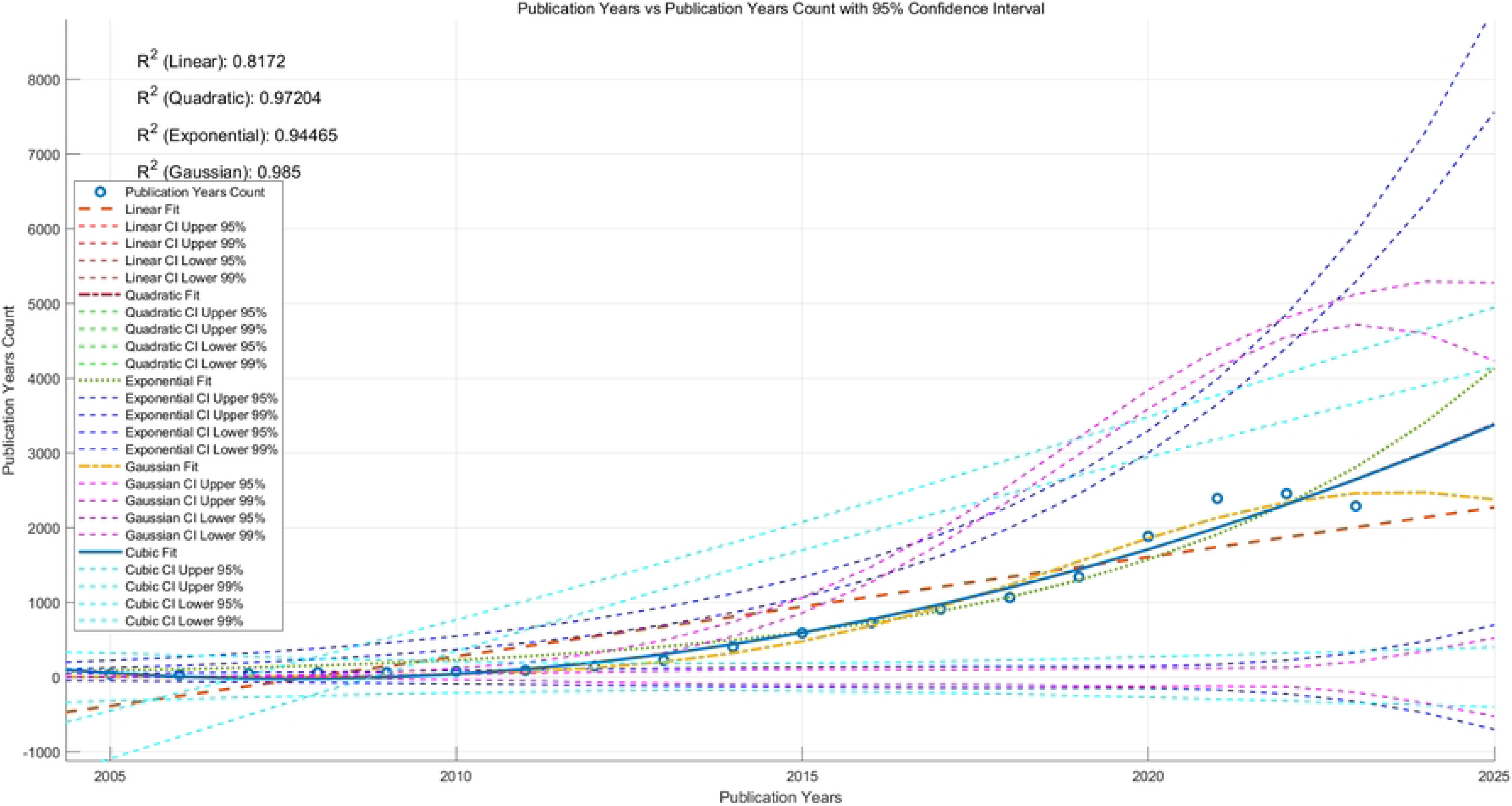

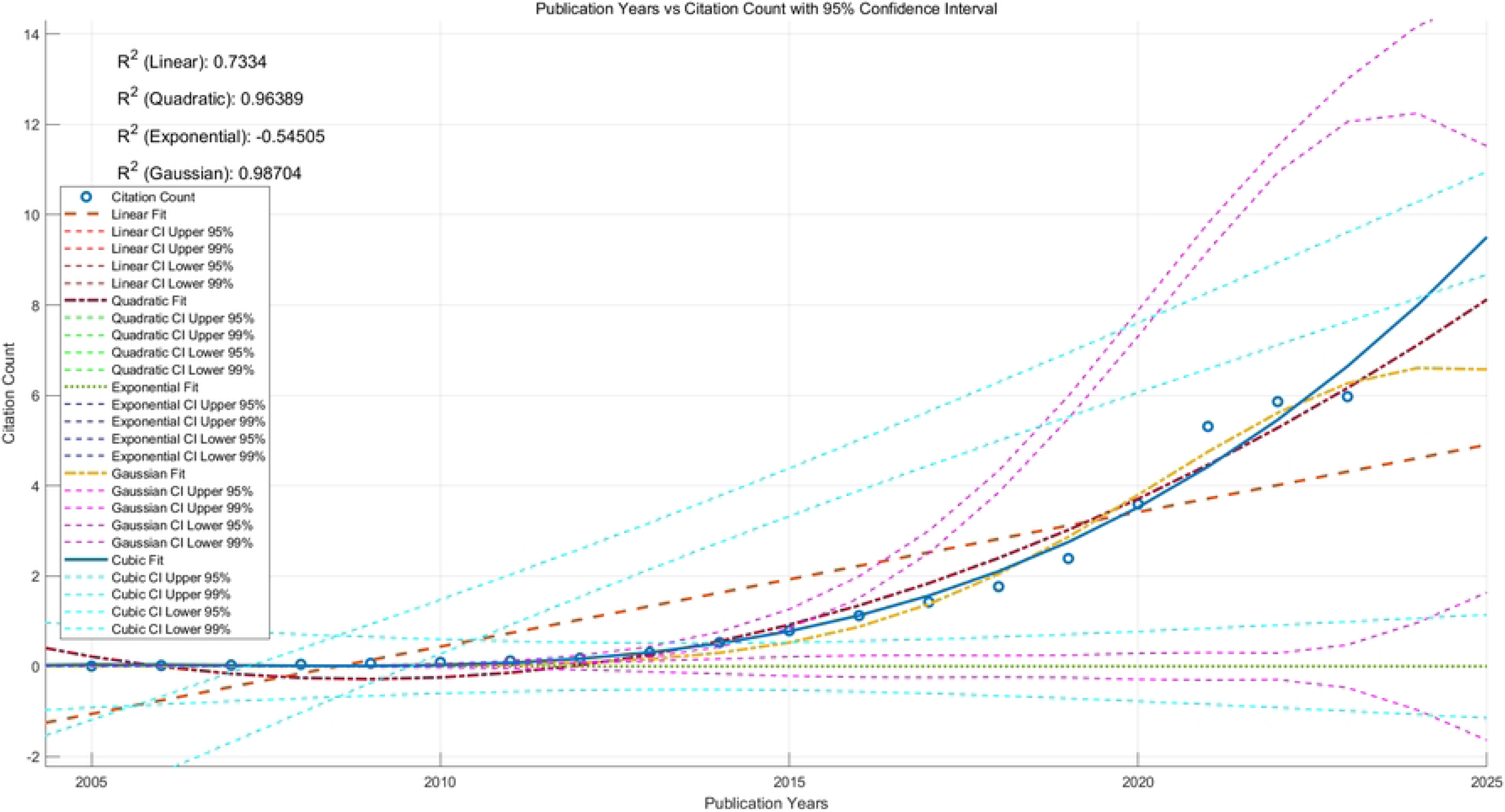

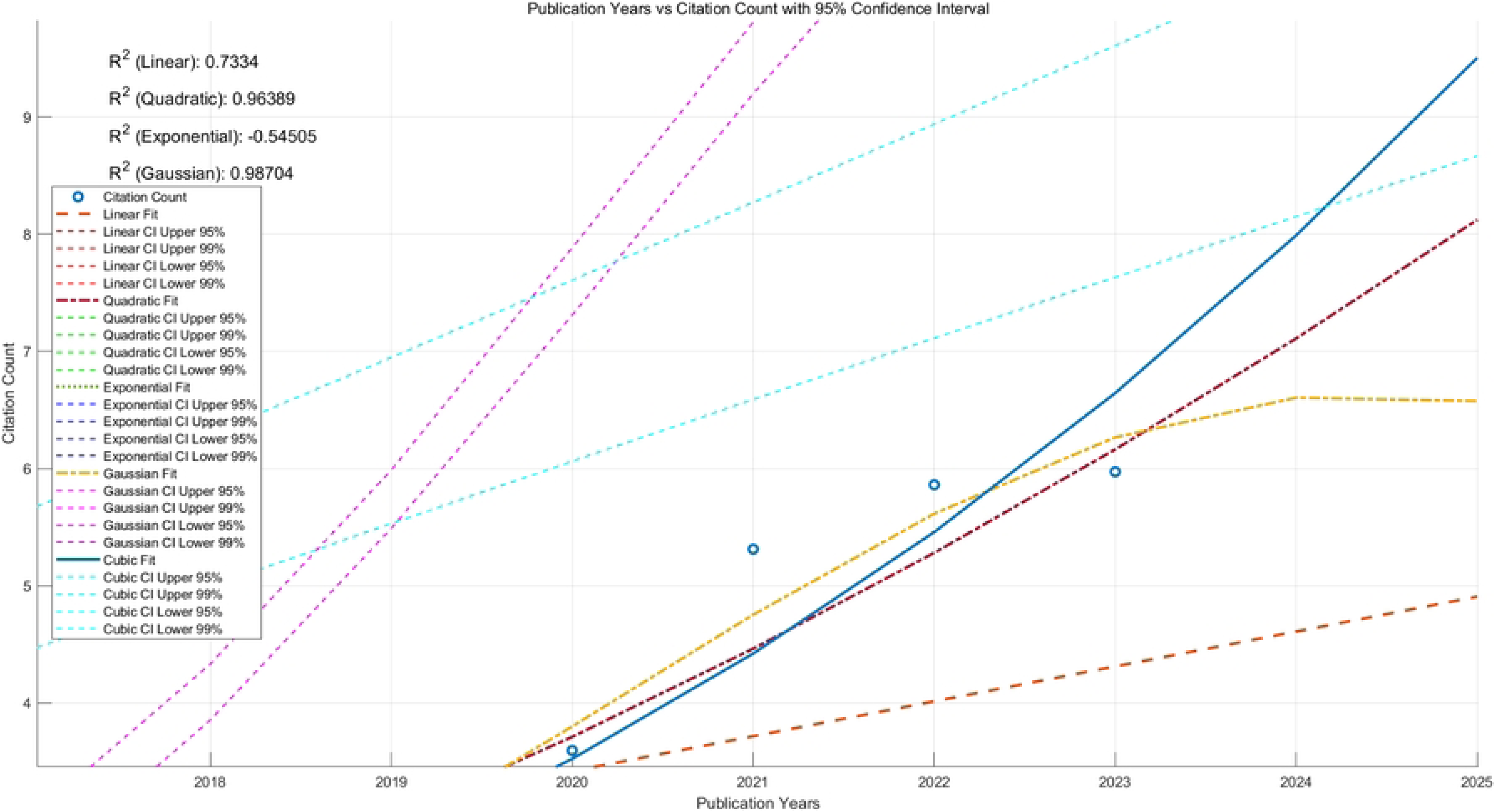

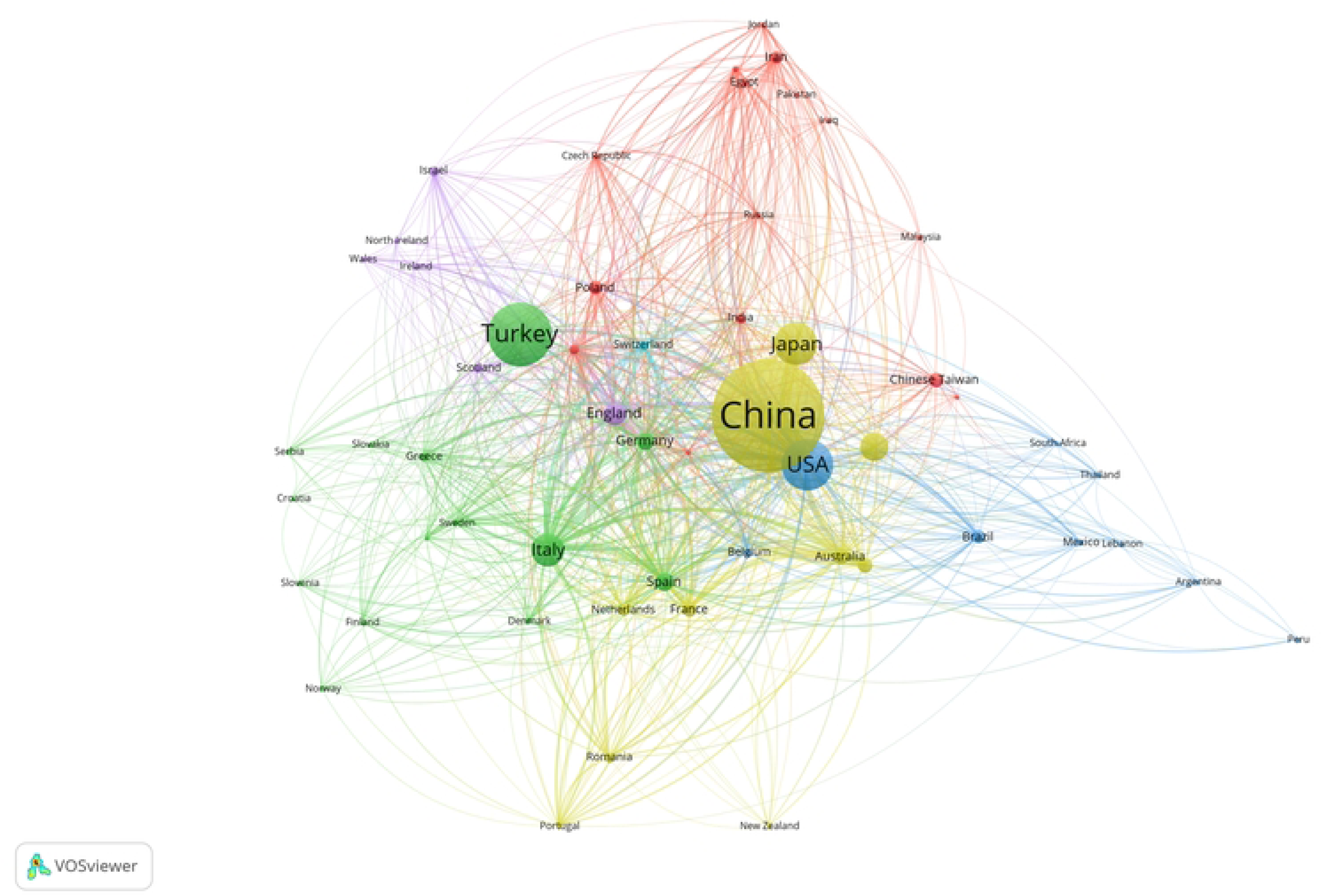

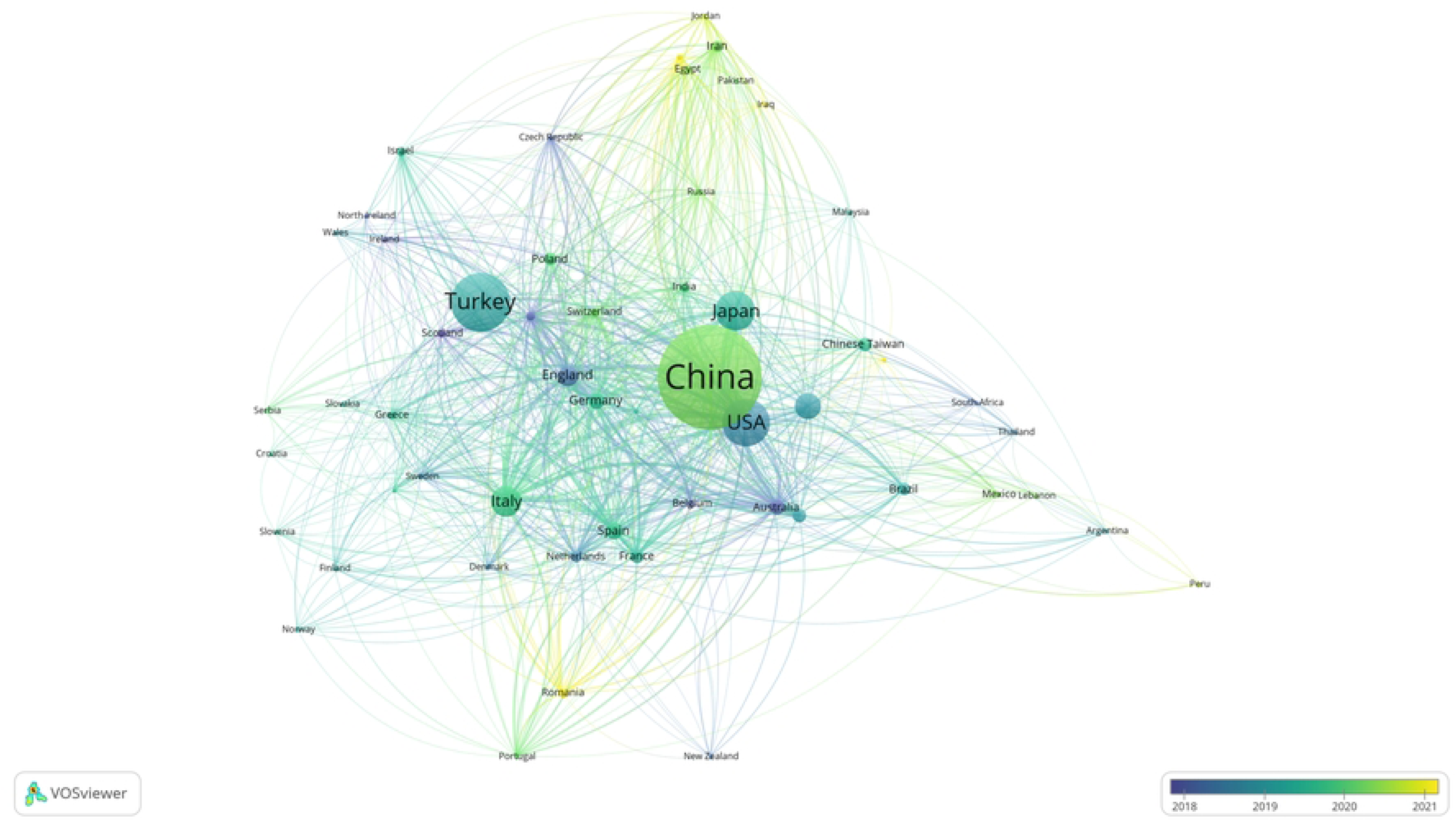

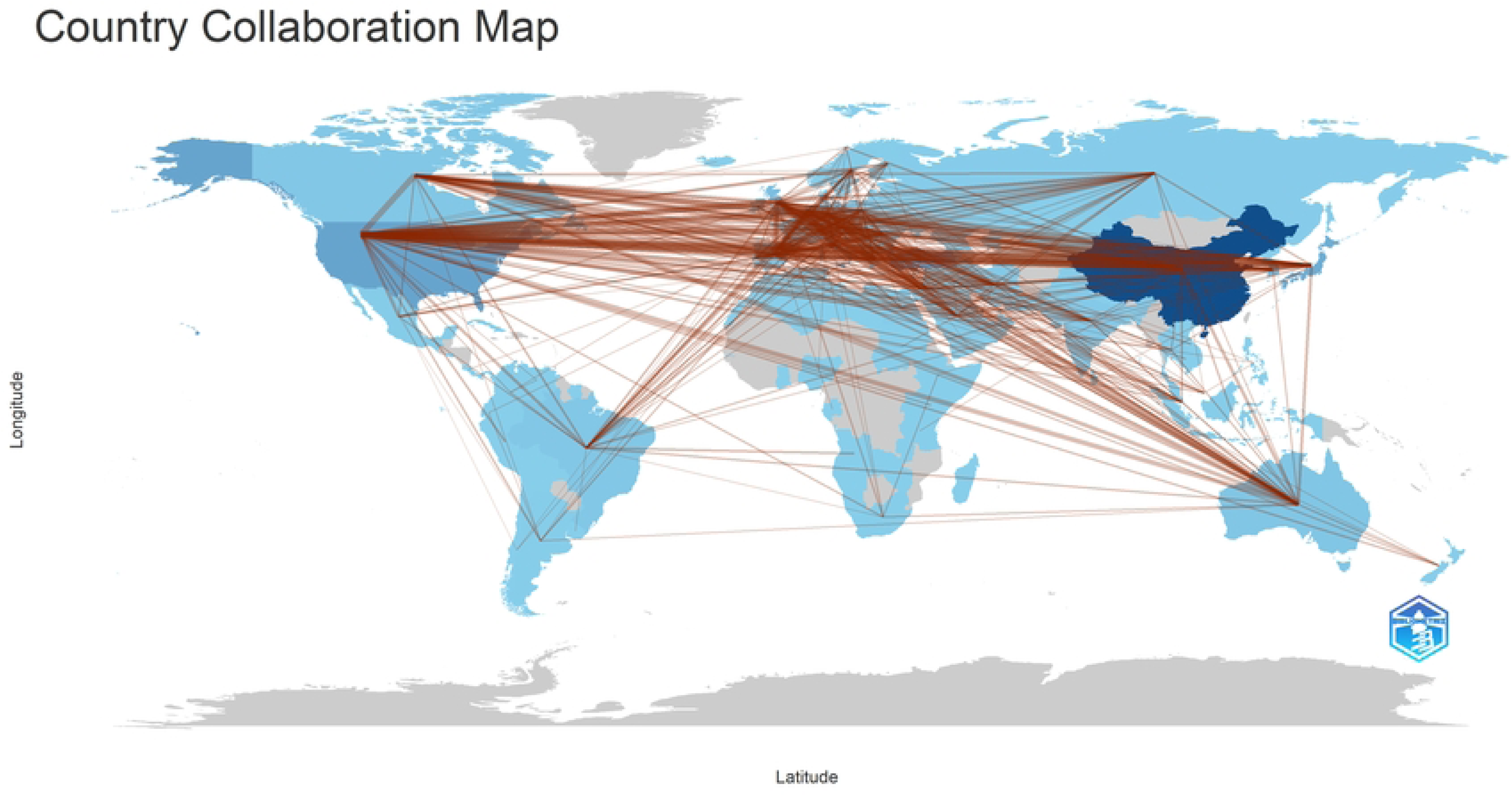

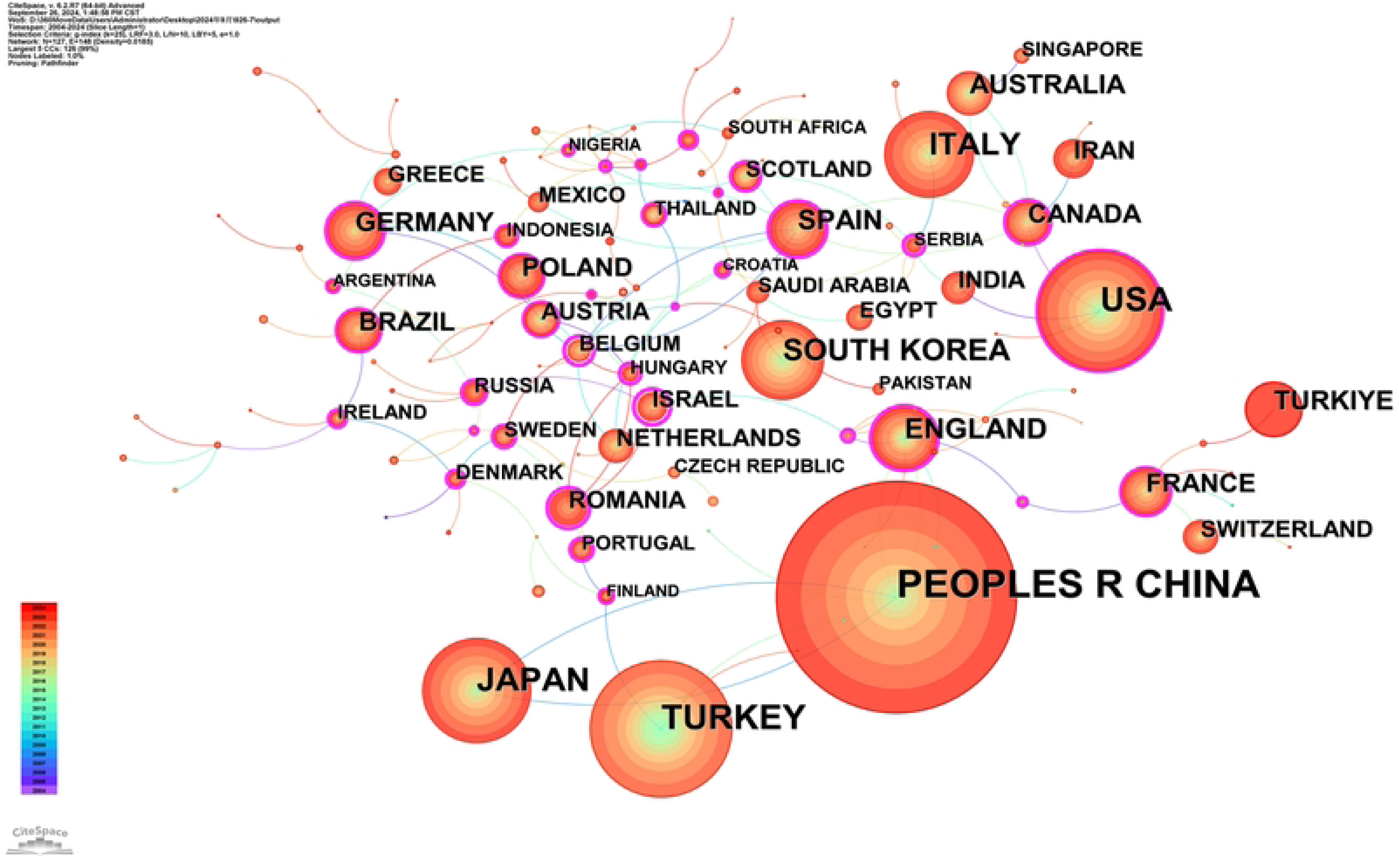

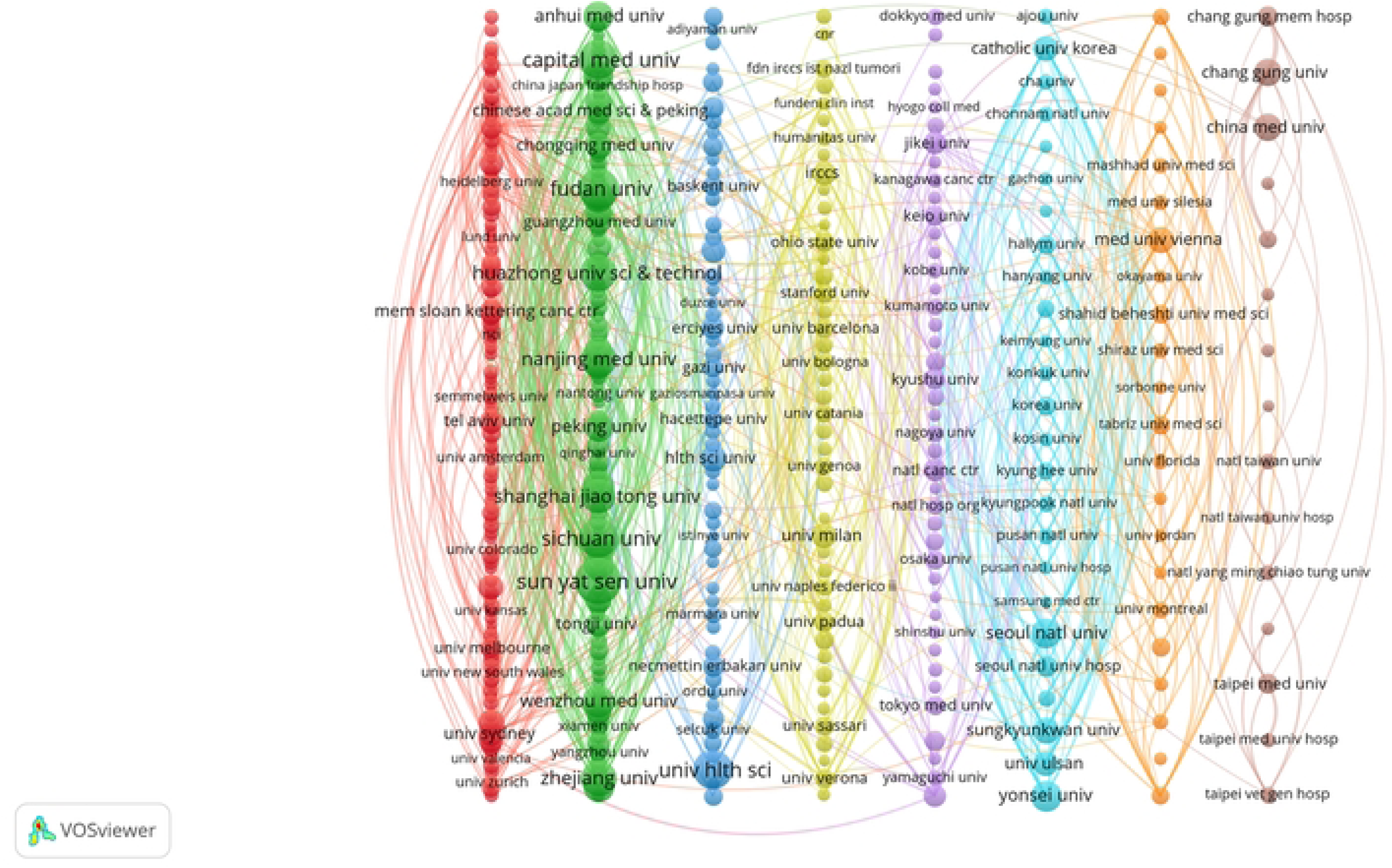

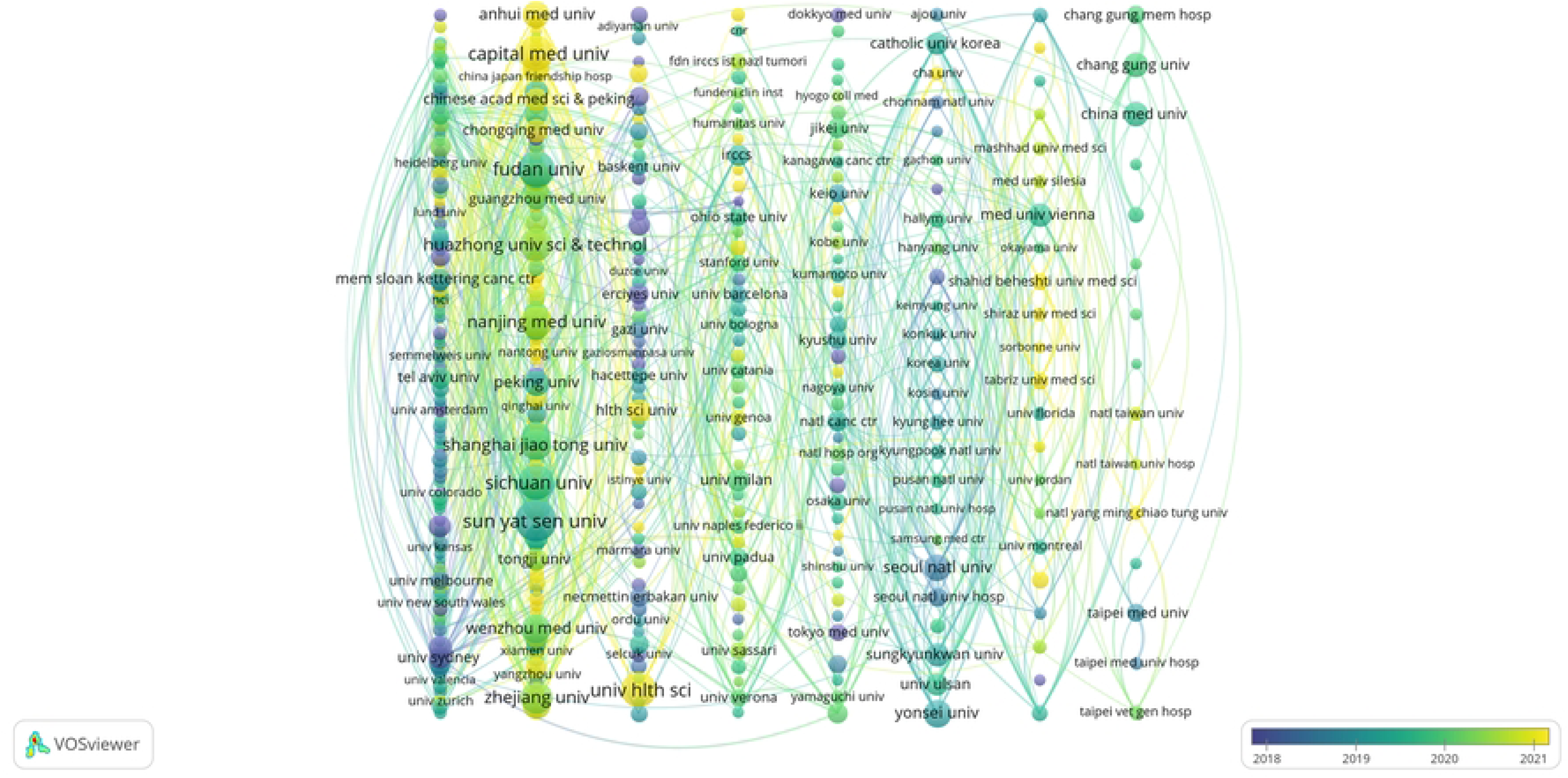

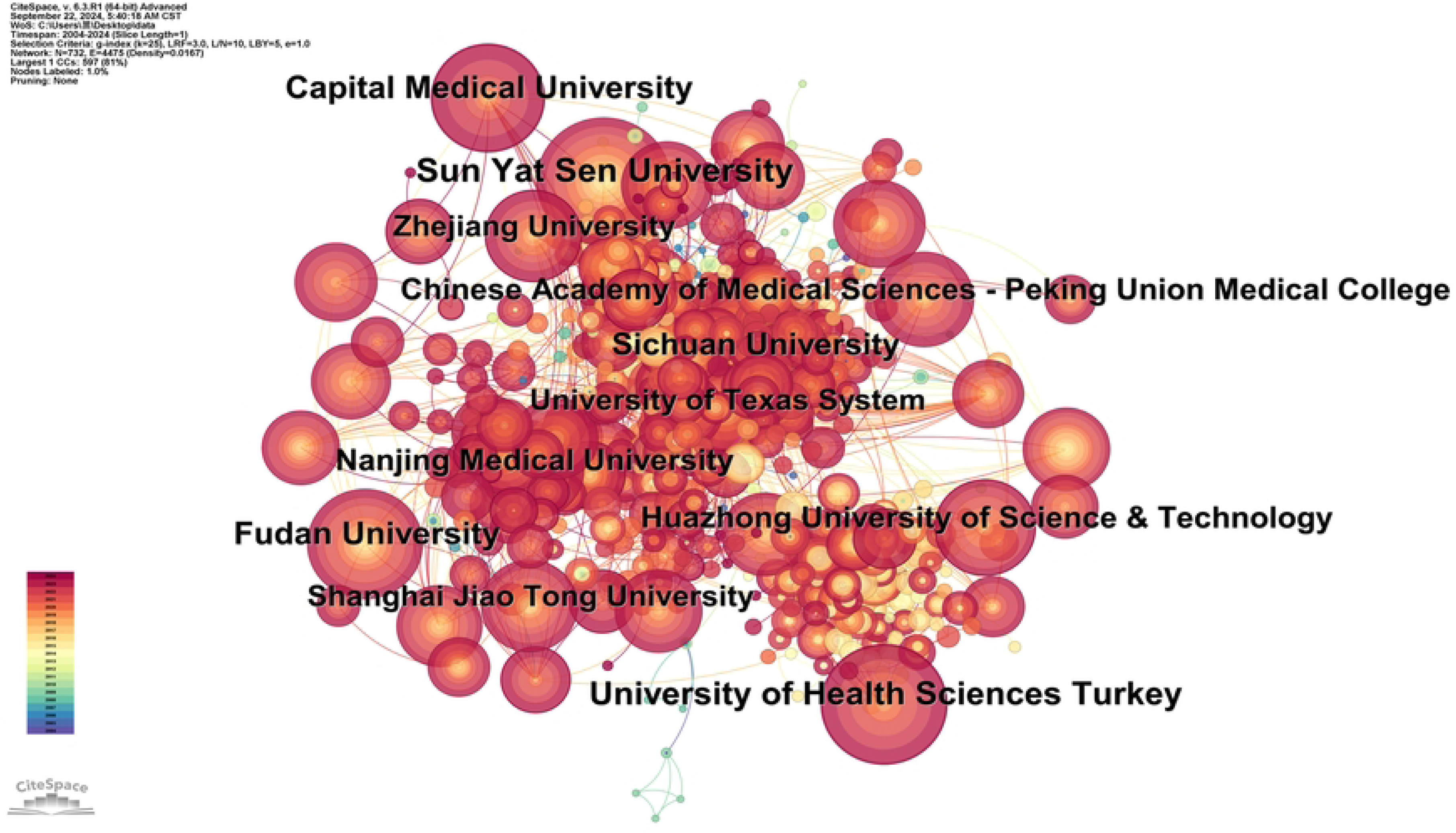

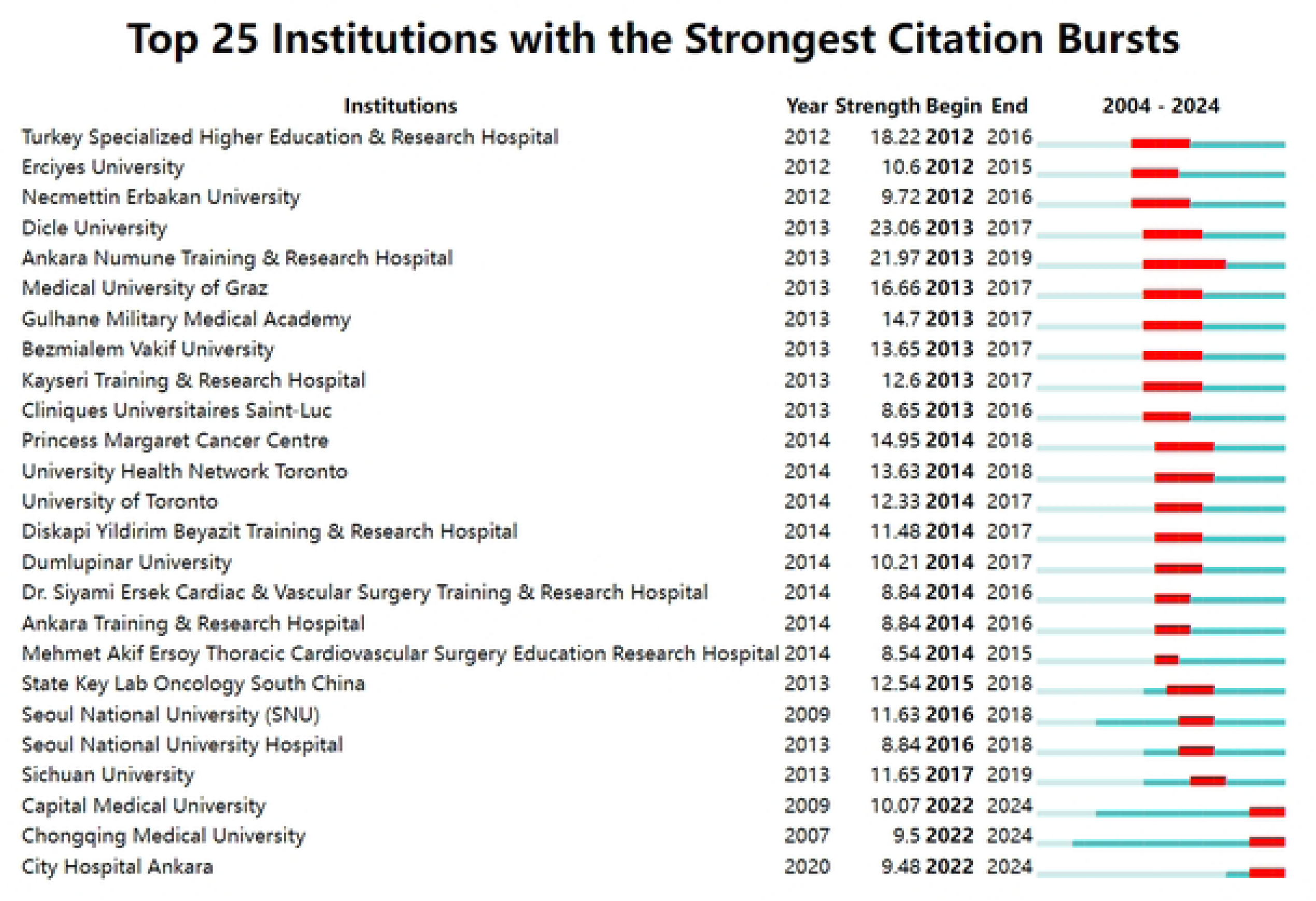

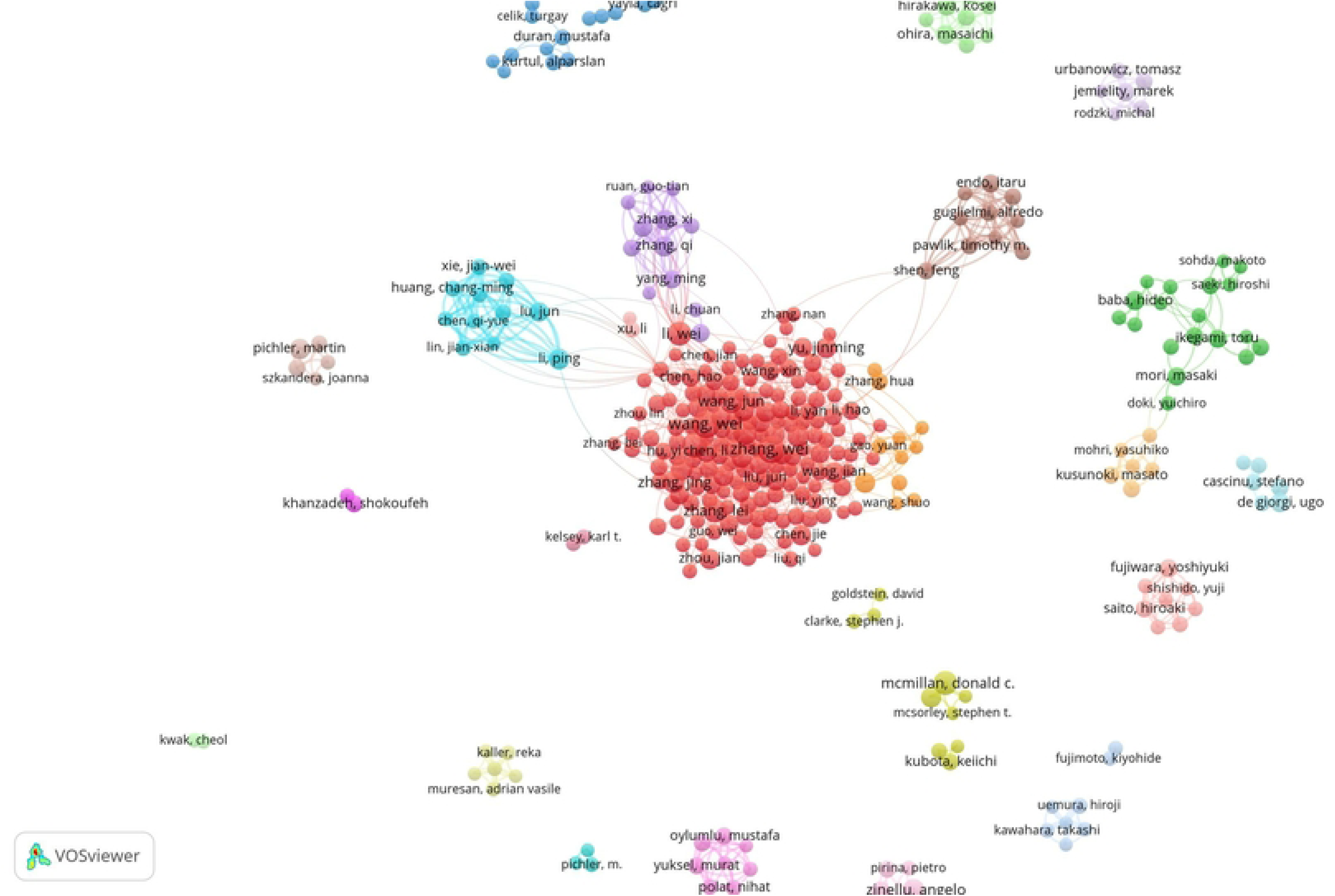

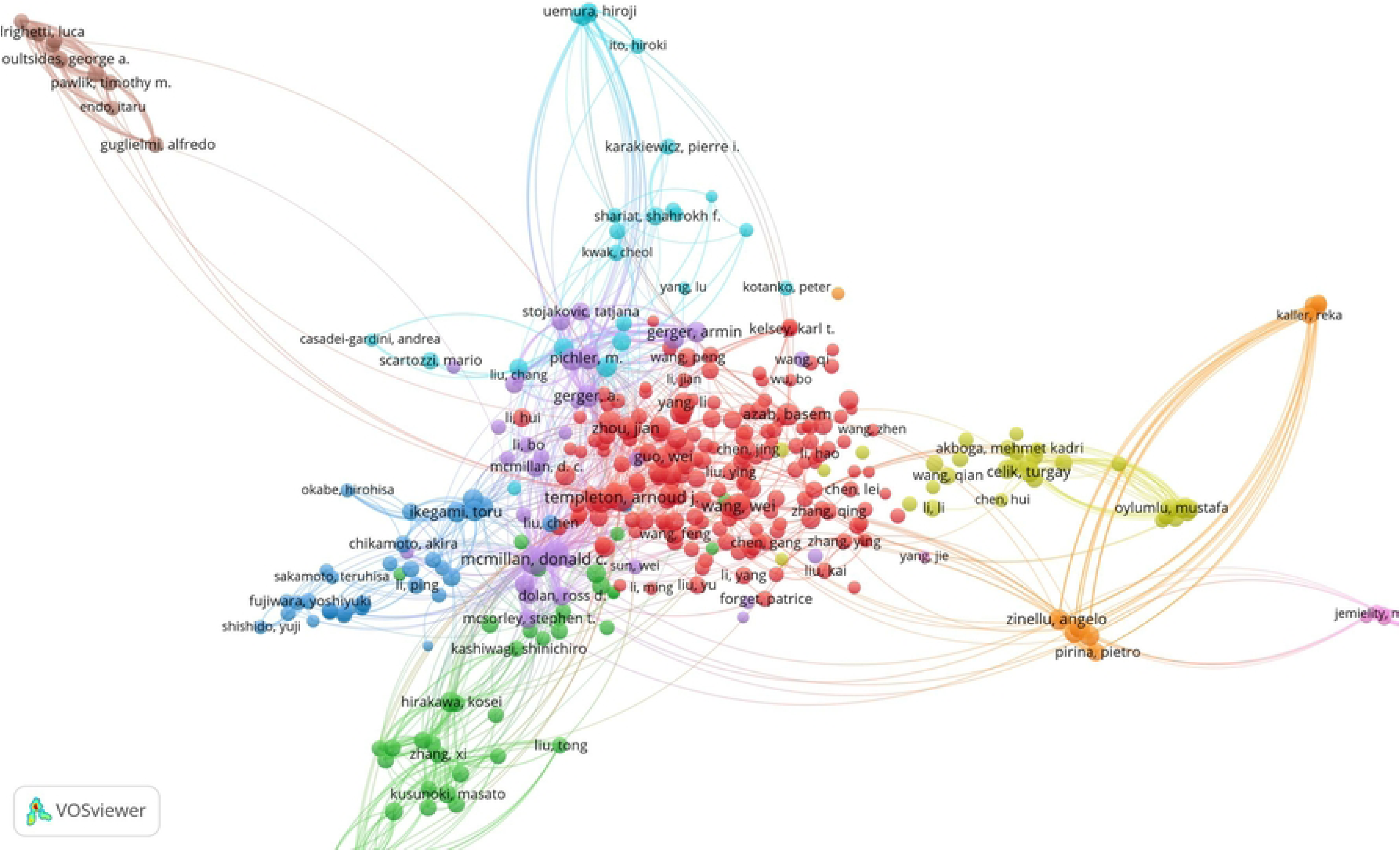

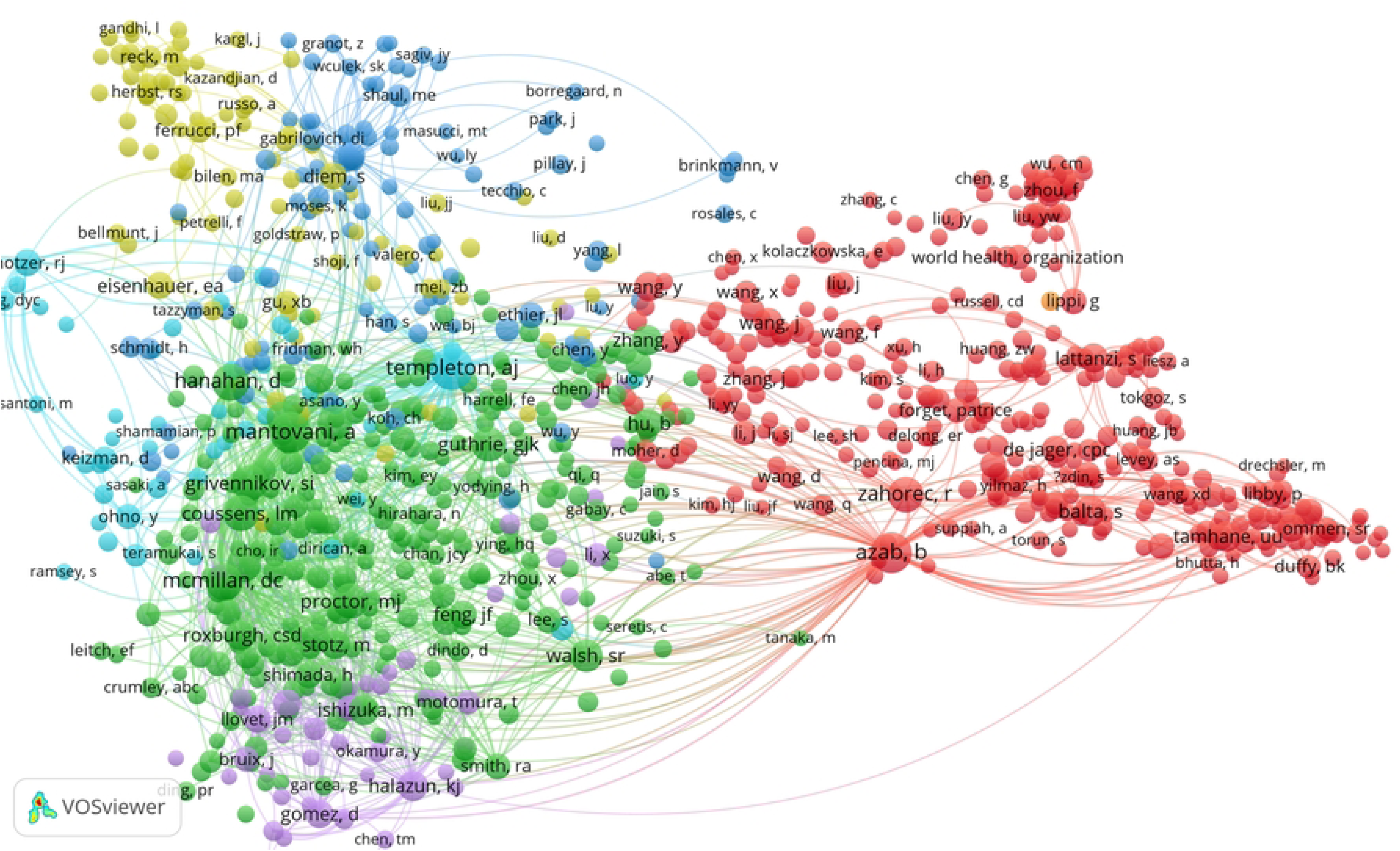

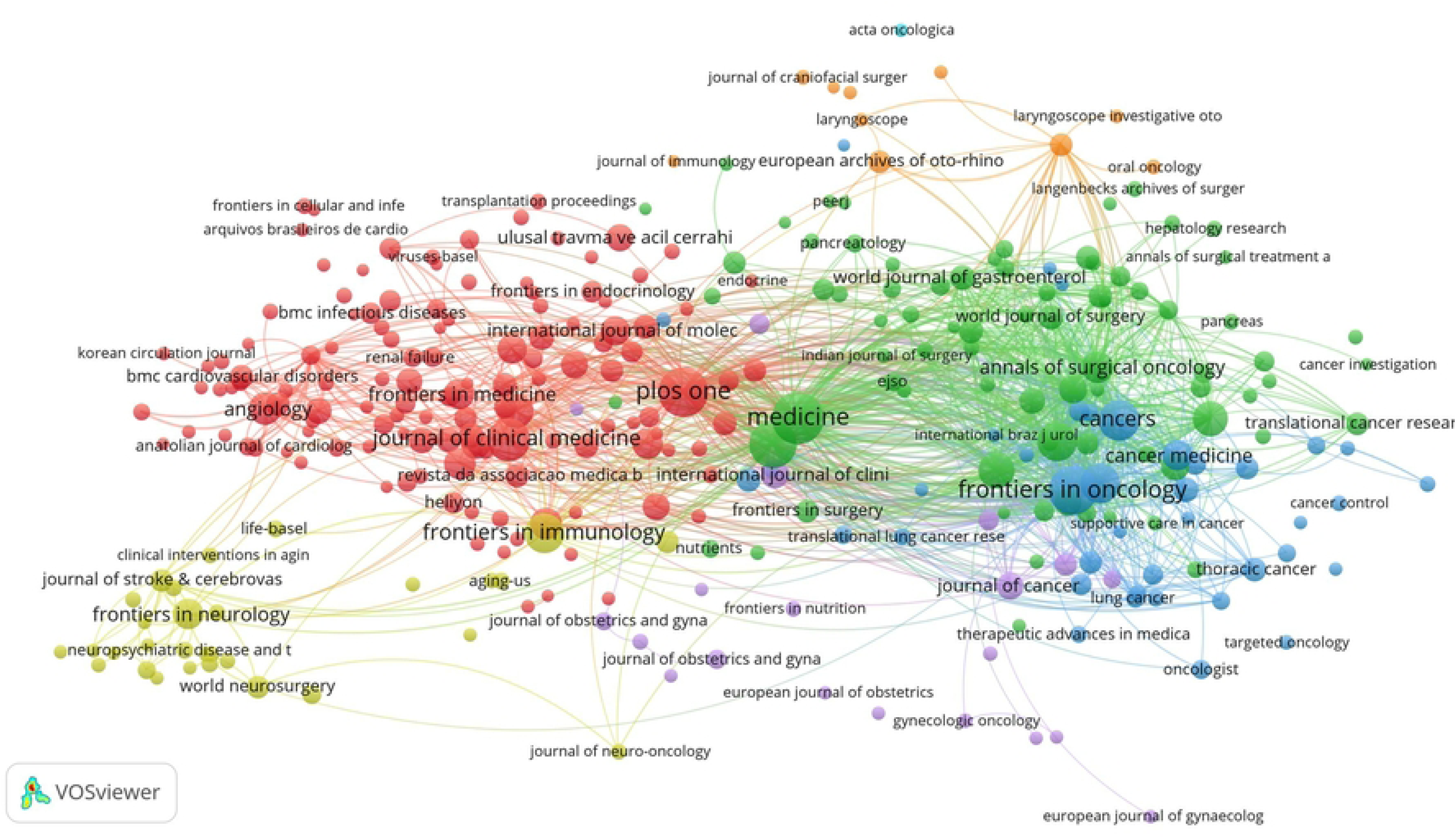

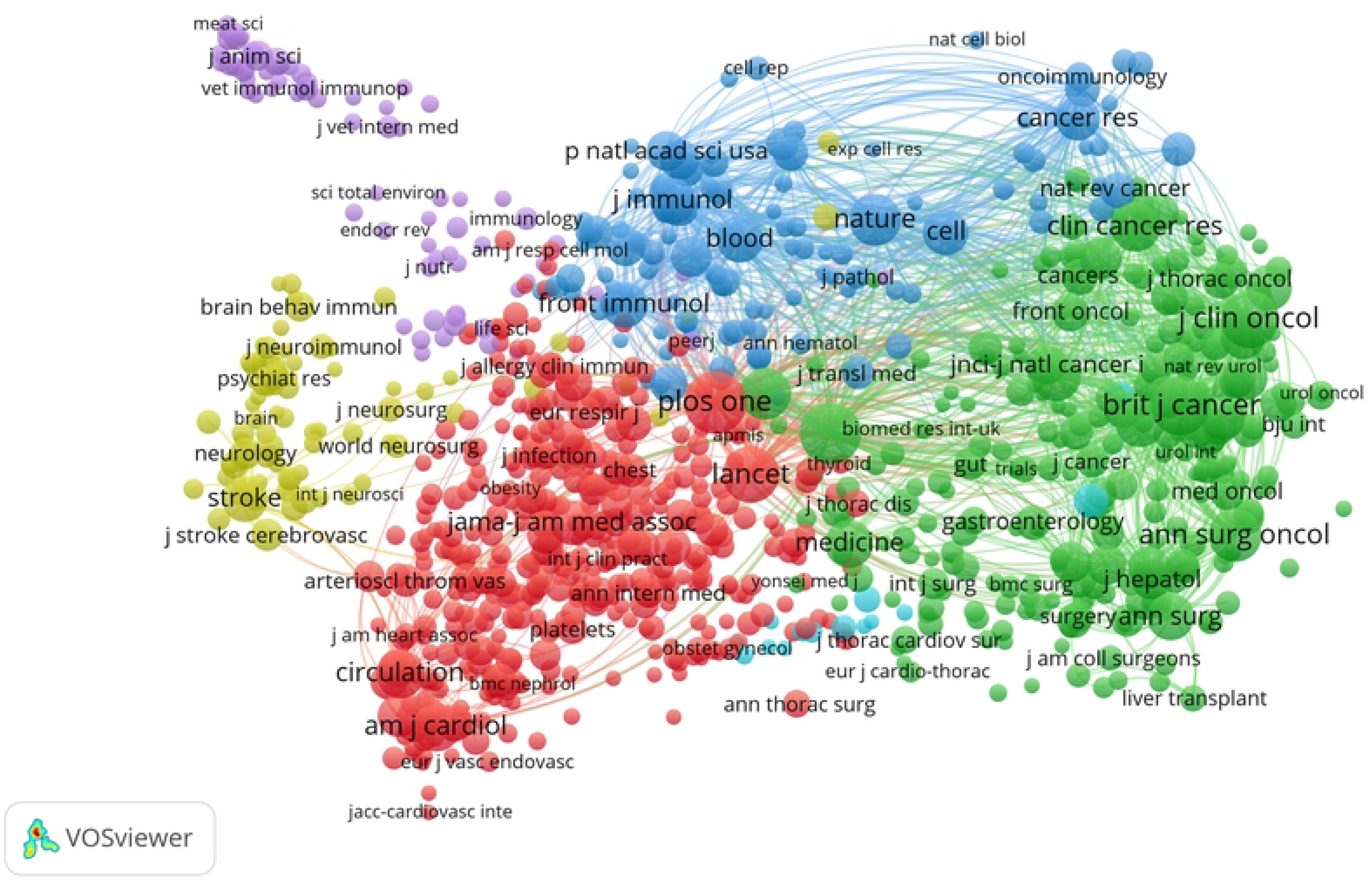

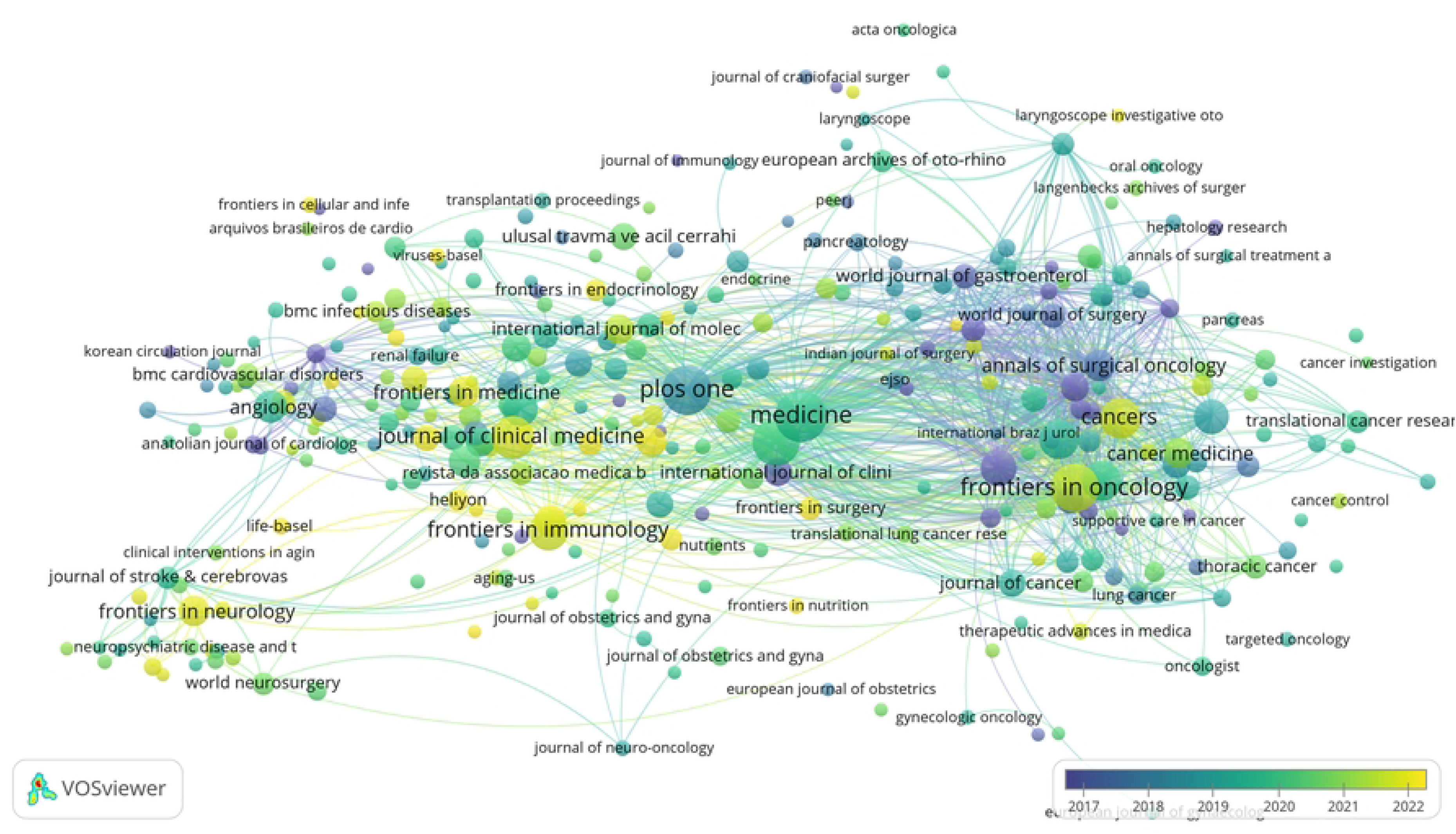

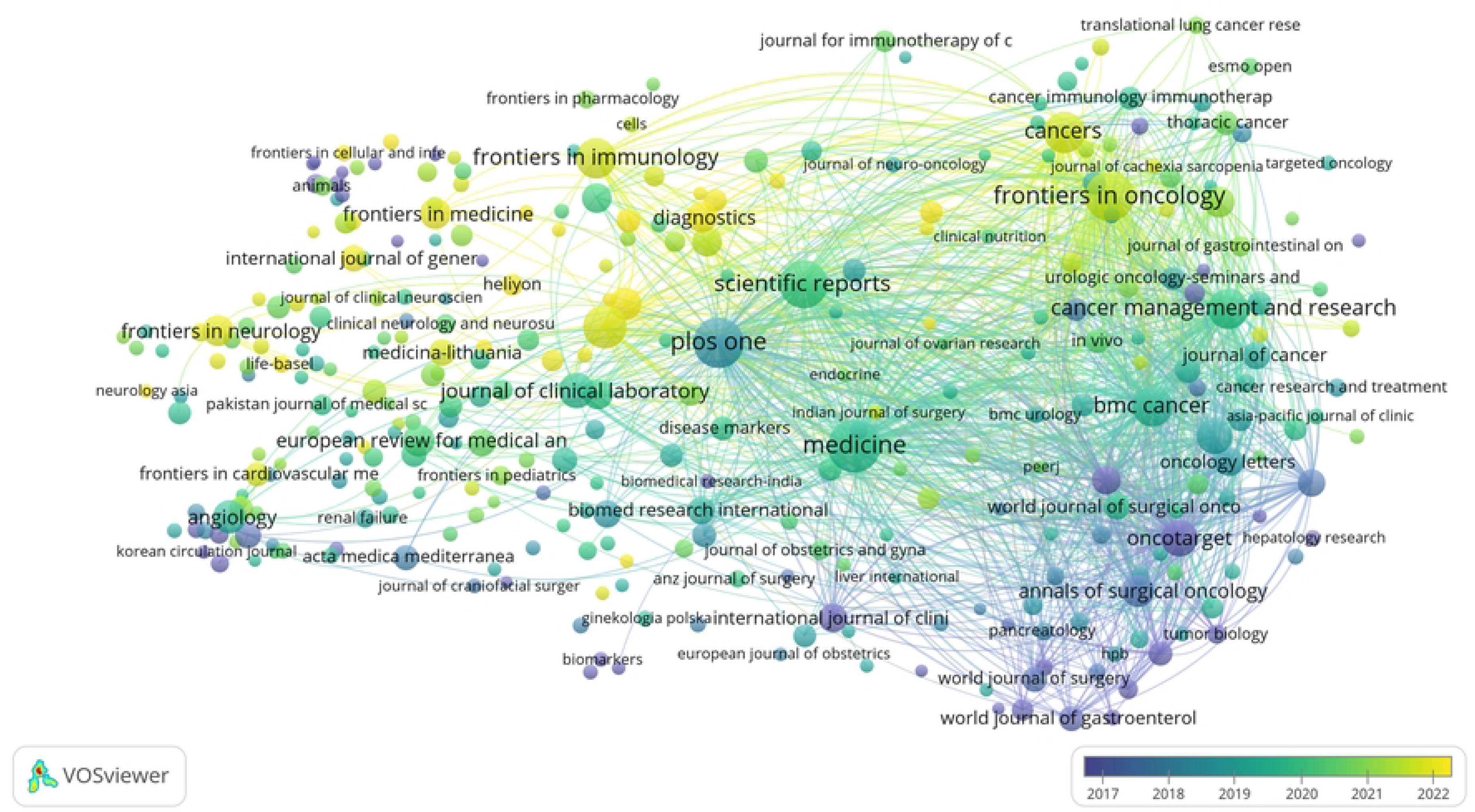

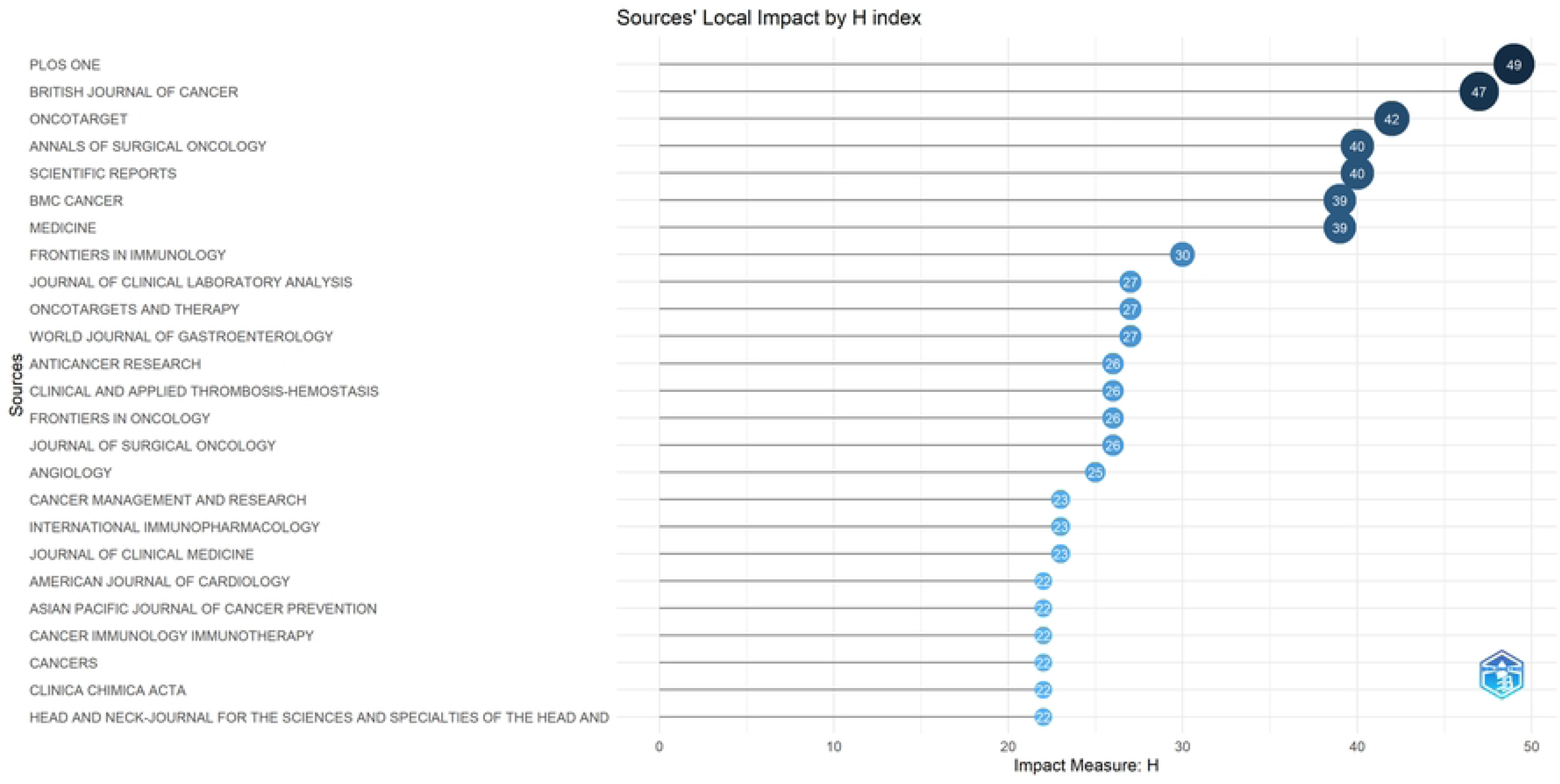

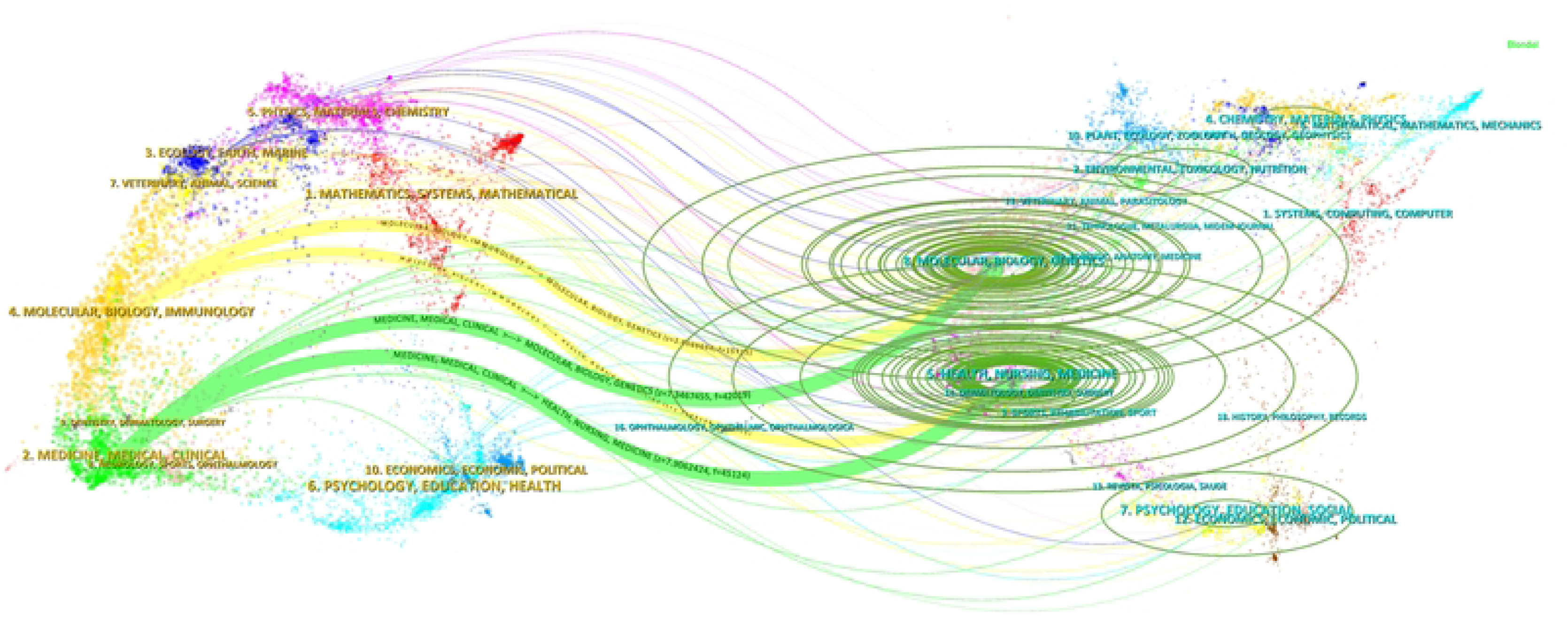

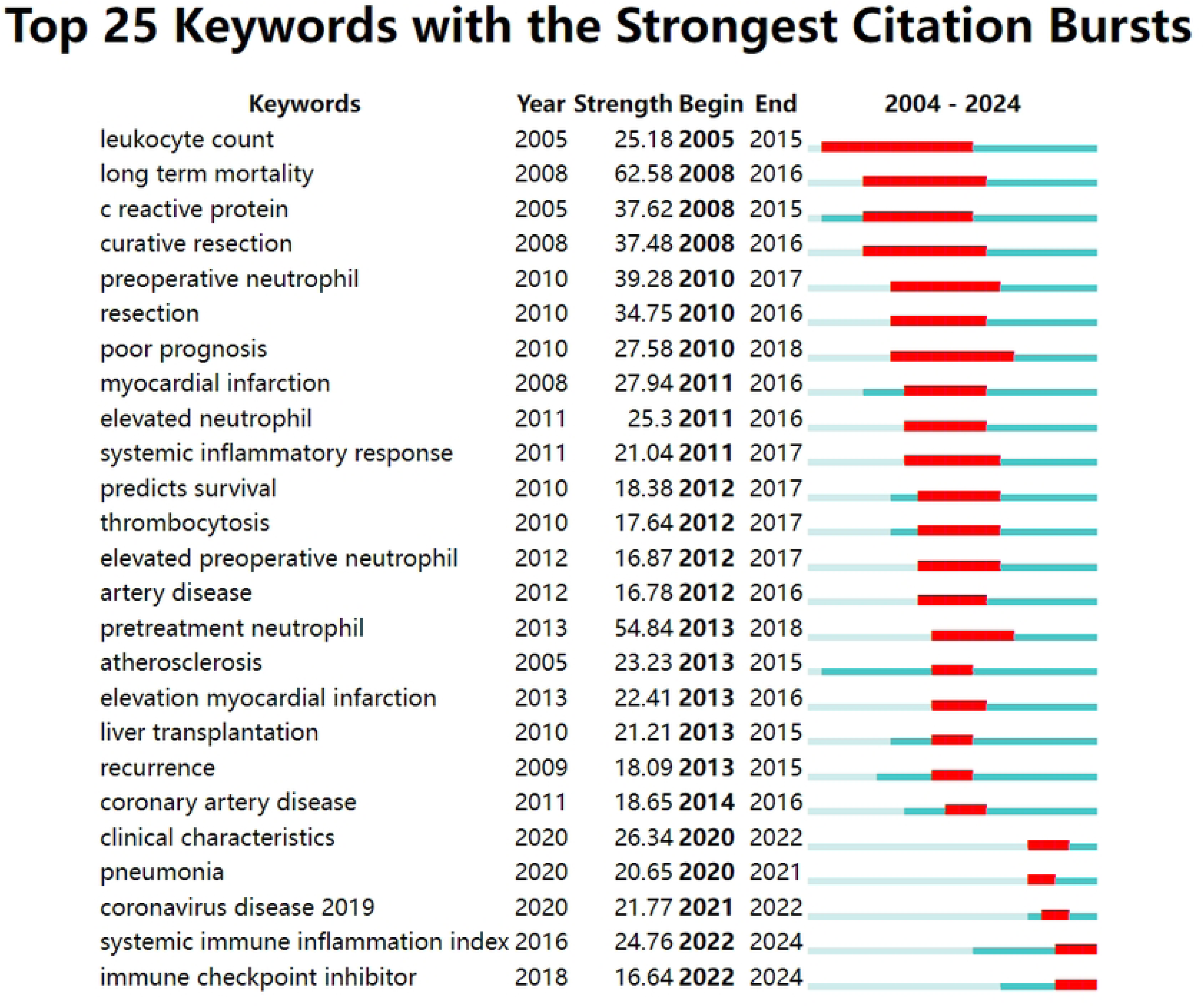

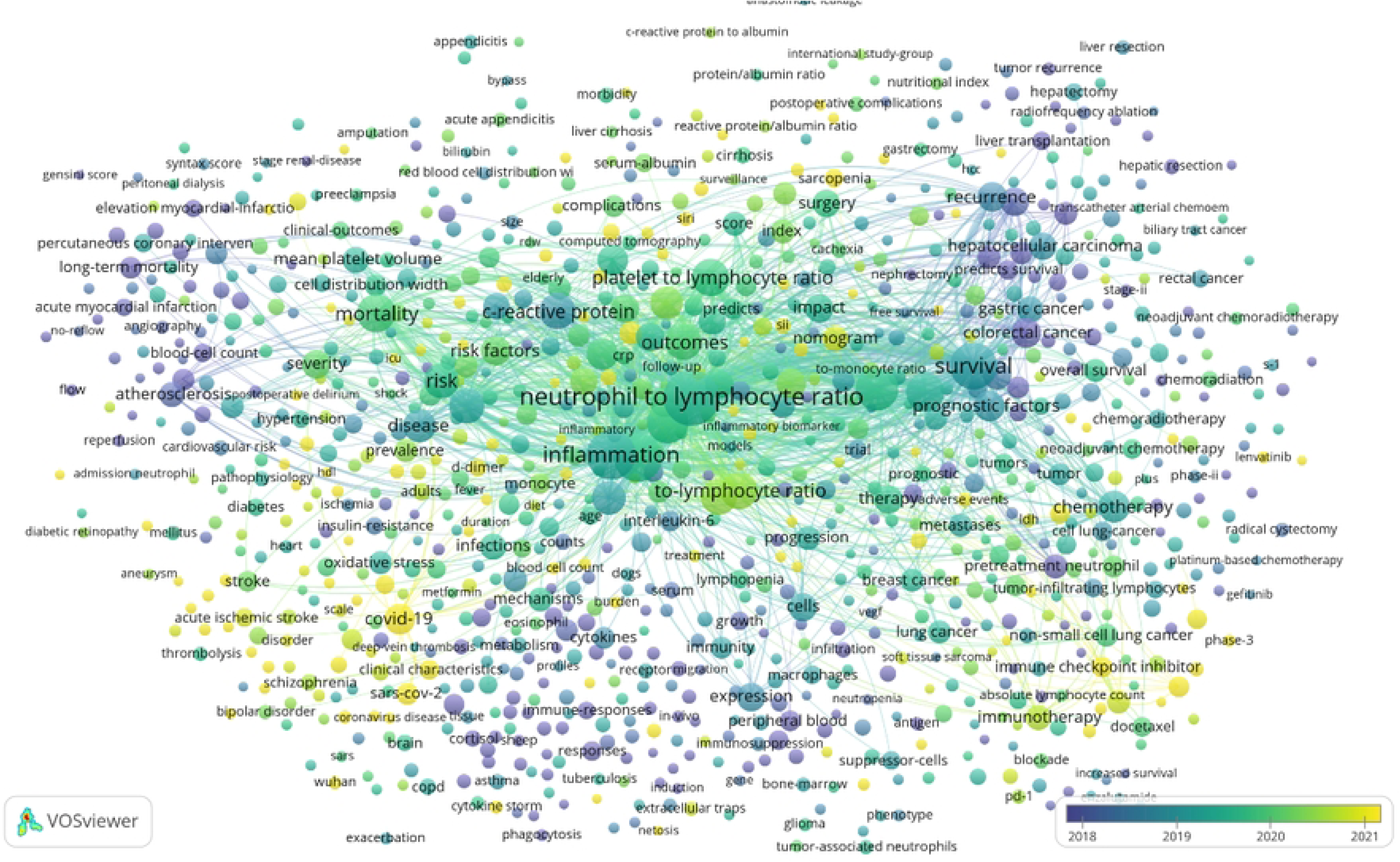

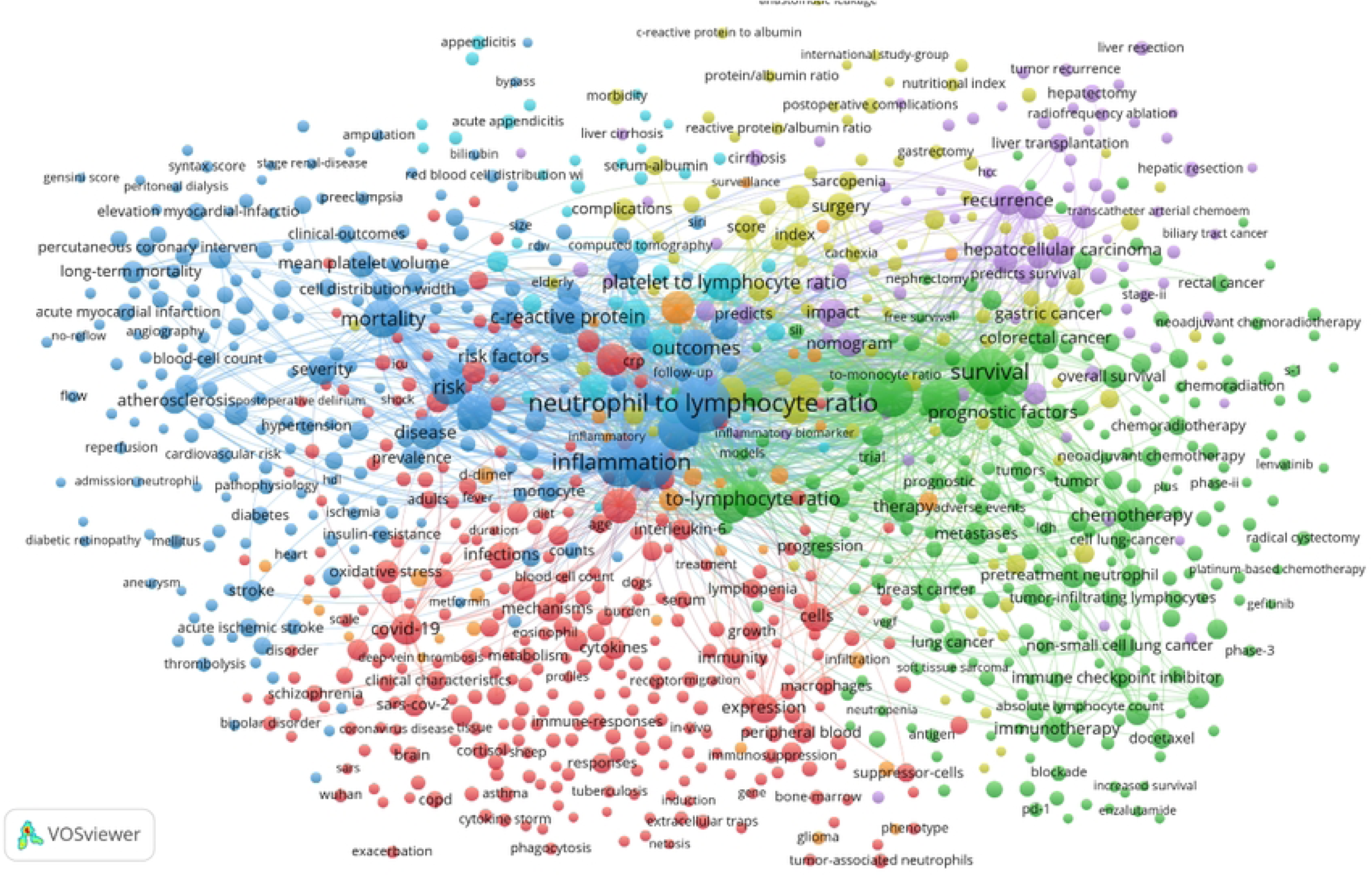

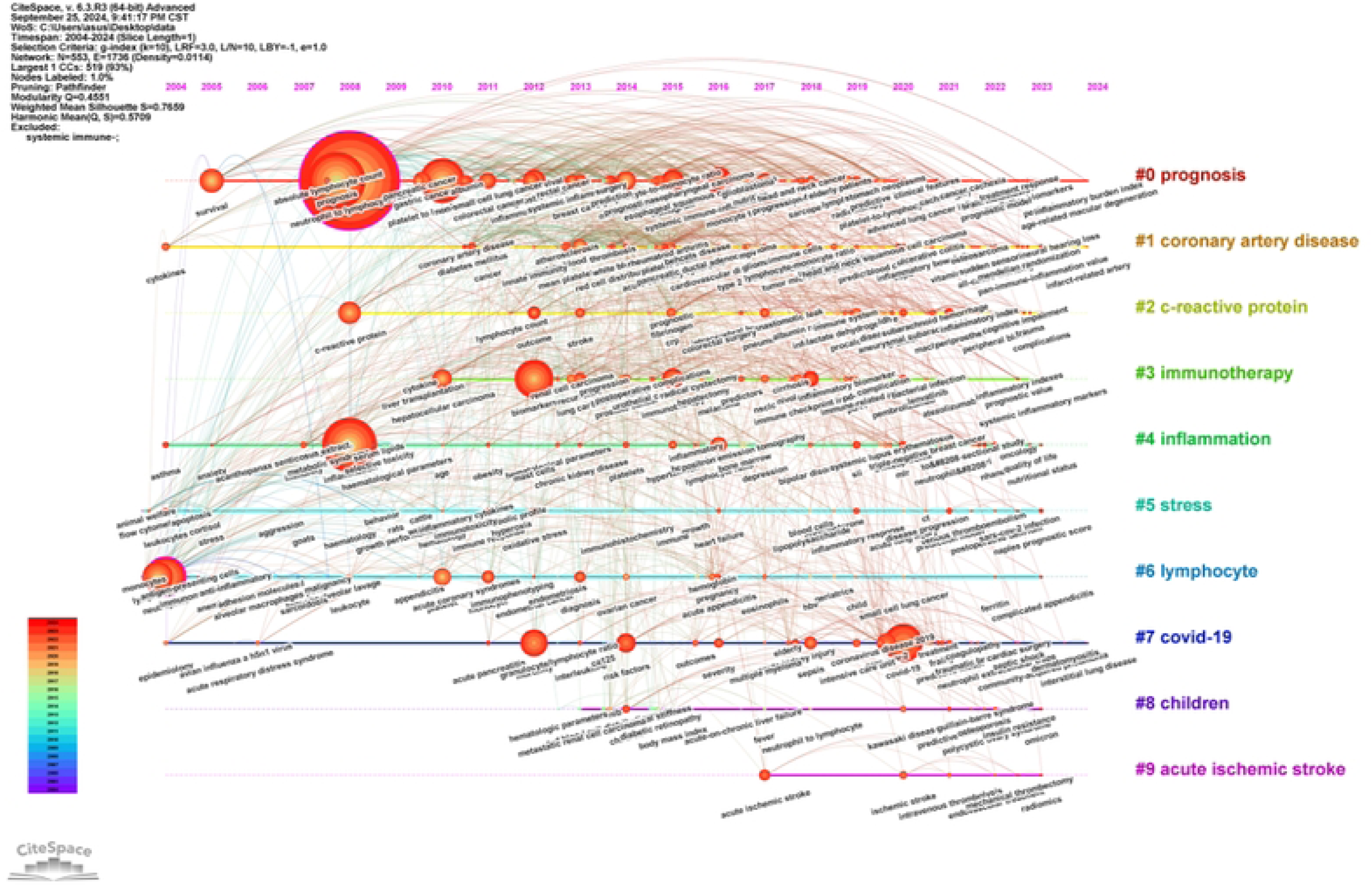

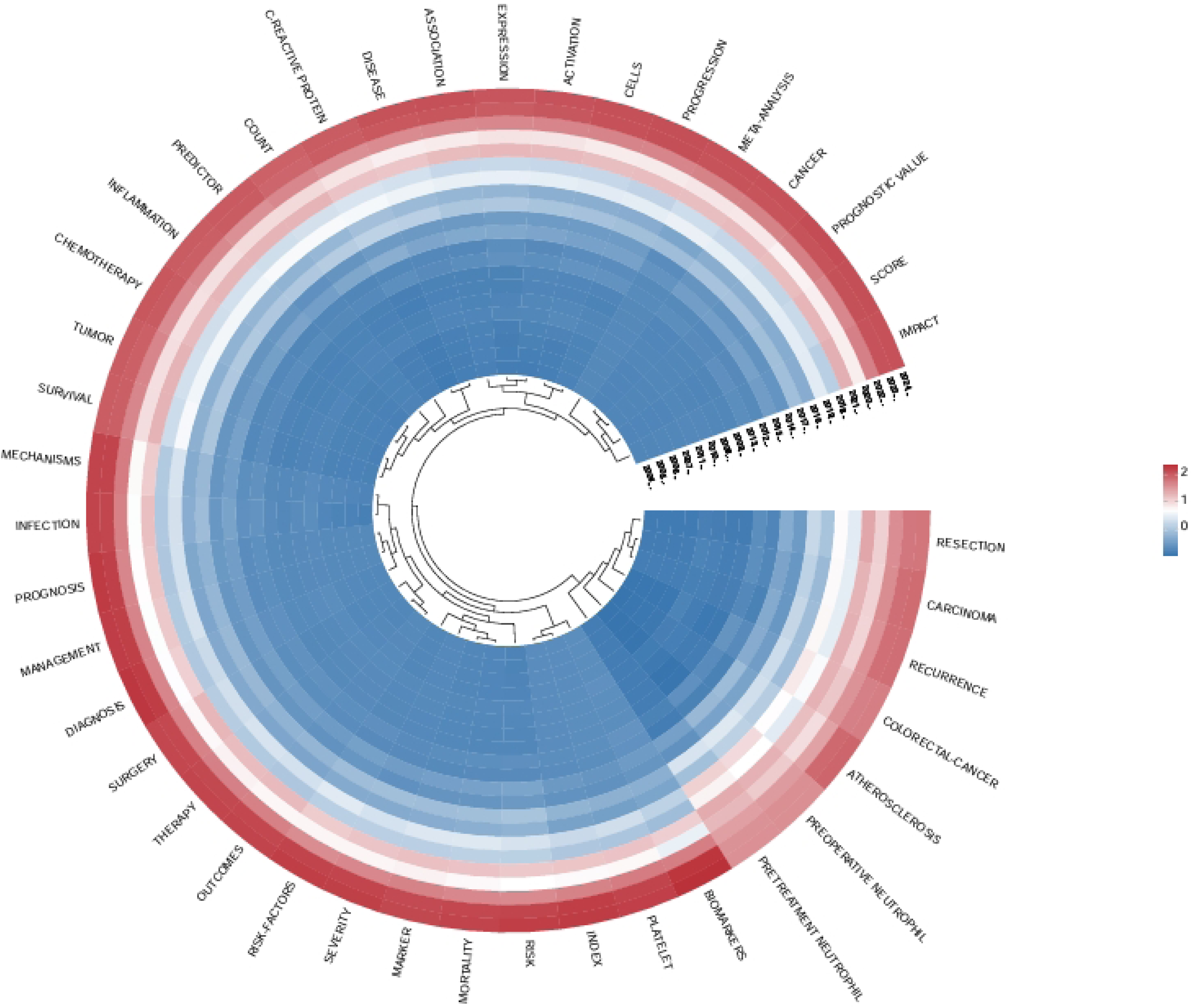

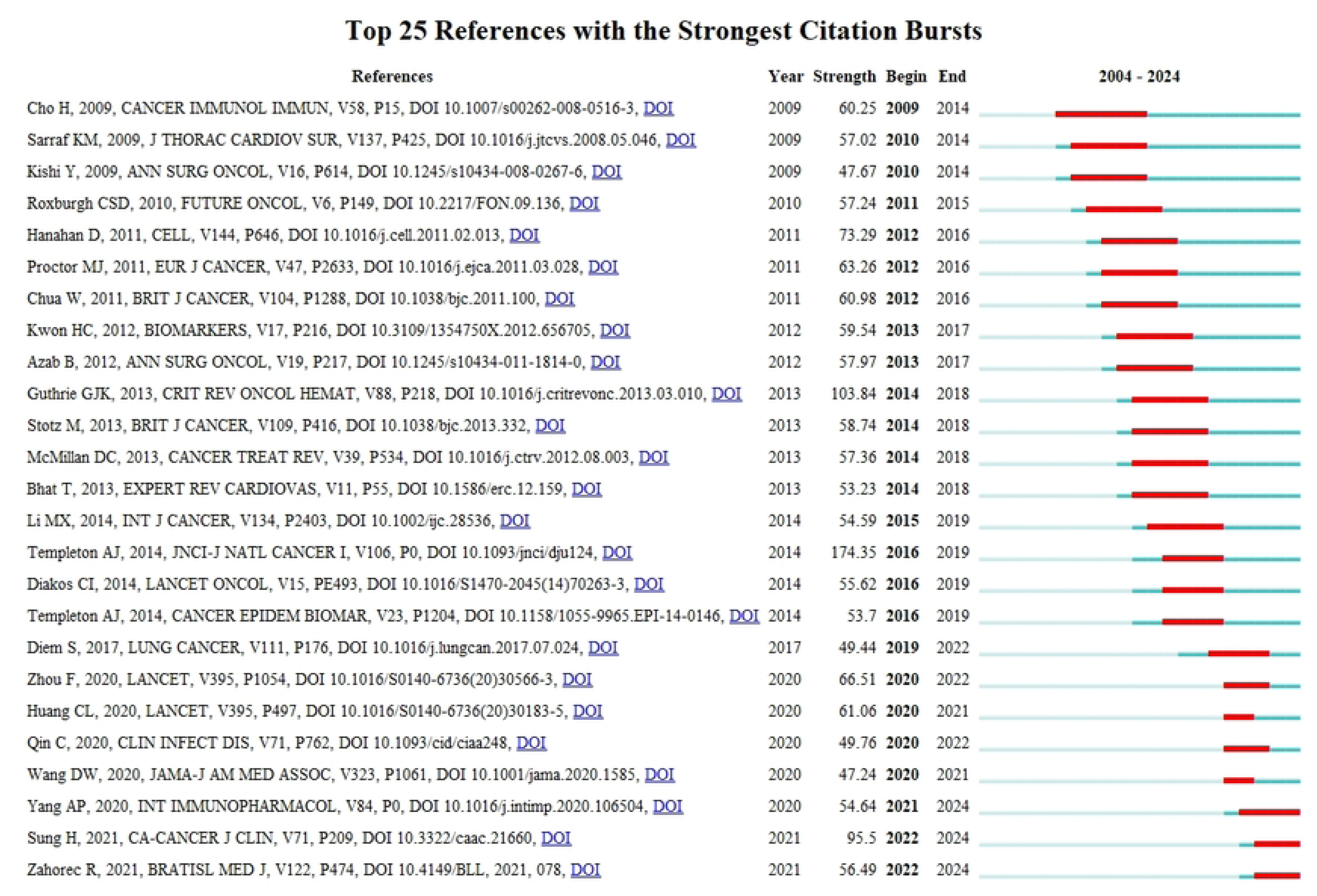

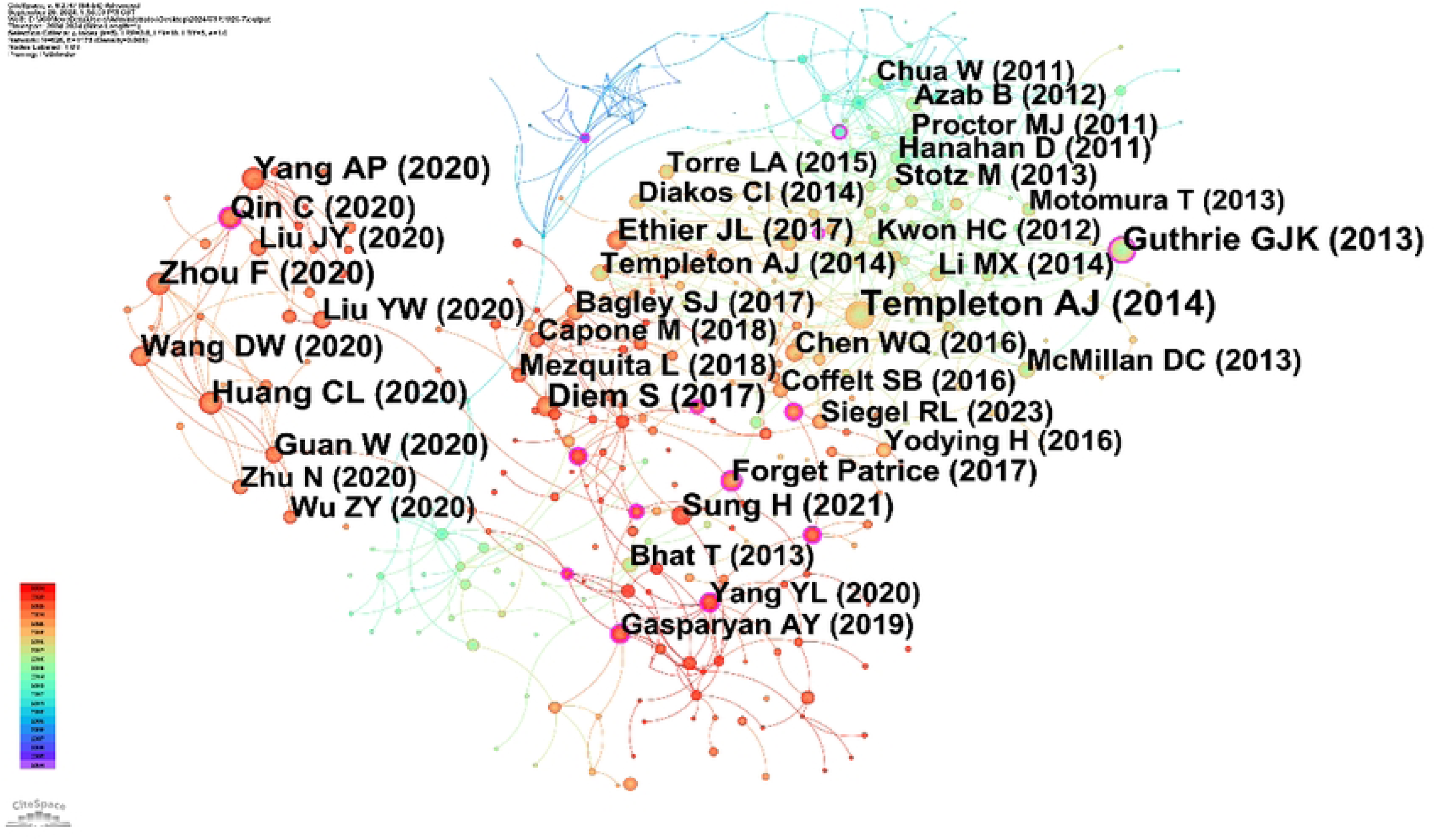

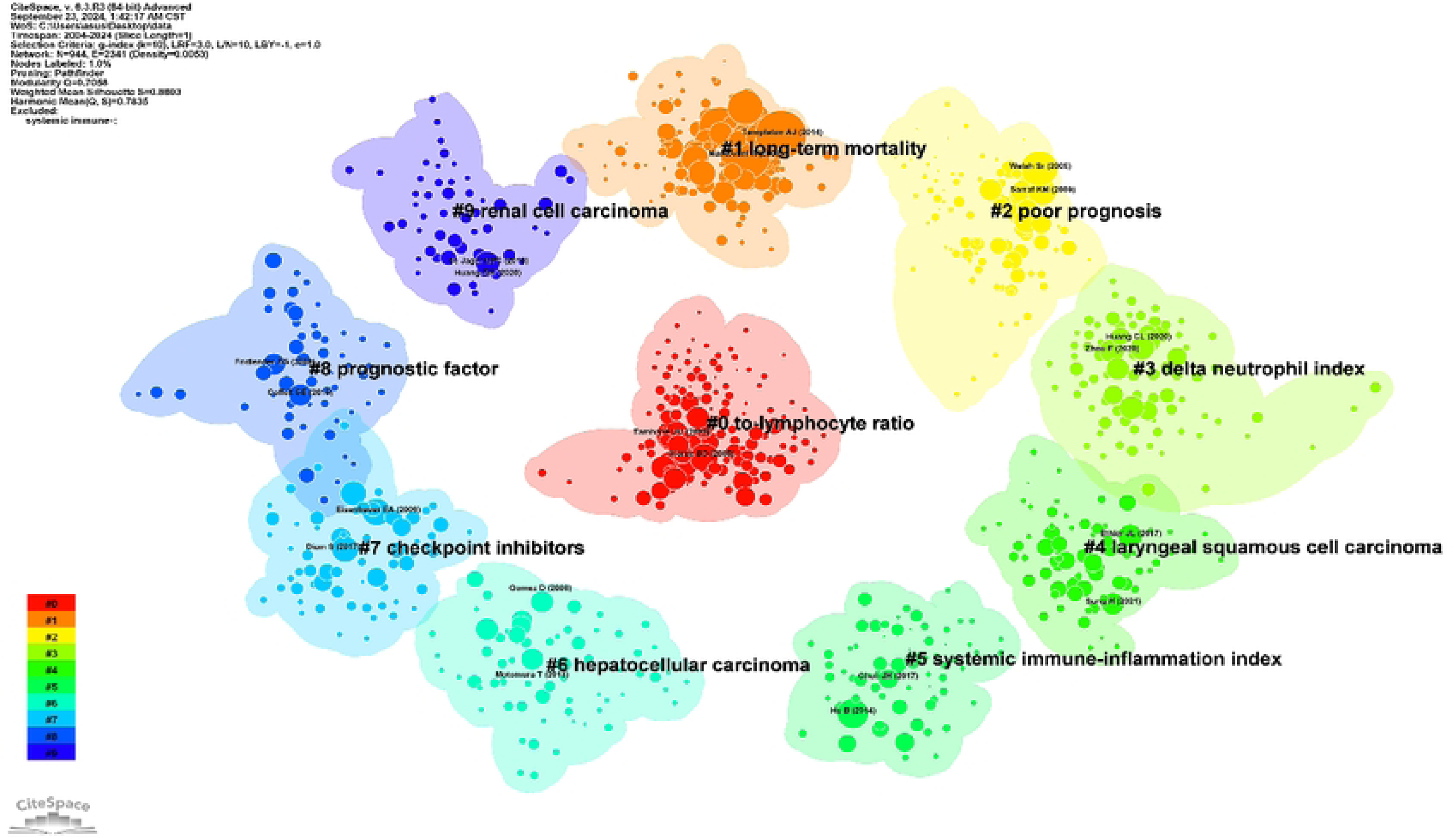

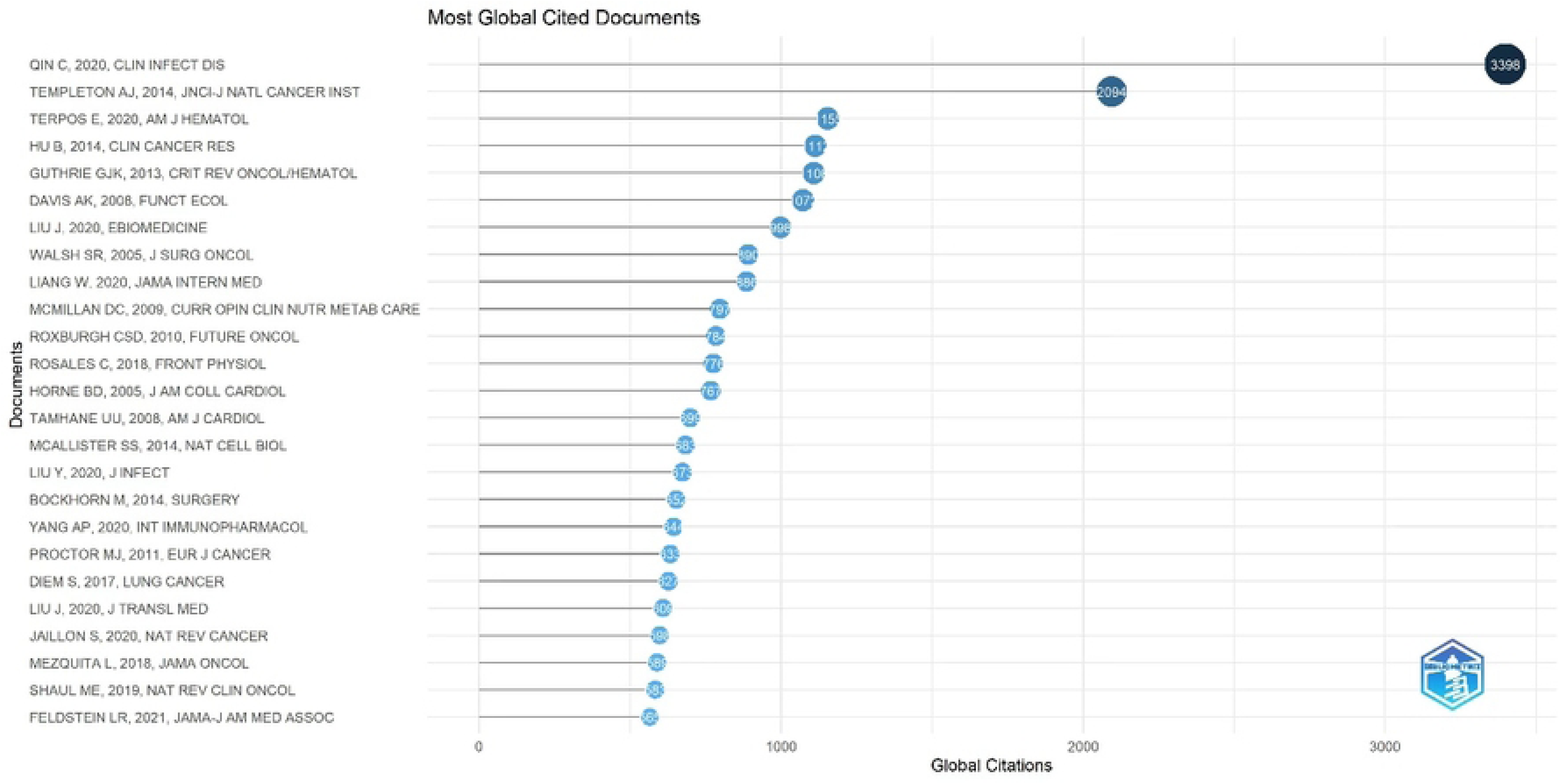

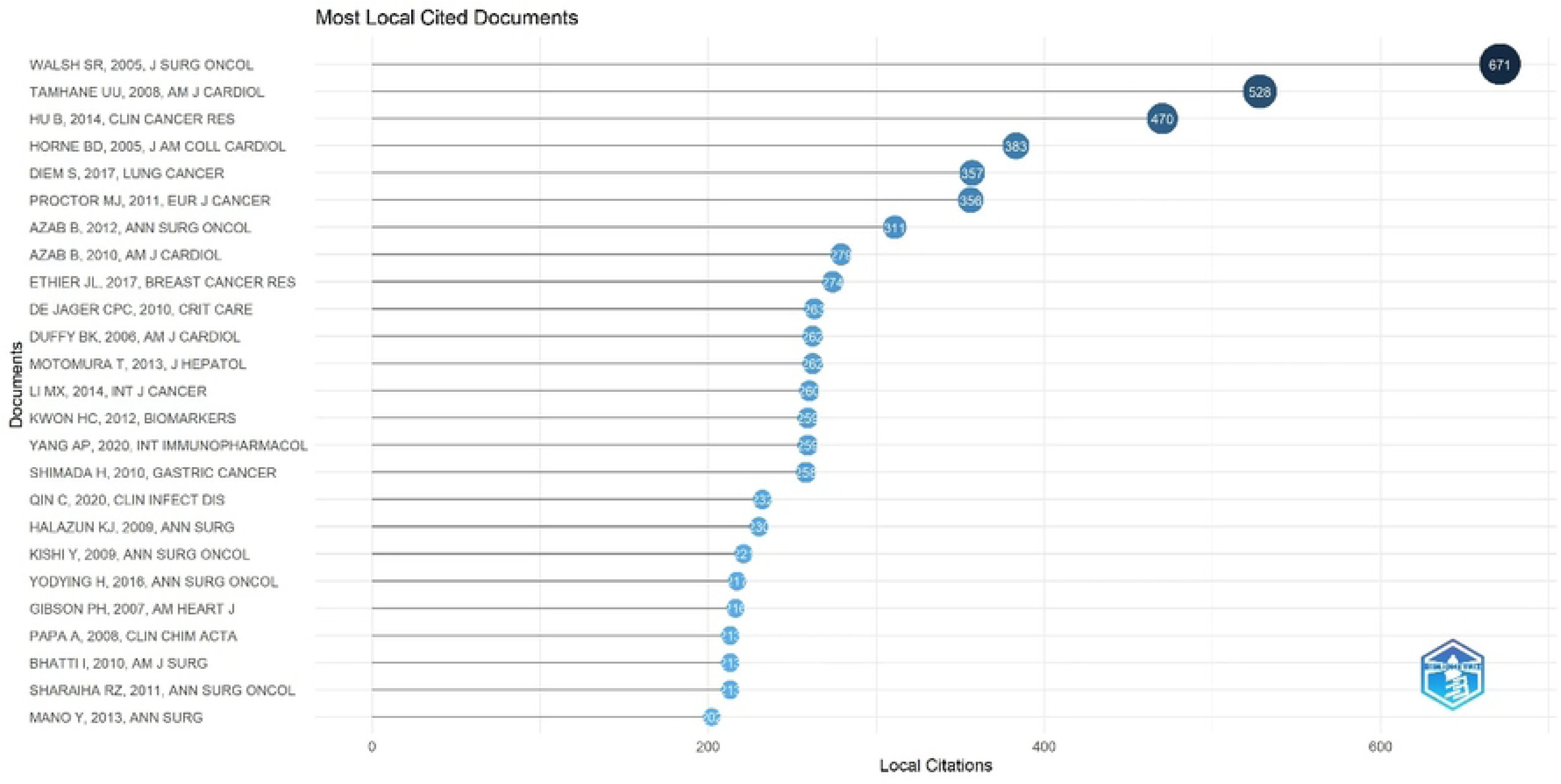

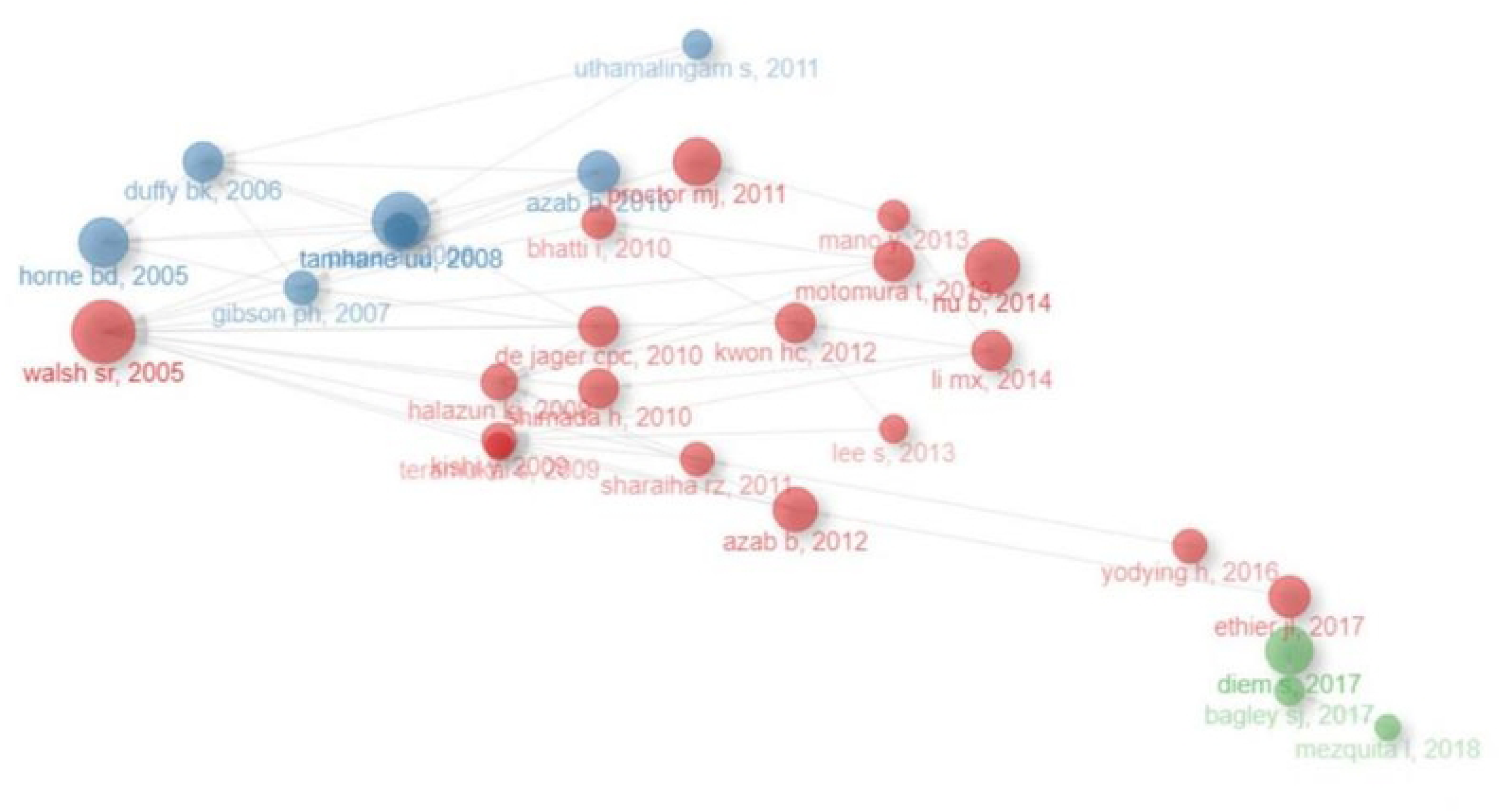

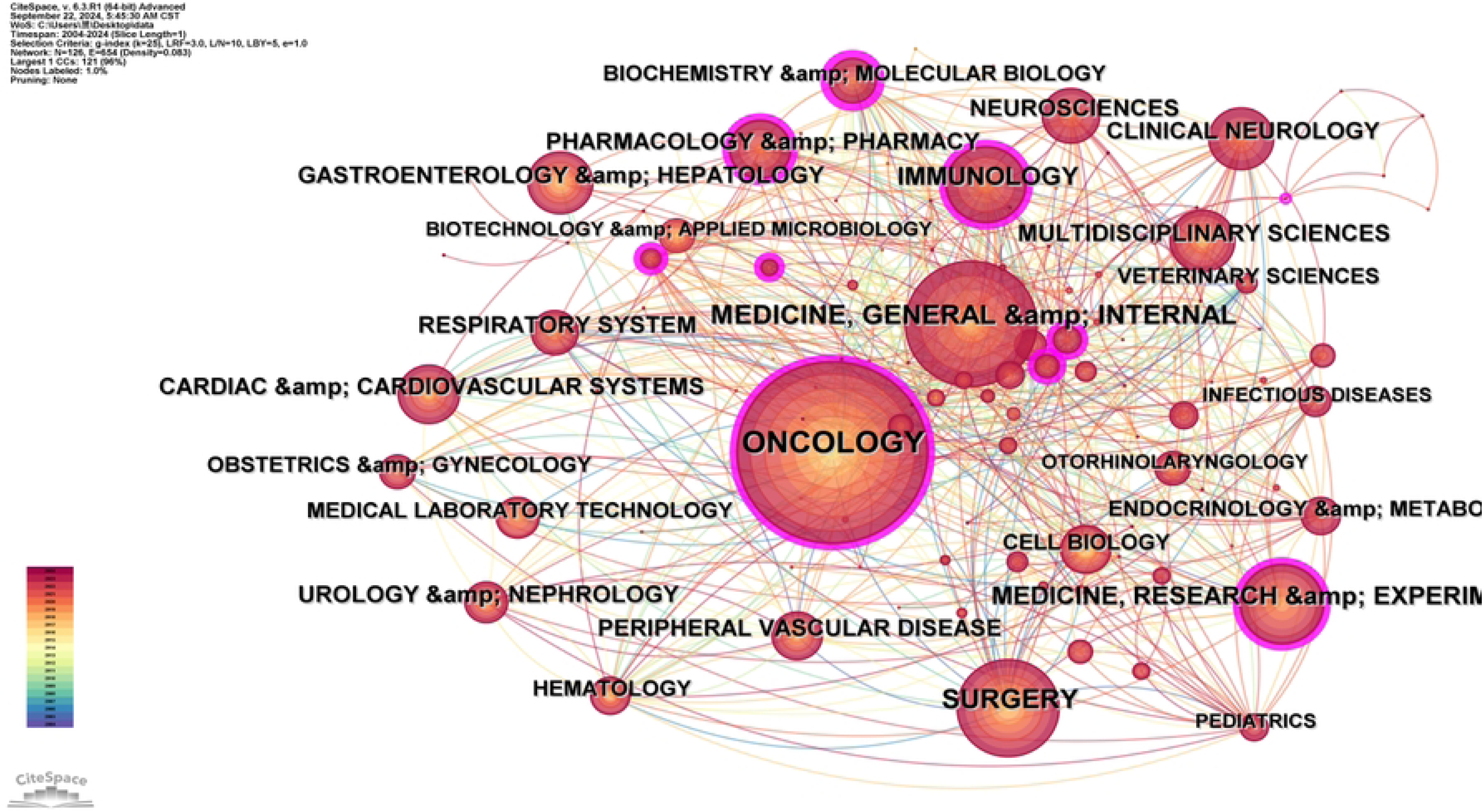

## Data Availability

All relevant data are within the manuscript and its Supporting Information files.

## Statements and Declarations

### Ethical approval and informed consent statements

Not applicable. This article does not contain any studies conducted by any of the authors on human participants or animals. Additionally, this study was granted exempt status by the institutional review board because of its use of deidentified publicly available data. Therefore, there is no need to grant permission in the ethical approval and consent to participate sections. All methods were carried out following relevant guidelines and regulations.

